# The Effects of Development Assistance on Sexual and Reproductive Health Services in Low- and Middle-Income Countries: A Cross-Country Panel Data Analysis

**DOI:** 10.1101/2023.08.24.23294532

**Authors:** Björn Ekman, Jesper Sundewall, Jessy Schmit

**Affiliations:** Department of Clinical Sciences, Malmö, Lund University, Sweden; HEARD, University of KwaZulu Natal, South Africa; Lund University, Sweden

**Keywords:** evaluation, development assistance, sexual and reproductive health, health services, low- and middle-income countries

## Abstract

Using data on 119 low- and lower-middle income countries from 2002 to 2020, we apply fixed-effects (FE) methods to evaluate the impacts of three different classifications of development assistance on access to three types of sexual and reproductive health (SRH) services: access to skilled birth attendance, prevalence of modern contraceptives, and coverage of antiretroviral therapies against HIV/AIDS. The results suggest that aid has had a small, but positive effect on these outcomes over this period. For example, SRH aid has increased service coverage rates by between 0.190 and 0.628 percentage points. The results also indicate that the effect of aid has improved across the period and is larger in low-income countries compared with lower-middle income countries. The findings also indicate that bilateral aid may be somewhat more effective than other types of aid. Importantly, the results suggest that development assistance is more effective if it reaches a certain share of overall health spending. The findings are robust to a series of sensitivity checks. The results of the study lend support to the continued allocation of aid to low-income countries to support the provision of sexual and reproductive health services. Both providers and recipients of SRH aid would be advised to identify ways to improve the effectiveness of development assistance in this area.

## 1 Introduction

Sexual and reproductive health and rights (SRHR) concern the physical, emotional, psychological, and social well-being of individuals in relation to all aspects of sexuality and reproduction. SRHR are central to people’s health and well-being and, more broadly, to the economic development of countries. A growing body of research has highlighted the important benefits to individuals, communities, and societies of investing in SRHR (Starrs, Ezeh et al. 2018b; United Nations Population Fund 2019; Kaiser, Ekman et al. 2021). Consequently, SRHR is seen as a cornerstone of human development and are reflected in several of the sustainable development goals and targets.^1^ Through international agreements, such as the Programme of Action of the International Conference on Population and Development (ICPD), developing countries and their development partners have committed to enhancing investments in SRHR (United Nations Population Fund 2014).

Along with several other dimensions of human health, measures of SRHR outcomes have improved in most low- and lower-middle income countries (LLMICs) over the past decades or so. Among other things, maternal and child mortality rates have gone down, and the health of children and adolescents has improved (WHO, UNICEF et al. 2019; WHO 2023). Studies across countries and regions have shown that these improvements are the result of several factors, including overall economic and social development (Khorrami, Stone et al. 2019). Higher incomes and increased schooling of girls have strengthened the situation of women in many countries which, in turn, have contributed to reduced levels of poverty. However, the critical lesson from the 2018 Guttmacher-Lancet commission on SRHR is that there is an unfinished agenda with respect to improving the SRHR of people in LLMICs and that *“[…] almost all of the 4·3 billion people of reproductive age worldwide will have inadequate sexual and reproductive health services over the course of their lives.“* (Starrs, Ezeh et al. 2018b). The manifestation of this inadequacy is that each year, more than 30 million women do not give birth in a health facility, 45 million women have no or inadequate antenatal care, more than 200 million women want to avoid pregnancy but lack access to modern contraception, and some two million women and men are infected by HIV (Starrs, Ezeh et al. 2018b).

While challenges exist in all countries and regions, it is in low- and lower-middle income countries where the majority of remaining shortfalls are found (Ravindran and Govender 2020; Requejo, Diaz et al. 2020). Table 1 presents recent average values of the main sexual and reproductive health (SRH) service indicators and outcomes across income groups. The data show that the majority of the SRHR disease burden is concentrated in LLMICs. For example, maternal mortality is, on average, ten times higher in low-income countries compared with upper-middle income countries.

**Table 1:** SRH indicators by income group, global averages, and SDG targets.

| SRH Indicators | LIC | LMIC | UMIC | HIC | Global | Year | SDG |
| --- | --- | --- | --- | --- | --- | --- | --- |
| Adolescent fertility (births per 1,000 women ages 15-19) | 91.7 | 40.7 | 29.3 | 11.2 | 41.1 | 2020 |  |
| Contraceptive prevalence, any modern method (% of married women ages 15-49) | 30.8 | 48.3 | 73.5 | NA | 55.75 | 2019 |  |
| Antiretroviral therapy coverage (% of people living with HIV) | 77.6 | NA | NA | NA | 72.0 | 2020 | UC |
| Births attended by skilled health staff (% of total) | 66.7 | 77.3 | 98.6 | 98.7 | 82.6 | 2019 | UC |
| Pregnant women receiving prenatal care at least once (%) | 81.9 | 86.1 | 98.2 | NA | 88.0 | 2019 | UC |
| Maternal mortality ratio (per 100,000 live births) | 453 | 253 | 41 | 11 | 211 | 2017 | 70 |
| Neonatal mortality rate (per 1,000 live births) | 26.4 | 21.8 | 5.7 | 2.7 | 17.0 | 2020 | 12 |
| Under-5 mortality rate (per 1,000 live births) | 66 | 44.9 | 11.2 | 4.9 | 36.6 | 2020 | 25 |

Note: Income groups are indicated as defined by the World Bank in 2021. Year indicates the latest year for which data was available for all income group aggregates in the WDI database. Abbreviations: HIC, high-income countries; LIC, low-income countries; LMIC, lower-middle-income countries; NA, not available; UMIC, Upper middle-income countries; UC, Universal coverage; SDG, Sustainable Development Goals. Source: World Development Indicators (WDI).

Improving SRHR outcomes in the group of lower- and lower-middle income countries will require the effective and sustainable funding of curative and preventive health care services, including provision of contraceptives, access to skilled birth attendants, and HIV treatment programs. However, LLMICs have limited resources to effectively tackle challenges related to SRHR. Consequenctly, most LLMICs are, to a varying degree, relying on aid in the form of Official Development Assistance (ODA^2^) for financing SRHR and other health services (Piemonte 2020). In low-income countries, ODA accounts for, on average, 20 percent of health spending (World Bank 2021). SRHR services are particularly dependent on sustained levels of ODA. To reach the sustainable development goal (SDG) targets and to continue improving outcomes in SRHR, most low-income countries will require substantial additional resources, including ODA and other types of support, over the coming decade.

However, the country-level evidence on the effectiveness of SRH-ODA to improve access to services and health outcomes is still limited. Studies on the effectiveness of ODA have focused on broad development goals such as economic growth and democracy (Herzer and Nunnenkamp 2012; Arndt, Jones et al. 2015; Lee, Yang et al. 2016; Niño-Zarazúa, Gisselquist et al. 2020) while relatively few studies have investigated the effect of ODA on welfare-related outcomes (e.g., health, gender equality) and even less on SRH services and specific SRH outcomes.

Against this background, the purpose of our study is to contribute to an improved understanding of the relationship between SRH-ODA and the provision of sexual and reproductive health services. More specifically, we use a purposive panel data set of 119 LLMICs across the years 2002 to 2020 to estimate the effects of different types of development assistance on three SRHR service outcome indicators for which there are sufficient data: prevalence of modern contraceptive use; skilled birth attendance (SBA); and coverage of anti-retroviral therapies (ART) for HIV/AIDS. The effects of development assistance are measured by modelling the relationship between the different measures of health-related ODA and the service outcomes while controlling for a set of other factors that also may affect these outcomes. Our main analytical approach is to make use of the panel data structure and estimate the effects of ODA using a fixed-effects (FE) estimator which removes all time-constant unobservable effects. Controlling for a set of observable factors that may also affect the outcomes of interest allow for a causal interpretation of the effects of ODA on the identified outcomes.

We find that ODA has had a positive and significant effect on all SRH service indicators over the study period. In particular, SRH-ODA and total health ODA have had an impact on all three services in this period while support classified as reproductive health aid (RH-ODA) has not had a statistically significant effect on ART coverage rates. Furthermore, the effects of ODA have improved over the study period as shown by a stronger effect size in the latter part of the period compared with the earlier part. There is also evidence of a threshold effect of ODA whereby the effect of aid is larger if it reaches a certain share of total health spending. Finally, we find that both bilateral and multilateral aid have had an impact on the service outcomes. The findings are broadly robust to alternative model specifications and to several sensitivity analyses.

In the next section, we discuss the challenges of estimating the effects of aid on relevant outcomes along with an overview of the current evidence on the effects of health ODA on SRHR outcomes. Section 3 presents the analytical framework of the study and describes the data and the methods that we use in the analysis. Section 4 presents the findings of the study with a focus on the main results. Section 5 provides a discussion of the results along with some conclusions.

## 2 Review of the evidence on SRH-ODA

Development assistance operates on several levels and is administered through many different channels. Understanding the effects of ODA on selected and relevant outcomes necessarily requires adopting a multi-pronged approach with different types of analyses that rely on various sources of data and information. Furthermore, the development process as such, of inducing change across a range of economic, social, and political domains with the aim of improving the lives of individuals, is a complex process that is only partly understood. Consequently, any attempt at investigating the effects of development assistance needs to be based on a sound analytical or theoretical framework and employ rigorous methods using relevant data, while acknowledging the scope and limitations of the analysis.

Given these general considerations, approaches to evaluations of development assistance can broadly adopt either of two perspectives: macro-level analysis or micro-level analysis. Macro-level analyses look at the effects of ODA on national, sub-national, or sector (meso) levels. Micro-level analyses investigate the effects of specific interventions or projects on the level of individuals or households. Furthermore, studies can use either qualitative or quantitative (statistical) research methods, or a combination of these two types of methods (mixed-methods). Each of these approaches can address specific questions and thereby contribute in different ways to the overall evidence of the effects of ODA on relevant outcomes.

The general aim of a quantitative approach, such as statistical or econometric modelling, is to obtain an estimate of the effect of an intervention (such as a reform, policy program, or, in this case, the allocation of financial resources) on some relevant outcome (such as GDP growth, poverty reduction, or as in the current case, changes in service utilization). A challenge, and limitation, when conducting a quantitative analysis on the macro-level is that the unit of analysis cannot be randomly allocated to, in this case, either receive aid or not receive aid. An implication of the inability to randomize countries is that the estimated effects will only provide indications of the strength and size of the effects of aid on the outcomes. As the changes in the outcomes of interest are most likely determined by several factors, some of which can be measured with the available data and others that cannot be measured, it is important to adopt methods that can limit the problems of non-random allocation. Nonetheless, along with other types of evidence, the findings of such analyses will constitute a valuable contribution to the general question of the role of development assistance.

### 2.1 Existing evidence of the effects of ODA on sexual and reproductive health outcomes

The research literature on the effects of ODA on SRHR indicators and outcomes is large and consists of studies across a range of study designs and methods. For the purposes of this study, we included studies that aimed to report the effects of ODA using some quantitative research design. Outcomes of interest for the review included gender-based violence (GBV), maternal and new-born health, abortion, HIV/AIDS, other sexually transmitted diseases, reproductive cancer, family planning, female genital mutilation (FGM), and menstrual health. We reviewed literature covering the years 2002-2020 to match the period of the data collected for this study.

In all, 25 studies were included in the review, the majority of which focused on infant health and the treatment of HIV/AIDS. Six studies focused on maternal health and related service provision. Three studies analyzed the effects of ODA on family planning and only one study addressed abortion as an outcome. Notably, gender-based violence, reproductive cancers, sexually transmitted diseases other than HIV/AIDS, FGM and menstrual health were absent from the results, indicating considerable gaps in the current literature.

Overall, the evidence suggests that ODA has had a positive effect on the outcomes studied, i.e., infant health, maternal health, HIV/AIDS, and family planning and abortion. That said, there are studies that did not find a positive effect of ODA. Results of the effect of ODA on infant mortality rates are mixed where four studies found evidence that ODA significantly reduces infant mortality rate (IMR) (Islam 2003; Kotsadam, Østby et al. 2018; Win and Cho 2018; Akinlo and Sulola 2019), while three other studies do not detect any significant effect of ODA on IMR in their analysis (Arndt, Jones et al. 2015; Pallas and Ruger 2017; Toseef, Jensen et al. 2019).

With respect to maternal health one study found evidence for ODA leading to a significant reduction in maternal mortality (Pickbourn and Ndikumana 2016). Another study that looked at UK aid found that health and family planning programming have saved 103,000 women’s lives between 2010 and 2014 (Friberg, Baschieri et al. 2017). However, two other studies reported mixed results, with one study reporting contradictory effects depending on the type of financing provided by the World Bank (Coburn, Reed et al. 2017), and another study found that only aid allocated directly to the reproductive or maternal health sector reduced maternal mortality (Banchani and Swiss 2019).

Regarding HIV/AIDS, the evidence is similarly mixed. Two studies reported that ODA has an effect on reducing HIV/AIDS-related mortality (Nunnenkamp and Öhler 2011; Hsiao and Emdin 2015). However, evidence for ODA lowering HIV/AIDS incidence is inconclusive, (Nunnenkamp and Öhler 2011; Lee, Yang et al. 2016). Two studies evaluating the effect of ODA on HIV/AIDS prevalence found no evidence (Lee, Yang et al. 2016) and mixed evidence (Yogo and Mallaye 2015) of a significant effect of ODA, respectively. One study showed that increases in foreign aid for HIV contributed to increases in antiretroviral therapy coverage in 13 African countries (Bendavid, Leroux et al. 2010).

For family planning evidence is even more limited. One study observed that ODA significantly reduces adolescent fertility rates (Zhuang, Wang et al. 2020), and another showed that USAID family planning funding significantly increased the prevalence rates of modern contraceptives (Shepard, Bail et al. 2003). Finally, only one study evaluated the effect of ODA on abortion under the Mexico City Policy program (Brooks, Bendavid et al. 2019).

Based on the review of the included studies, we find that the quality of the evidence for the effects of ODA on SRH outcomes is moderate largely because none of the studies included in the review were experimental in study design. Most studies (n=14) applied a fixed-effects analysis using panel data. Three studies adopted a quasi-experimental approach, using a difference-in-difference (DID) identification strategy. Although they do not employ randomization, both fixed-effects and DID provide some evidence as to a possible causal relationship between ODA and SRH outcomes. One study used a structural causal model approach, and the remaining studies (n = 7) were observational in design. The relatively large differences between the studies in terms of study design, definition of ODA and the variety of outcomes measured limit the ability to state conclusively about the causal effect of ODA on SRH outcomes. The current study contributes to the evidence by rigorous analysis of a large panel data set as described in the next section.

## 3 Analytical framework, data, and methods

### 3.1 Analytical framework

The statistical analysis of this study is guided by an analytical framework that describes the process of investing in health broadly, and in SRHR more specifically. The framework builds on the existing knowledge and understanding of this process and helps to identify the objectives of the study. The analytical framework is illustrated in Figure 1.

**Figure 1:**
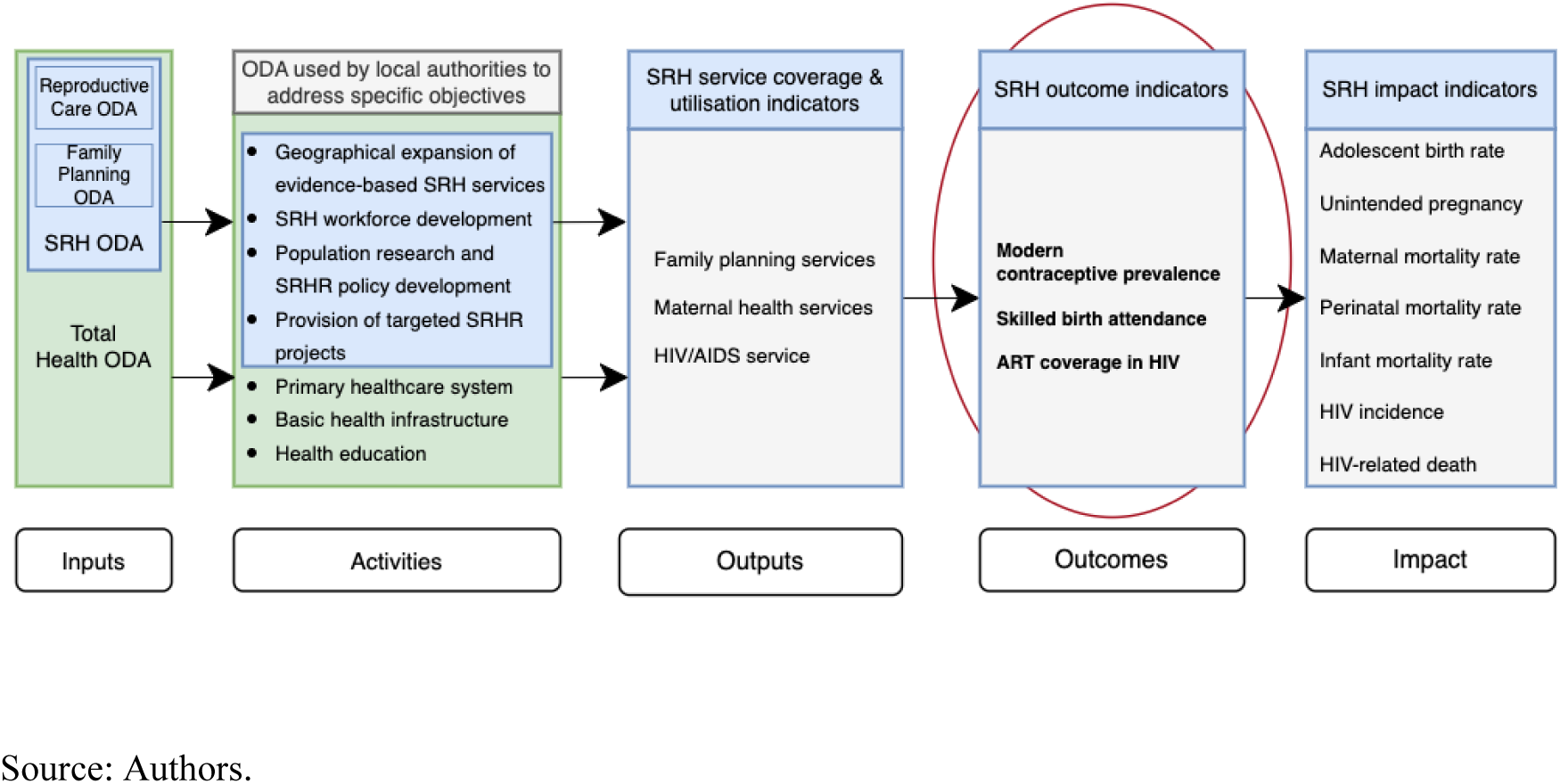
Analytical framework.

As noted above, we adopt this approach to investigate the effects of various types of development assistance for health on a set of measures of access to and coverage of SRH services. The framework captures the process of financial and other inputs to enhance the coverage of SRH services, which, in turn, is assumed to impact the longer-term SRH outcomes, including mortality rates, pregnancy rates, and HIV-related morbidity and mortality rates. However, attributing the direct effect of development assistance to these distal outcomes is difficult (Addison, Mavrotas et al. 2005; Sumner and Glennie 2015). Consequently, we focused our analysis on the effects of ODA on SRH service outcomes on which there are reliable data.

### 3.2 Data

In this study, we evaluate the effects of development assistance on the coverage of key sexual and reproductive health services of recipient populations on a macro level, i.e., at the country level, which is the study’s unit of analysis. To meet this end, we collected openly available country level data on a range of SRH and other indicators. The main databases from which the data were collected include the World Development Indicators (WDI), the World Governance Indicators (WGI) of the World Bank, and the OECD’s Creditor Reporting System (CRS) database for ODA data.

A total of 119 countries were included in the sample. All these countries were classified as either low-income or lower-middle-income by the World Bank in 2002.^3^ Data on these indicators cover the years 2002 to 2020, i.e., a period of 19 years. The starting year of 2002 was from when data on ODA disbursements for the sample became available and 2020 was the most recently available year for the data at the time of data collection. See also Tables A.1 and A.2 in Appendix A. Table 2 presents descriptive statistics across the two income groups and for the total sample; additional descriptive statistics are presented in Appendix B.

**Table 2:** Descriptive statistics of sample, 2002-2020 by income group in 2002.

| VARIABLES | LIC 2002 |  | LMIC 2002 |  | Total Sample |  |
| --- | --- | --- | --- | --- | --- | --- |
|  | N | Mean<br>2002-2020 | N | Mean<br>2002-2020 | N | Mean<br>2002-2020 |
| **Panel A: Outcomes** |  |  |  |  |  |  |
| Skilled birth attendance (%) | 428 | 68.22 | 522 | 93.62 | 950 | 82.17 |
| Contraceptive Prevalence (%) | 334 | 30.55 | 171 | 46.86 | 505 | 36.07 |
| ART coverage (%) | 1,176 | 23.38 | 661 | 29.00 | 1,837 | 25.40 |
| **Panel B: ODA** |  |  |  |  |  |  |
| Total health ODA, US\$ millions | 1,226 | 171.4 | 1,009 | 42.21 | 2,235 | 113.1 |
| SRH-ODA, US\$ millions | 1,226 | 74.93 | 960 | 20.01 | 2,186 | 50.81 |
| RH-ODA, US\$ millions | 1,223 | 12.74 | 892 | 1.999 | 2,115 | 8.211 |
| Total health ODA, US\$ pc | 1,226 | 10.25 | 1,009 | 15.69 | 2,235 | 12.71 |
| SRH-ODA, US\$ pc | 1,226 | 3.851 | 960 | 3.495 | 2,186 | 3.695 |
| RH-ODA, US\$ pc | 1,223 | 0.688 | 892 | 0.348 | 2,115 | 0.545 |
| **Panel C: Covariates** |  |  |  |  |  |  |
| GDP per capita (constant, PPP) | 1,148 | 4,027 | 970 | 10,231 | 2,118 | 6,868 |
| GHE as share of CHE | 1,106 | 29.42 | 918 | 53.52 | 2,024 | 40.35 |
| Government Effectiveness (0-5) | 1,224 | 1.601 | 1,004 | 2.166 | 2,228 | 1.855 |
| Corruption Control (0-5) | 1,227 | 1.679 | 1,013 | 2.145 | 2,240 | 1.890 |
| Population Density (per sq. km) | 1,215 | 114.3 | 999 | 136.9 | 2,214 | 124.5 |
| Primary school enrolment (% gross) | 922 | 102.1 | 799 | 104.9 | 1,721 | 103.4 |
| Female labour market part. (%) | 1,235 | 57.98 | 931 | 41.45 | 2,166 | 50.87 |
| Number of countries | 65 |  | 54 |  | 119 |  |

Note: CHE = Current health expenditure; GHE = Government health expenditure.
Source: OECD-CRS, WDI, WGI.

In all, the sample includes 2,261 country-year observations. Around 55 percent of the countries were classified as low-income in 2002. During the sampling period, some of the countries transitioned from one income classification to another, mainly from low-income to lower-middle income. While service coverage is generally higher in lower-middle income countries than in low-income countries, these transitions are not expected to affect the analytical approach or the final results.

Figure 2 shows the changes in the average service coverage rates for the sample countries over the period of analysis. The data show that coverage of both skilled birth attendance and ART have increased over the study period. However, it should be noted that ART coverage started from a level close to zero as it was not broadly offered until around 2002-2003.

**Figure 2:**
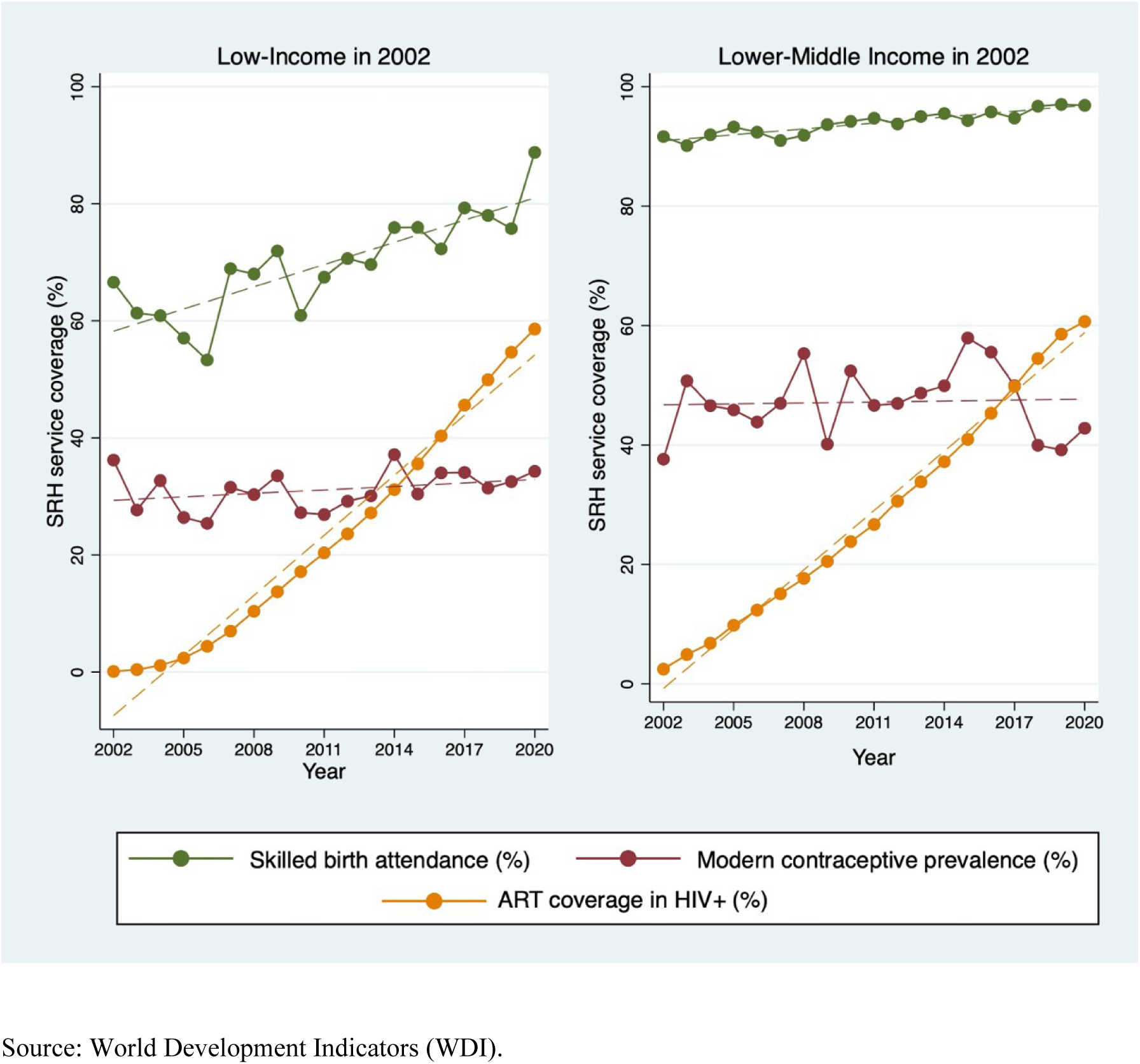
Average SRH service coverage rates, LIC and LMIC, 2002–2020.

Contraceptive prevalence on the other hand has remained at relatively similar levels over the period, in both LICs and LMICs.

Three measurements of ODA were used in the analysis, namely total health ODA, sexual and reproductive healthcare ODA (SRH-ODA), and reproductive healthcare ODA (RH-ODA). Each measurement consists of one or more “sector codes” in the OECD CRS database. All disbursements, including grants and loans, under each of the sector codes were included. Total Health ODA consists of sectors 120 and 130 combined, SRH-ODA includes all ODA under sector code 130, and RH-ODA consists of disbursements under sector code 13020. Including different measures of ODA was considered important because of the overlaps between ODA funded programs (for example some broader health programs also support SRH, this is particularly the case for maternal health) and because of inconsistencies in reporting ODA statistics (for example, some SRH support is reported as *basic health care* and vice versa). In addition, by including several measurements of ODA, we can assess the effects of different health ODA classifications and reduce the likelihood of the problem of measurement error which may affect the accuracy of the effect estimates.

Figure 3 shows annual disbursements of ODA under OECD-DAC sector code 130, sexual and reproductive health (SRH) and its sub-categories over the study period.

**Figure 3:**
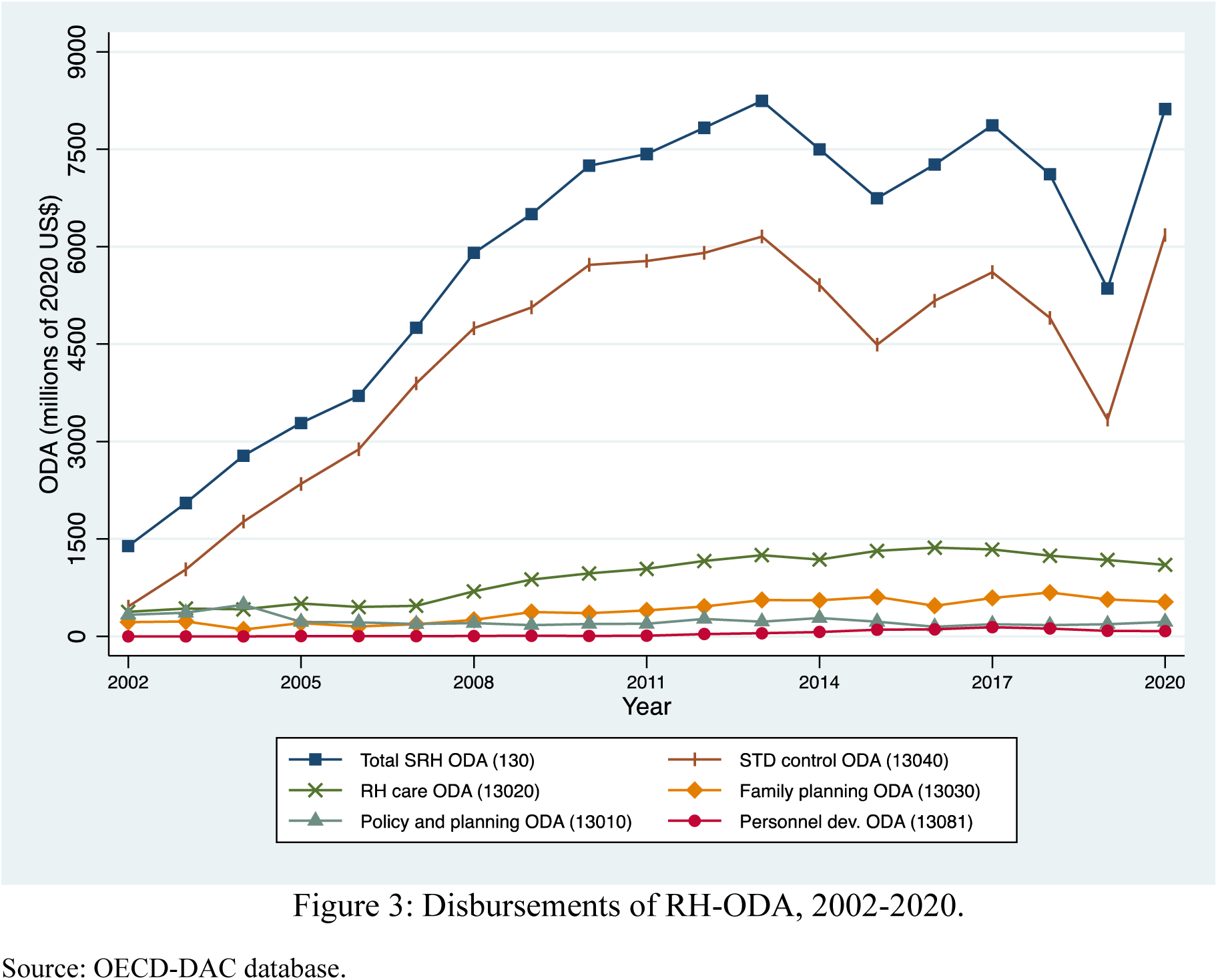
Disbursements of RH-ODA, 2002-2020.

The large increase in disbursements to sector code 13040, STD control ODA, is all but exclusively due to the increases in the treatment of HIV/AIDS in the form of anti-retroviral therapies (ART). All other areas of SRH support have seen only modest increases or no changes in disbursement levels across this period.

### 3.3 Methods

Evaluating the effects of development assistance on the identified outcomes at the country level requires overcoming three analytical challenges. First, measures of ODA are not always accurate. This may be particularly so for development assistance to specific areas of SRH but also with respect to measures of service coverage and possibly also other variables. Consequently, the imperfect measures of the main analytical variable may pose a problem for the validity of the estimation results. Second, the econometric model may be affected by the problem of endogeneity (or unobserved confounding). A common reason for endogeneity in empirical analysis is the potential failure to include a factor that determines the outcome and that is also associated with any of the included variables. The problem of endogeneity due to omitted variable would lead to biased and inconsistent estimates of the effects (Wooldridge 2010). And finally, the estimation may also be affected by the problem of simultaneity if the values of the outcome variable are affected by the values in preceding periods through some dynamic aspect of the data generating process (DGP).

To address these methodological challenges, our main approach is to use fixed-effects (FE) panel data analysis. This approach allows for the estimation of the effects of development assistance on our selected outcomes while controlling for both observable factors and for time-constant unobserved factors, i.e., factors that are relatively stable over time in a certain country but that we do not have data on. Examples of such factors are religious beliefs, social norms and values, the quality of the countries’ institutions and health system, and public management capacity. These are factors that may affect the recipient country’s ability to make effective use of the aid received. The main FE model takes the following form:

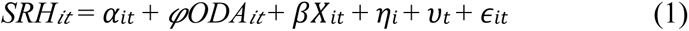

The subscripts *i* and *t* denote country and year, respectively. *SRH𝑖𝑡* measures the SRH services of interest in the study. *ODA𝑖𝑡* measures the sectoral aid disbursed to a country in a certain year. In the main model ODA is expressed in per capita terms to adjust for population size. The coefficient 𝜑_𝑖𝑡_ is the main ODA effect estimate of interest. 𝑋_𝑖𝑡_ is a vector of country-level covariates that might impact SRH services in a country, as shown by previous studies or suggested by theory. The country fixed effects (𝜂_𝑖_) control for unobserved time-invariant factors at the country level. The time fixed effects, 𝜐_𝑡,_ account for overall time trends across countries. Finally, 𝛼_𝑖𝑡_ and 𝜖_𝑖𝑡_ are the intercept and the idiosyncratic error term, respectively.

Model 1 was estimated using a linear-linear specification. i.e., effect estimates represent the average percentage point change in service coverage associated with a 1 USD per capita increase in sectoral health ODA. The covariates GDP per capita and population density (per sq. km) were log transformed to facilitate the interpretation of coefficients.

The FE model was compared with alternative models. The Durbin–Wu–Hausman test results suggested that FE models were preferred over the random-effects (RE) model. In turn, for each combination of ODA measure to SRH service indicator, the random-effect models were favored over the pooled OLS models based on the Breusch-Pagan Lagrange multiplier test. All models were run with standard errors clustered at the country level, given significant results (p<0.05) from the Wald test for groupwise heteroskedasticity and Lagrange-Multiplier test for serial correlation.

Regarding economic factors, the log of income per capita (GDP) is included in the model to account for a country’s state of economic development. A higher GDP has been associated with higher primary care utilization, including SRH services as well as maternal health outcomes (Arsenault, Kim et al. 2020). The model also includes public domestic health expenditure as a share of current health expenditure (GHE/CHE), which captures a government’s investment in healthcare and citizens’ human capital. Based on existing evidence, a domestic healthcare financing structure with a large share of public spending (relative to private and out of pocket expenditure) is expected to be favorable for SRH service provision (Novignon, Olakojo et al. 2012; Ravindran and Govender 2020).

Next, the model includes indicators of corruption and government effectiveness indexes from the World Governance Indicators (WGI) database to account for the influence of governance quality on health services delivery. Good governance may promote service access through stronger administrative institutions, better policies, and improved resource utilization and allocation (Makuta and O’Hare 2015; Lee, Yang et al. 2016).

Primary school enrolment (gross shares) and female labour market participation rate (among women above the age of 15) are added to control for population education level and female empowerment. Across the world, there is evidence that empowerment through education, labor, and political participation are enabling factors for healthcare utilization, including SRH services (Arsenault, Kim et al. 2020; Khatiwada, Muzembo et al. 2020; Dickson, Adde et al. 2021).

Finally, the model accounts for population density (number of people per square kilometer of land) as higher density may improve service access by generating economies of scale in healthcare provision. Higher share of urban population and national population density have been shown to be positively related to accessing maternal health coverage, including antenatal care visits and skilled birth attendance (Hanlon, Burstein et al. 2012). Our main specification measures ODA in per capita terms (constant 2020 USD) to determine the effect of ODA after accounting for countries’ population sizes. Technically, the model is based on a linear function, which provides estimates of the absolute change (in percentage points) in SRH service coverage associated with a USD per capita change in health ODA disbursement.

### 3.4 Sensitivity analysis

To assess the robustness of the results, we perform a series of sensitivity analyses with respect to missing values, measurement error, potential outliers, model specification, and reverse causality. First, to compensate for the missing values of certain observations, we impute values using interpolation (i.e., inserting values based on the values before and after the missing values). Using imputed values in the estimation of the main models did not affect the main results of the analysis. Additionally, to address the risk of measurement error, we averaged each variable over three-year periods giving a panel dataset of 119 countries measured over six 3-year periods.^4^ This reduces the variation in the yearly time-series data to provide more stable observations. Overall, the use of the three-year average values along with the other sensitivity analyses resulted in estimates that were in line with the main results of the analysis.

Second, the potential problem of a strong influence of outliers (observations with values very far from the average values) was investigated by using plots to identify such observations and calculating Cook’s Distance statistics. The main models were estimated after removing Cook’s Distance outliers and two additional outlier robust FE estimators were employed (the S-estimator and the MM-estimator). Adjusting the main models for some of the potential outliers affected the coefficient estimates for some of the variables but did not affect the main results of the analysis.

Third, to assess the sensitivity of the main models to the functional form of the main measures, we re-estimated the models in the log-log specification. We used the logarithmic values of all variables, including development assistance measures (in millions USD). This log-log form measures elasticities i.e., the percent change in SRH service outcomes associated with a one percent change in SRH-ODA disbursements. The use of the alternative functional form variables affected the statistical significance of some of the effects of ODA on these outcomes but did not alter the main results more generally.

Fourth, it is possible that the effects of ODA are not contemporaneous such that ODA allocated in one year only affects the outcomes in the following years. We adjusted our main models to investigate such a delayed effect by using lagged values of health ODA (ODA_t-1_) and other independent variables (X_t-1_) as regressors for current service coverage. The results of this adjustment suggest that ODA may indeed have delayed effects on these outcomes in the sense that our main results were somewhat strengthened by the adjustment.

Finally, to account for a potential dynamic relationship between ODA and SRH services, we implemented a two-step system-general method of moments (GMM) estimation, characterized by the inclusion of the lagged dependent variable (service coverage) among the regressors. Next to the maximum likelihood estimation and structural equation modelling (ML-SEM) approach (Moral-Benito, Allison et al. 2019), GMM estimators are most commonly used to estimate dynamic panel models (Baltagi 2021). Our main findings were broadly robust to all of these alternative estimation approaches.

## 4 Results

This section presents the results of the econometric analysis. We first present the main results and then describe additional results of relevance. We then present the results of the sensitivity analysis. Detailed results are presented in Appendix C. Main results and in Appendix D. Complementary Results. The results of the sensitivity analysis are presented in Appendix E.

### 4.1 Effects of ODA on SRH services

The main finding of our analysis is that development assistance for health and for SRH has had, on average, positive effects on service coverage during the period 2002-2020 in our sample of countries (Table 3). Total health ODA has a positive effect on two out of three outcomes. A 1 USD per capita increase in SRH-ODA is, on average, associated with a 0.628 percentage point increase in contraceptive prevalence, a 0.190 percentage point increase in skilled birth attendance and a 0.531 percentage point increase in ART coverage. With respect to RH-ODA, the results suggest that also this type of ODA was, on average, positively associated with service coverage although the results were not statistically significant.

**Table 3:** Effects of ODA per capita on SRH services, 2002-2020.

| VARIABLES | SRH-ODA | Total health ODA | RH-ODA |
| --- | --- | --- | --- |
| **Panel A) Access to modern contraceptives** |  |  |  |
| Health ODA per capita | 0.628*** | 0.242** | 0.935 |
|  | [0.330 - 0.926] | [0.032 - 0.453] | [-1.490 - 3.361] |
| **Panel B) Skilled birth attendant (SBA)** |  |  |  |
| Health ODA per capita | 0.190* | 0.059 | 0.902 |
|  | [-0.009 - 0.389] | [-0.028 - 0.146] | [-1.111 - 2.916] |
| **Panel C) Anti-retroviral therapy (ART)** |  |  |  |
| Health ODA per capita | 0.531*** | 0.332*** | 0.575 |
|  | [0.380 - 0.682] | [0.193 - 0.471] | [-1.081 - 2.231] |

Note. Standard errors are clustered at the country level. \*\*\* $p < 0.01$ , \*\* $p < 0.05$ , \* $p < 0.1$ , and 95 % CI in brackets. Health ODA per capita is a placeholder for the type of ODA used in the model as indicated in the column headings.
Source: OECD-CRS, WDI, WGI.

More specifically, the analysis shows that Total health ODA and SRH-ODA had statistically significant positive effects on modern contraceptive prevalence over the sampling period. Looking at SRH-ODA, in our sample of LLMICs, a 1 USD *per capita* increase in SRH-ODA (e.g., an increase from 3 to 4 USD per capita) is, on average, associated with a 0.628 percentage point increase in the contraceptive prevalence rate (e.g., an increase from 20.0 percent to 20.628 percent). This suggests that to increase contraceptive prevalence rate from the average for LICs 2002-2020, which was 30.55 percent, to 35 percent, SRH-ODA would have to increase from, on average 3.85 USD/capita to 10.40 USD/capita, (i.e., an increase of SRH-ODA of 6.54 USD/capita or approximately 170 percent).

The findings suggest that ODA has had a small positive contribution to skilled birth attendance in the sample of countries during this period. However, apart from SRH-ODA, the positive effects are not statistically significant. Instead, government health expenditure as a share of current health expenditure and population density both had statistically significant positive effects on SBA.

Regarding treatment for HIV, our estimations show that ODA has, on average, had a positive effect on the coverage of ART in the studied countries. More specifically, Total health ODA and SRH-ODA had a statistically significant positive effect on ART service coverage whereas RH-ODA had a positive but not statistically significant effect. A 1 USD increase of SRH-ODA is, on average, associated with a 0.531 percentage point increase in antiretroviral therapy coverage (e.g., from 20 percent to 20.531 percent). For example, increasing SRH-ODA from 3 to 4 USD per would be associated with an average 0.531 percentage point increase in ART coverage. This suggests that to increase ART coverage from the current LIC average of 80 percent to the UNAIDS target of 90 percent would require an increase of SRH-ODA of 18.83 USD/Capita (i.e., an increase of nearly 500 percent from the 2002-2020 average SRH-ODA).

### 4.2 Effects of ODA by income group

Income has generally been an important factor for the SRH outcomes. In particular, GDP per capita has had a strong effect on skilled birth attendance in the countries over the analysed period. In addition, the observed effects of ODA generally appear to be stronger in low-income countries than in lower-middle-income countries (Figure 4).

**Figure 4:**
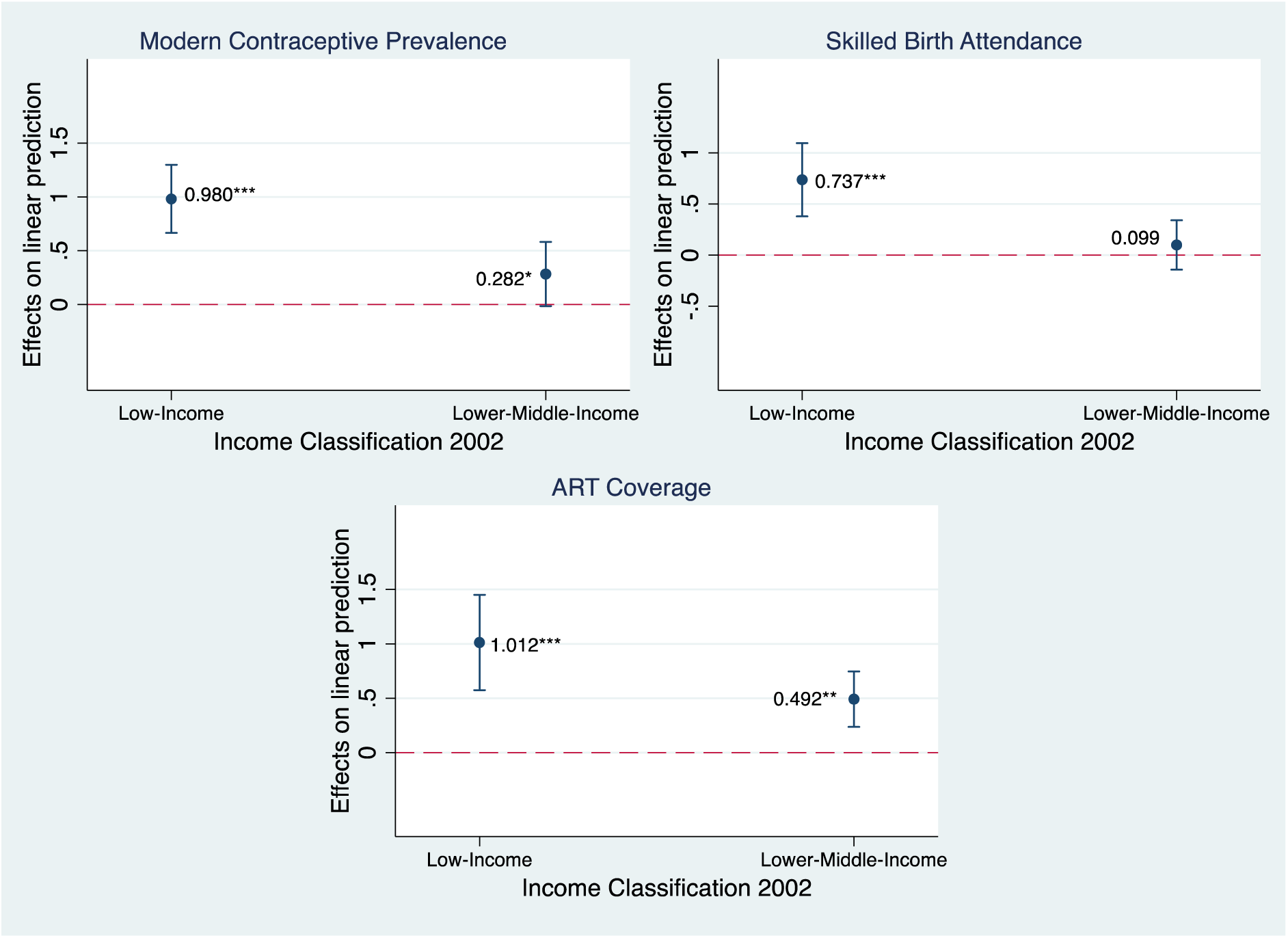
Marginal effect of SRH-ODA (2020 USD, pc) on SRH service outcomes, by countries income classification in 2002 (based on 3-Year-Averages Models)

### 4.3 The effects of ODA by volume, time, and type

While the main findings of the analysis suggest that, in general, ODA to SRH-services has had a positive effect over this period, there may be reasons to investigate whether the effect has varied by volume, across time, and by type of donor. The issue of a possibly different effect of ODA depending on the volume of aid is important as it may suggest that the amount of ODA allocated to a country needs to reach a certain level before it begins to affect the outcome in any meaningful way. The question of whether the effect of ODA is stronger during the later years of the study period is important as it captures potential improvements in development partners’ and recipient countries’ effective use of ODA for the intended purposes. And finally, understanding if ODA channeled by different donors may have policy implications for the allocation of SRH-ODA.

We find that SRH-ODA may have a “threshold effect” in that SRH-ODA may need to reach a certain volume relative to domestic government health expenditure to have a significant positive contribution to SRH service outcomes (Figure 5). However, we also find that the marginal effect of ODA plateaus above certain levels (roughly when SRH-ODA exceeds 25 percent of government health expenditure). This finding is important as it suggests that development partners should find an appropriate balance between providing sufficient levels of ODA to, on one hand, reach the threshold level and, on the other, not provide excessive levels of ODA that the recipient country is unable to utilize effectively.

**Figure 5:**
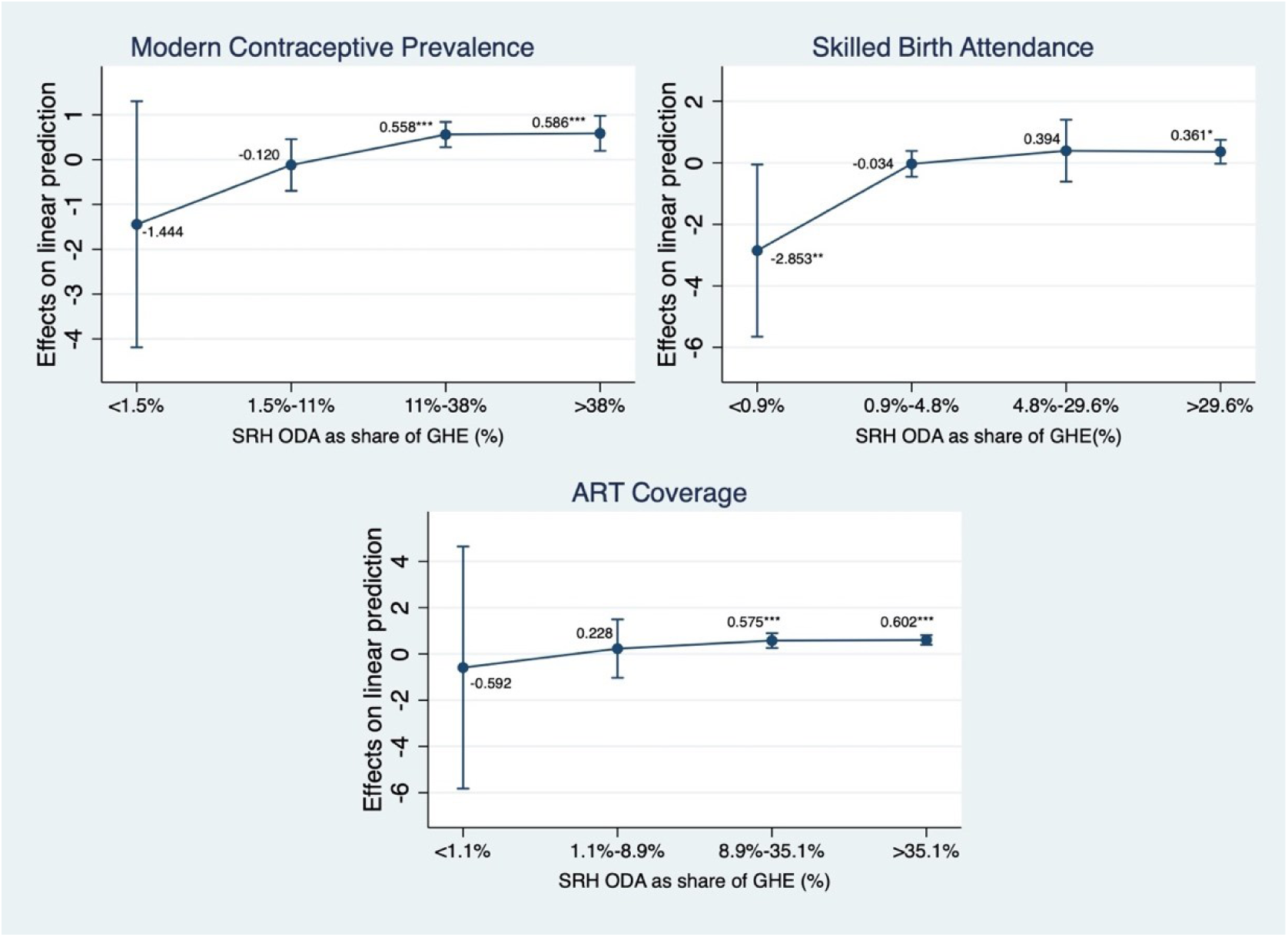
Marginal (threshold) effect of SRH-ODA (2020 USD, pc) on SRH services, by ODA relative to government health expenditure (based on 3-Year-Average Models)

Similarly, we investigated whether there is a difference in the effect of SRH-ODA depending on the volume of SRH-ODA relative to the total ODA received by a country. The findings suggest that the positive contribution of SRH-ODA is most apparent when SRH-ODA represents a larger share of total ODA (Figure 6).

**Figure 6:**
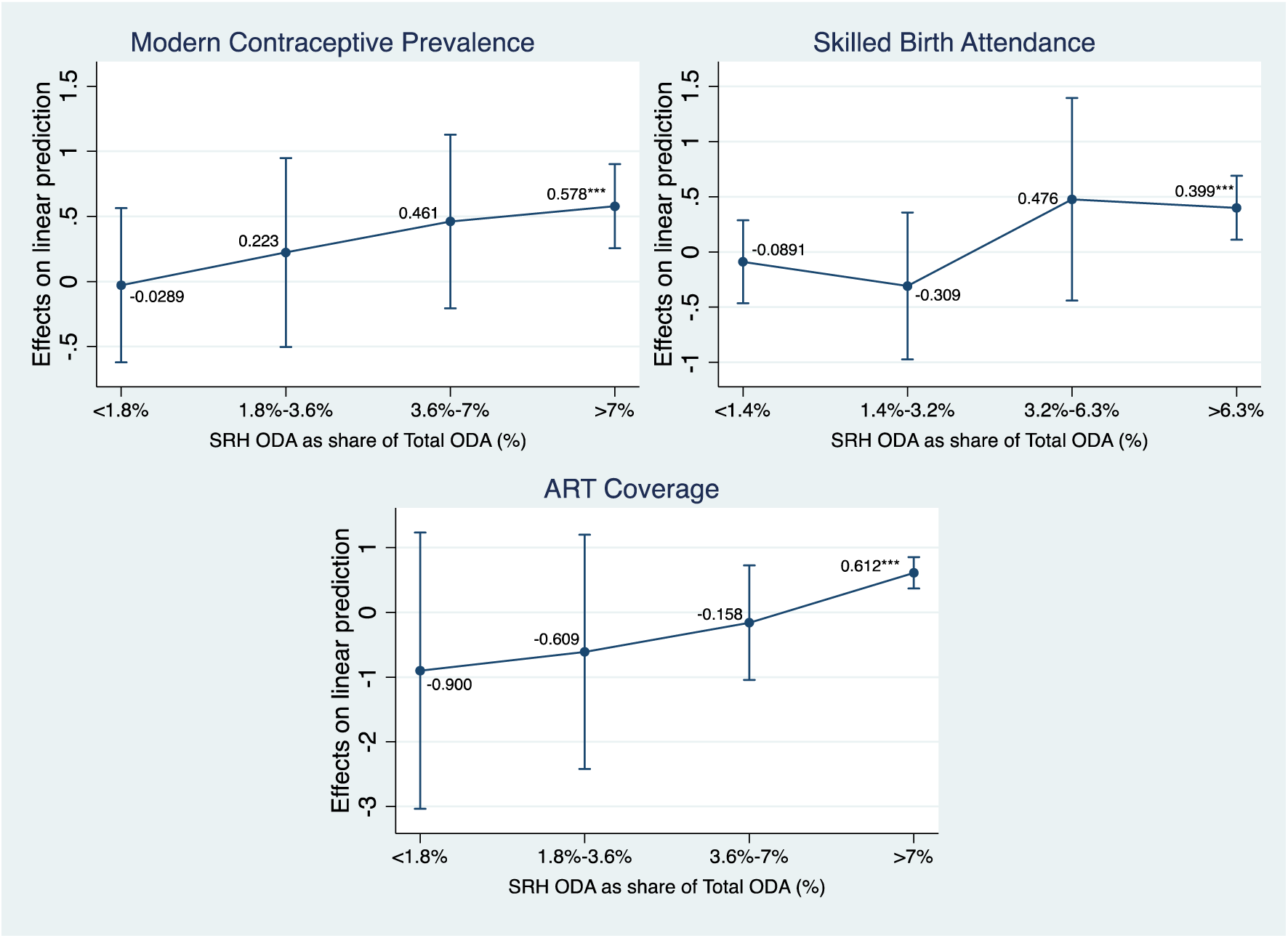
Marginal effect of SRH-ODA (2020 USD, pc) on SRH services, by SRH-ODA relative to Total ODA (based on 3-Year-Average Models)

We also find evidence suggesting that the effects of SRH-ODA have increased over time (Figure 7). The finding of a positive learning effect is also important as it provides support to the continued allocation of ODA to support the provision of SRH services, especially in low-income countries.

**Figure 7:**
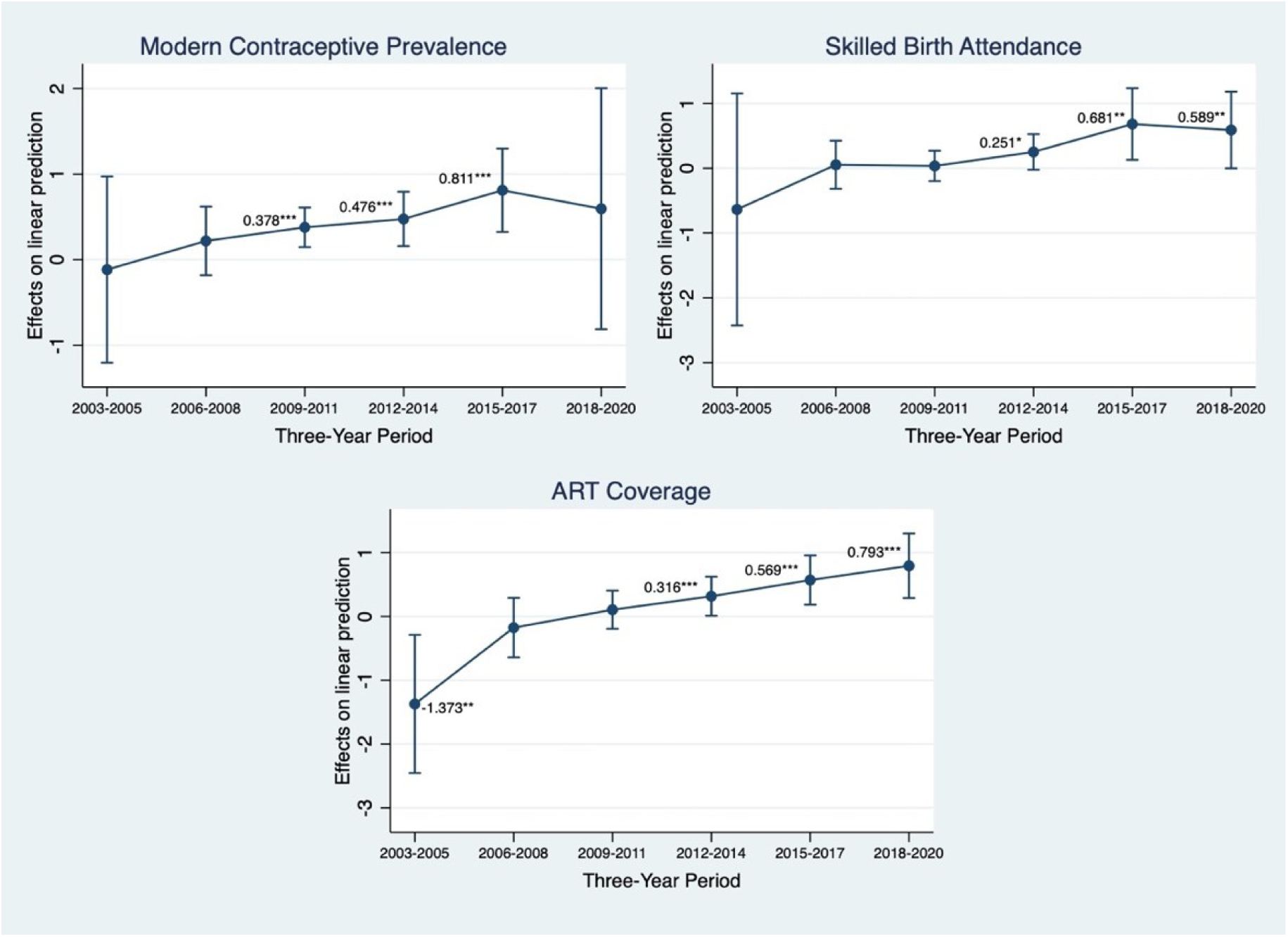
Marginal effect of SRH-ODA (2020 USD, pc) on SRH service outcomes, by 3-Year period (based on 3-Year Averages Models)

Finally, we find some support for the proposition that the effect of ODA varies by donor type. Bilateral ODA appears to be more effective than multilateral development assistance (Figure 8).

**Figure 8:**
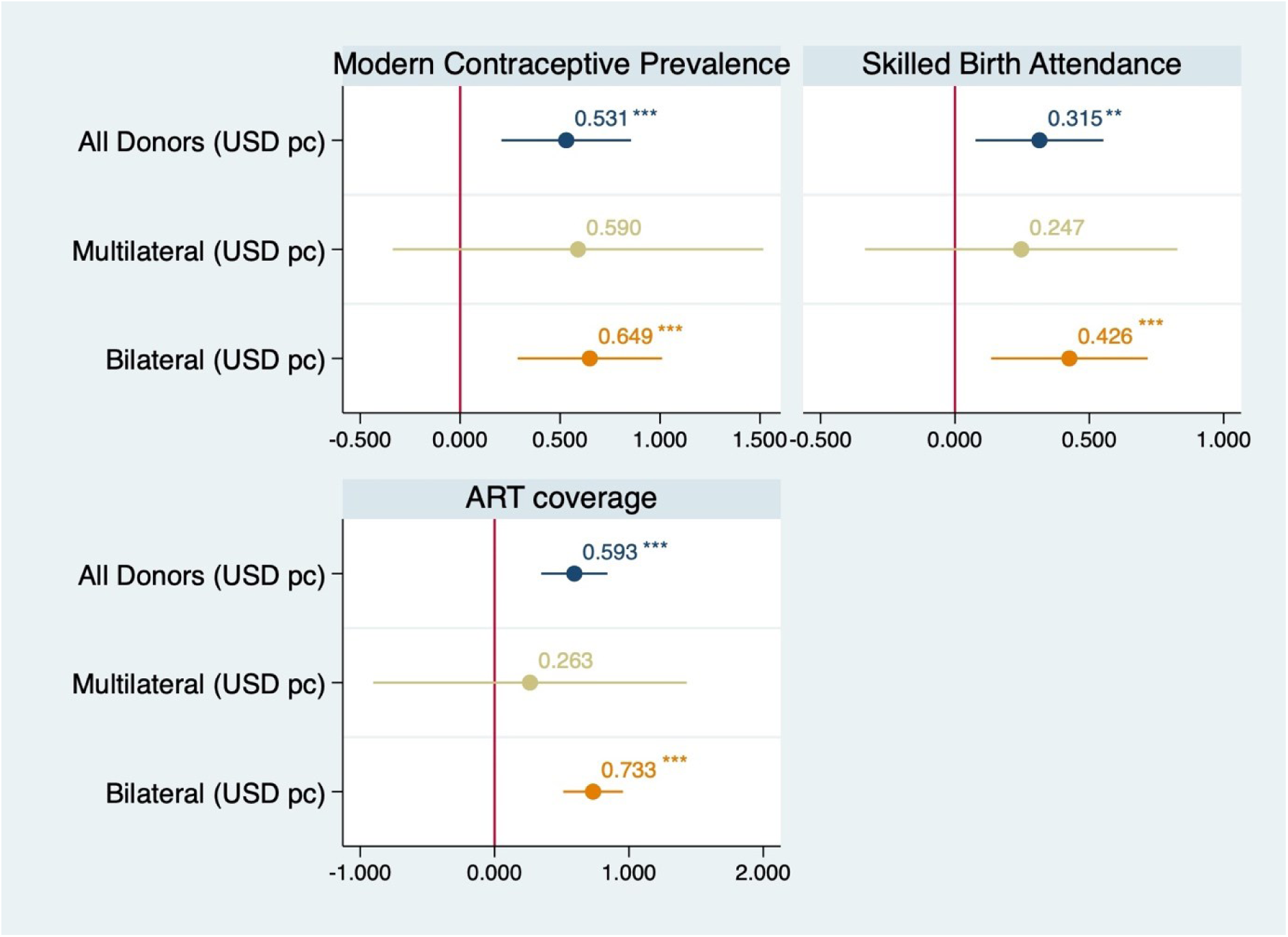
Effects of SRH-ODA (2020 USD, pc) on SRH service outcomes, by donor type (based on 3-Year Averages Models)

### 4.4 Sensitivity analyses

To assess the robustness of the main results, we conducted a series of sensitivity analyses as described in sub-section 3.4. First, we investigated potential bias from measurement error and missing data by using interpolation and 3-year average values to fit the models. Second, we re-fitted the models after removing observations that may be defined as outliers, including by Cook’s Distance tests. Third, we estimate the main models using alternative specifications, including linear-log and log-log transformations. Fourth, we investigate the potential of a delayed impact of ODA on service outcomes with a lagged independent variables (IVs) model. And finally, we implemented a dynamic FE estimator, the system-GMM estimator, to account for the potential endogeneity of the regressors. The results of these analyses are presented in Appendix E. Sensitivity analysis.

## 5 Discussion and conclusions

The findings of this study suggest that, on average, ODA has had a positive effect on skilled birth attendance, access to contraceptives, and coverage of anti-retroviral therapies in low- and lower-middle-income countries during the period 2002-2020. While the effects are small and vary across these outcomes, the findings provide general support the continued allocation of development assistance to improve SRHR service outcomes in low- and lower-middle-income countries. However, further analyses of the data have revealed that the effects of ODA are not uniform across different levels of ODA, time periods, and types of ODA. In this section, we critically discuss these findings and point out remaining gaps in the evidence base. Based on the discussion, we draw conclusions of relevance to research, practice, and policy development.

### 5.1 Discussion

The main findings of the study are broadly in line with the findings of previous studies in this field. However, they also indicate that the effects of aid may be more complex than would initially appear to be the case. The magnitude of the effect seems to be stronger for SRH-specific ODA than for Total health ODA, which is reasonable given the narrower focus of SRH-ODA. Our findings are thus in line with similar studies that have identified a positive effect of SRH-ODA on SRH-related mortality (Wilson 2011; Taylor, Hayman et al. 2013). While most studies in our review focused on the effects of aid on long-term outcomes (such as maternal or infant mortality), a few studies have examined the effect of ODA on service indicators. In line with our findings, Bendavid et al. (Bendavid, Leroux et al. 2010) observed a positive correlation between ODA and ART coverage, although their study was limited to the effect of ODA on HIV specifically. Shepard et al. (Shepard, Bail et al. 2003) estimated the effect of USAID family planning funding during the period 1989-1998 on the contraceptive prevalence rate and found a significant positive association. This is in line with our findings, though their analysis was much more limited in terms of the sources of ODA included and covered a different time period. Grepin (Grépin 2012) studied how different types of health ODA correlated with maternal health service provision but found no significant associations for any of the types of health ODA. Our analysis reported similar results where we noted a positive, though insignificant, effect of health ODA on deliveries attended by a skilled birth attendant. The findings also suggest that other factors may have played a more significant role than health ODA in strengthening these services over the past two decades. In particular, economic development and population density seem to be of importance, as does control of corruption for some outcomes. Our findings thereby resonate with Lee, Yang and Kang (Lee, Yang et al. 2016) who argue that ODA needs to be accompanied by efforts to improve governance and accountability in order to be effective. Toseef et.al made similar claims in a more recent study (Toseef, Jensen et al. 2019).

In this study, we measured the effects on service indicators using three measures of ODA (Total health ODA; SRH-ODA and RH-ODA). The strongest and most significant effect was observed for SRH-ODA. Even though we did not expect a strong effect of reproductive health ODA on contraceptive prevalence and ART coverage, as funding to these areas should be reported in other ODA sector codes, the relatively weak association between reproductive health ODA and the skilled birth attendance indicator is noteworthy. One possible explanation for the lack of a strong association could be that the share of births attended by skilled personnel was at a relatively high level already in 2002 – around 70 percent on average in the sample of LLMICs – compared to the contraceptive prevalence rate and ART coverage, which were around 36 percent and 1 percent, respectively. This may point to the challenge of reaching universal coverage of SRH services as more resources may be needed to reach the remaining uncovered population groups to make up for the possible decreasing marginal returns to aid.

Given the recognition of the strong link between SRHR and general development, the importance of addressing SRHR is widely acknowledged today. It is therefore important to understand the overall contribution of SRH-ODA to the improvement of key SRH indicators. This study contributes to a better understanding of the overall effects of SRH-ODA on specific sexual and reproductive health indicators at the macro level using quantitative data. That said, other types of studies and additional evidence are needed to broaden the evidence base. Rather than more studies of the same kind, we argue for the value of different types of analyses, including qualitative and quantitative (as well as mixed methods approaches) studies at both macro and micro levels. Quantitative studies using micro-level data could provide evidence of the effects of SRH-ODA at the level of households and communities. Qualitative studies could explore how ODA contributes to policy development for advancing SRH or how issues of SRH-ODA are perceived and experienced by the beneficiaries. Combined, such a “patchwork” of evidence would provide a more comprehensive answer to the question of how SRH-ODA can be most effectively implemented.

Our analysis does not capture how large the share of SRH interventions funded by SRH-ODA is in each country. We know that in several countries, the majority of SRH services are funded with the support of ODA (The Partnership for Maternal 2019). When a large share of a country’s health sector or general government expenditure is financed by ODA, it is often referred to as “aid dependent” (Marty, Dolan et al. 2017; Olakunde, Adeyinka et al. 2019). Therefore, the share of SRH interventions funded by ODA in a country is one important factor to consider in order to understand the effect of ODA. For example, the HIV/AIDS response in Malawi is 90 percent funded using external resources,^5^ primarily ODA. In such a funding scenario, improvements in ART coverage will likely be largely attributable to SRH-ODA. However, one reason for countries like Malawi being so heavily reliant on ODA to finance the HIV/AIDS response is that development partners have earmarked large funds for this specific purpose. At the overall health sector level, however, reliance on external funding is much lower (although still high in absolute terms), about 50 percent of current health expenditure in Malawi. Consequently, the reliance on ODA appears higher when looking at certain sub-sectors, such as HIV/AIDS. This “earmarking” of resources for specific purposes is one reason for why we conducted our analysis based on several measurements of ODA and why we opted to control for the level of government spending on health. Our results appear robust with respect to both concerns, but earmarking could explain the slightly lower effect size that we observe when analyzing Total health ODA compared to the sub-category SRH-ODA.

Another reason why we included several measures of ODA in our analyses is that aid targeting SRHR is not always reported as SRH-ODA. Other studies trying to measure SRH-ODA have illustrated the difficulties of relying solely on SRH-ODA codes, as many SRHR interventions are reported using other statistical codes (Schäferhoff, Van Hoog et al. ; DSW and European Parliamentary Forum 2020). For example, programs targeting violence against women or supporting LGBTQI rights are commonly reported under sector codes outside the health sector. So, despite our expanded measure of health ODA, we acknowledge that even this measurement does not fully capture all ODA of relevance for SRHR, implying that we might have underestimated the effect of ODA on SRH outcomes.

Our analysis also provides further results of relevance. First, we find support for the existence of threshold effects of health ODA. In particular, health ODA appears to have an effect once it reaches a certain share of available domestic government health spending (measured as government health expenditure, GHE). While threshold effects have been explored in other studies of ODA (Wayoro and Ndikumana 2020), an exploration of the threshold effect of health or SRH-ODA as share of GHE was not found in any of the studies we have reviewed. There may be several reasons for the threshold effect we observed. One reason could simply be that below a certain level, health ODA does not have any material effect on available resources or on policy effectiveness. Similarly, additional analyses show that the effect of ODA appears to be stagnating as it expands beyond a certain share of domestic resources. As shown above, for modern contraceptive prevalence, we find a threshold at around 39 percent of total government health expenditures, where the effect of health ODA is less pronounced compared with when it constitutes between 11 and 38 percent. Results look similar for both skilled birth attendance and ART coverage, but at slightly different shares of health expenditure (see Figure 5 for details).

There may also be several different reasons for an upper threshold for the effect of health ODA on these outcomes and further investigation is needed to draw any conclusions. However, one reason could be that if health ODA makes up a very large share of total available financial resources, it may lead to a situation where too many programs, projects, and services are being implemented without a sufficient supply of other resources, such as personnel or medicines. It is broadly accepted that the effective delivery of SRH services (and other types of health care and other types of services) ultimately depends on the availability of competent and motivated staff. It may thus be the case that the effect of health ODA directed toward these services is reduced due to the lack of such staff. An additional and related reason may be that large amounts of ODA lead to coordination problems at the management level of the health system. The inability to effectively coordinate the allocation of resources may lead to diminished effectiveness (Sundewall and Sahlin-Andersson 2006; Chansa, Sundewall et al. 2008).

The second result of further relevance that we report is that the effects of health ODA on these service outcomes appear to have increased over the course of the study period. As shown in Figure 7, the general trend is that the effects of health ODA are larger toward the end of the period compared with the beginning of the period. The reasons for such an effect may, among others, be due to a learning effect, whereby countries and development partners become better at utilizing health ODA effectively. For example, overall management, planning, and coordination of resources may have become more well-functioning over time as those involved in managing health ODA and domestic resources have become more skilled at it. It may also be due to the effectiveness of the various agreements, such as the SDGs and the Paris Declaration on Aid Effectiveness, that have been reached over the past decades or so about the focus areas for ODA, and criteria and conditions for ODA allocation and disbursement. Pallas and Ruger (Pallas and Ruger 2017) argued that it takes time for commitments to translate into disbursements and it takes time for disbursements to generate outcomes, which could also contribute to, although not fully explain, the stronger effect towards the end of the time period.

Third, and finally, we also find that the source of ODA appears to matter. Specifically, we find that ODA, where the source is classified as ‘bilateral’, is associated with a larger effect compared with ODA classified as ‘multilateral’. It should be reiterated that this analysis is limited to ODA by *source of funds* (i.e., where funding is coming from), and not by, for example, *channel* (i.e., who implements the development program). Biscaye et.al. (Biscaye, Reynolds et al. 2017) conducted a review of 45 articles that all tested the association between bilateral and multilateral aid flows and did not find any consistent evidence of one being more effective than the other. That said, while different types of ODA play different roles, our finding that the effects of ODA may vary by source is noteworthy on a principal level as both sources of ODA most likely will continue to be important over the coming years. While these results are relevant to the understanding of the role of development assistance, the validity of the results will need to be confirmed by further studies using supplementary data and methods.

### 5.2 Conclusions

Our study suggests that overall SRH-ODA has a positive effect on ART coverage, the contraceptive prevalence rate and the share of births attended by skilled personnel. This is encouraging and at an overall level, our results lend support to continued investments in SRHR. However, the effects of ODA on these services that we report are relatively small and even if SRH-ODA were substantially increased it would not be sufficient to reach targets set for the outcomes we have studies. For example, our results suggest that to reach 90 percent coverage of ART, SRH-ODA would have to increase by almost 500 percent, which is not realistic to expect. This suggests that reaching the target of 90 percent coverage of ART, will likely not be achieved only through increased SRH-ODA. Development partners would be therefore advised to also continue improving the effectiveness of SRH-ODA. Furthermore, our analysis indicates that there may exist a threshold effect of ODA, which leads us to argue that investments in SRHR must be large relative to the share of domestic funding for health. This would imply that it is better to focus SRH-ODA on fewer and larger programs in a limited number of countries rather than spreading resources between many countries. As we have argued in this report, effective development assistance requires different types of evidence bases. This study has contributed to an improved understanding of the role of ODA in this area, but it is only one piece of the evidence required. Successful implementation of SRHR programs projects will require multiple sources of evidence from research applying, respectively, both qualitative, quantitative, and mixed-methods approaches.

## Data Availability

All data used in the study are open available from the sources provided in the text.

OECD/EC-JRC. Access and Cost of Education and Health Services: Preparing Regions for Demographic [Internet]. Paris: OECD Publishing; 2021 [cited 2022 April 5]. Available from: https://www.oecd.org/publications/access-and-cost-of-education-and-health-services-4ab69cf3-en.htm.

## Appendices

## Appendix A. Data Collection and Sample Construction

**Table A1.** Sample of 103 ODA-eligible countries classified as LLMIC in 2002.

| **Country** | **id** | **IncomeGroup2002** | **IncomeGroup2022** |
| --- | --- | --- | --- |
| Afghanistan | 1 | L | L |
| Albania | 2 | LM | UM |
| Algeria | 3 | LM | LM |
| Angola | 4 | L | LM |
| Armenia | 5 | LM | UM |
| Azerbaijan | 6 | L | UM |
| Bangladesh | 7 | L | LM |
| Belarus | 8 | LM | UM |
| Benin | 9 | L | LM |
| Bhutan | 10 | L | LM |
| Bolivia | 11 | LM | LM |
| Brazil | 12 | LM | UM |
| Burkina Faso | 13 | L | L |
| Burundi | 14 | L | L |
| Cabo Verde | 15 | LM | LM |
| Cambodia | 16 | L | LM |
| Cameroon | 17 | L | LM |
| Central African Republic | 18 | L | L |
| Chad | 19 | L | L |
| China | 20 | LM | UM |
| Colombia | 21 | LM | UM |
| Comoros | 22 | L | LM |
| Congo, Dem. Rep. | 23 | L | L |
| Congo, Rep. | 24 | L | LM |
| Cote d'Ivoire | 25 | L | LM |
| Dominican Republic | 26 | LM | UM |
| Ecuador | 27 | LM | UM |
| Egypt | 28 | LM | LM |
| El Salvador | 29 | LM | LM |
| Equatorial Guinea | 30 | L | UM |
| Eritrea | 31 | L | L |
| Eswatini | 32 | LM | LM |
| Ethiopia | 33 | L | L |
| Fiji | 34 | LM | UM |
| Gambia | 35 | L | L |
| Georgia | 36 | L | UM |
| Ghana | 37 | L | LM |
| Guatemala | 38 | LM | UM |
| Guinea | 39 | L | L |
| Guinea-Bissau | 40 | L | L |
| Guyana | 41 | LM | UM |
| Honduras | 42 | LM | LM |
| India | 43 | L | LM |
| Indonesia | 44 | L | LM |
| Iran | 45 | LM | LM |
| Iraq | 46 | LM | UM |
| Jamaica | 47 | LM | UM |
| Jordan | 48 | LM | UM |
| Kazakhstan | 49 | LM | UM |
| Kenya | 50 | L | LM |
| Kyrgyzstan | 51 | L | LM |
| Lao PDR | 52 | L | LM |
| Lesotho | 53 | L | LM |
| Liberia | 54 | L | L |
| Madagascar | 55 | L | L |
| Malawi | 56 | L | L |
| Maldives | 57 | LM | UM |
| Mali | 58 | L | L |
| Mauritania | 59 | L | LM |
| Moldova | 60 | L | UM |
| Mongolia | 61 | L | LM |
| Morocco | 62 | LM | LM |
| Mozambique | 63 | L | L |
| Myanmar | 64 | L | LM |
| Namibia | 65 | LM | UM |
| Nepal | 66 | L | LM |
| Nicaragua | 67 | L | LM |
| Niger | 68 | L | L |
| Nigeria | 69 | L | LM |
| North Macedonia | 70 | LM | UM |
| Pakistan | 71 | L | LM |
| Papua New Guinea | 72 | L | LM |
| Paraguay | 73 | LM | UM |
| Peru | 74 | LM | UM |
| Philippines | 75 | LM | LM |
| Rwanda | 76 | L | L |
| Samoa | 77 | LM | LM |
| Sao Tome and Principe | 78 | L | LM |
| Senegal | 79 | L | LM |
| Serbia | 80 | LM | UM |
| Sierra Leone | 81 | L | L |
| Solomon Islands | 82 | L | LM |
| South Africa | 83 | LM | UM |
| Sri Lanka | 84 | LM | LM |
| St. Vincent and the Grenadines | 85 | LM | UM |
| Sudan | 86 | L | L |
| Suriname | 87 | LM | UM |
| Tajikistan | 88 | L | LM |
| Tanzania | 89 | L | LM |
| Thailand | 90 | LM | UM |
| Timor-Leste | 91 | L | LM |
| Togo | 92 | L | L |
| Tonga | 93 | LM | UM |
| Tunisia | 94 | LM | LM |
| Turkey | 95 | LM | UM |
| Uganda | 96 | L | L |
| Ukraine | 97 | LM | LM |
| Uzbekistan | 98 | L | LM |
| Vanuatu | 99 | LM | LM |
| Vietnam | 100 | L | LM |
| Yemen | 101 | L | L |
| Zambia | 102 | L | LM |
| Zimbabwe | 103 | L | LM |

**Note.** Data on Income Group classification from the World Bank. **Abbreviations.** L, Low-income countries; LM, Lower-middle income countries.

**Table A2.** List of excluded countries with reasons for exclusion.

| Primary exclusion reason | Countries | Country characteristics |
| --- | --- | --- |
| Lacking information on service coverage indicators | Anguilla | SIDS |
|  | Cook Islands | SIDS |
|  | Kosovo | Post-conflict fragility |
|  | Tokelau | SIDS |
|  | Tuvalu | SIDS, LDC |
|  | Wallis and Futuna | SIDS |
|  | Niue | SIDS |
|  | Nauru | SIDS |
| Lacking info on GDP pc (PPP) | Cuba | SIDS |
|  | Djibouti | Fragile context |
|  | Syria | Extremely fragile context |
| Lacking info on primary school enrolment (% gross) | Haiti | SIDS, LDC |
|  | Turkmenistan | UMIC |
|  | Bosnia | UMIC |
| Lacking info on female labour market participation | Kiribati | SIDS, LDC |
|  | Korea | HIC |
|  | Micronesia | SIDS |
|  | Marshall Island | SIDS |
| Lacking info on government health expenditure (% country expenditure) | West Bank and Gaza | Fragile context |
|  | Montenegro | UMIC |
|  | South Sudan | Extremely fragile context |
|  | Somalia | Extremely fragile context |

**Note.** Table 1 provides a list of excluded low-and-lower-middle-income countries with reasons for exclusion for transparency. Country characteristics were assessed to see whether excluded countries significantly differed from included countries. **Abbreviations.** GDP, gross domestic product; HIC, high-income country; LDC, least developed country; SIDS, small island developing state, UMIC; upper-middle-income country. Information on country characteristics from the United Nations list of SIDS and OECD “States of fragility 2020” report.

**Table A3.**
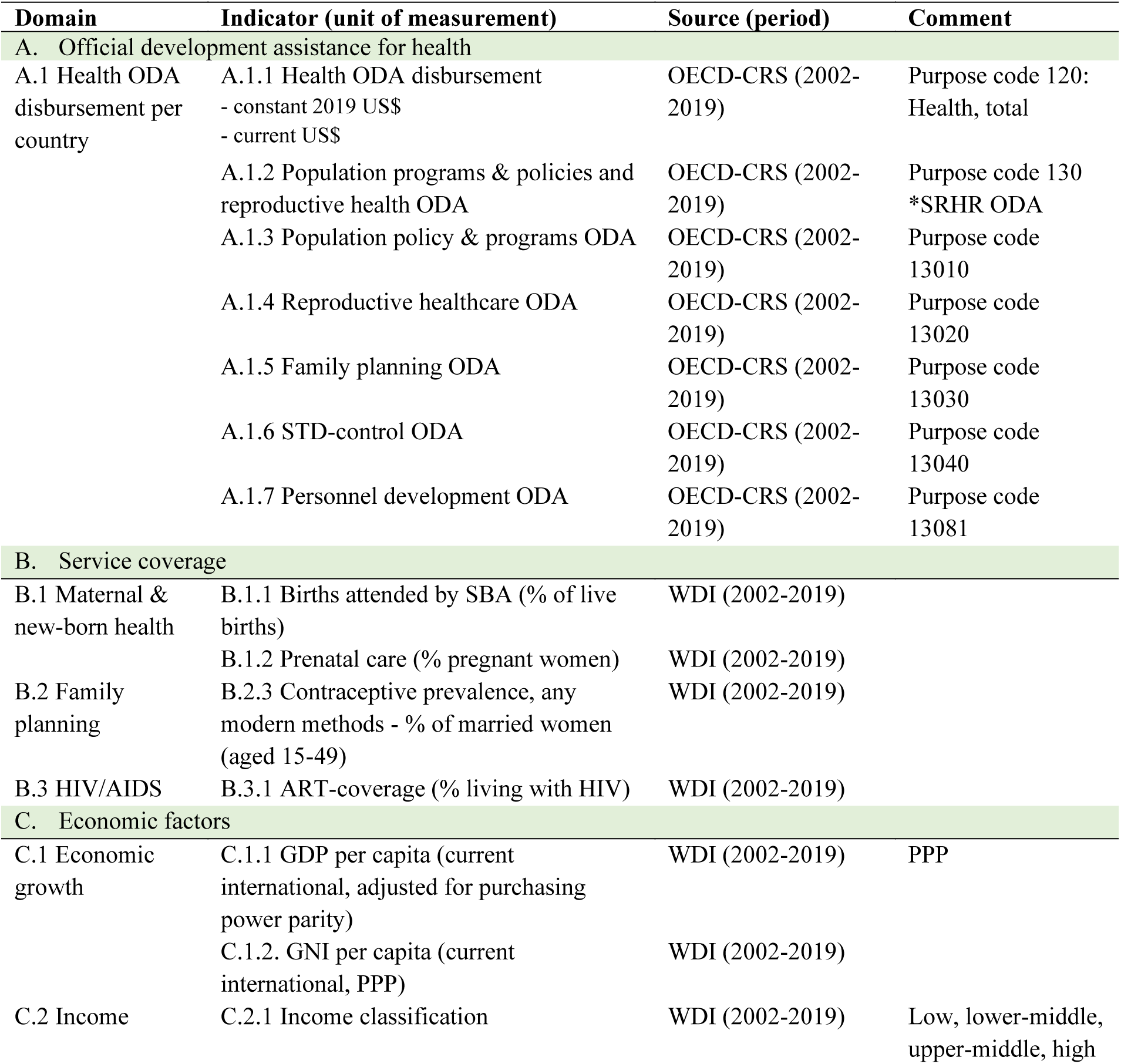
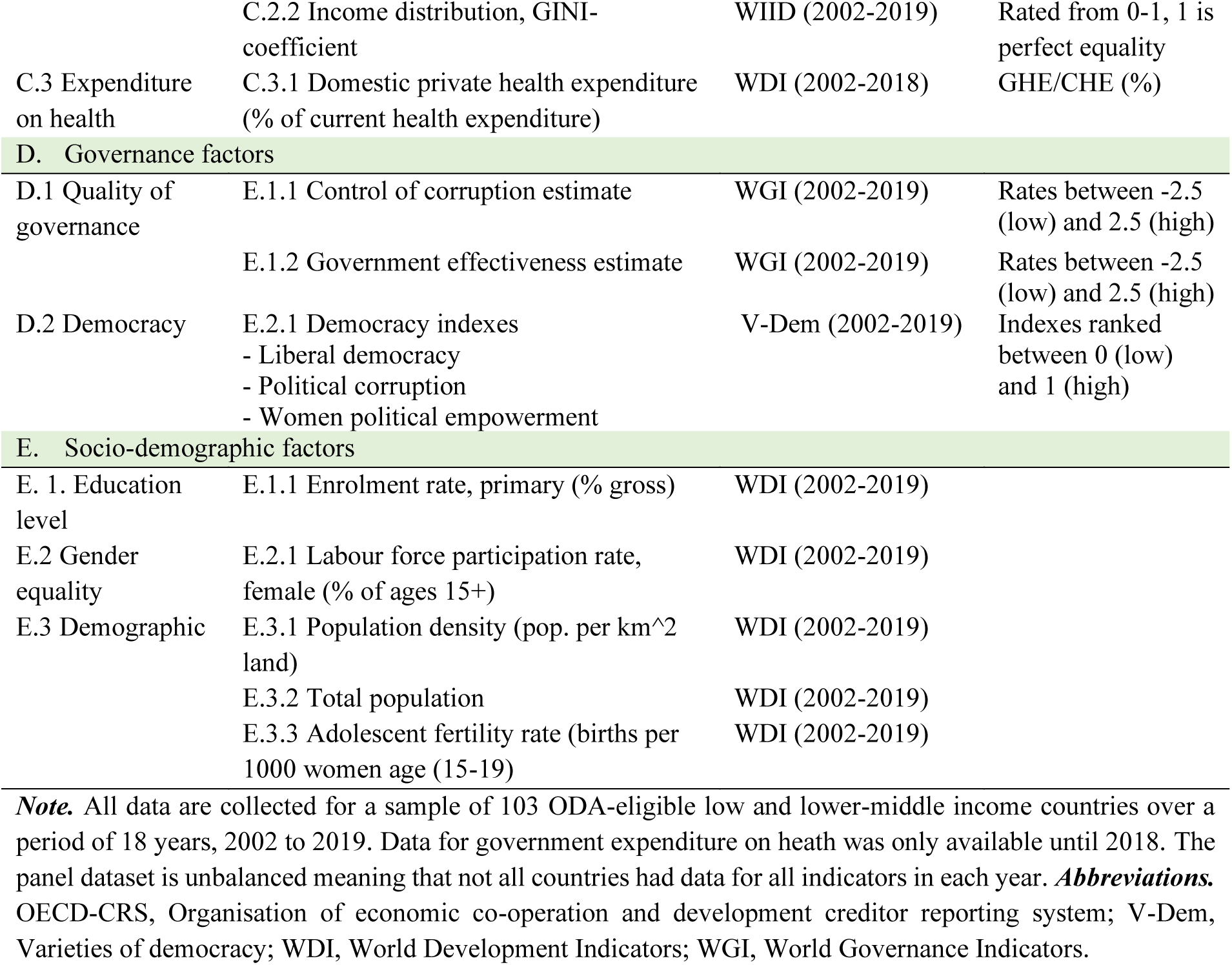
Collected indicators including sources of data.

**Figure A1.**
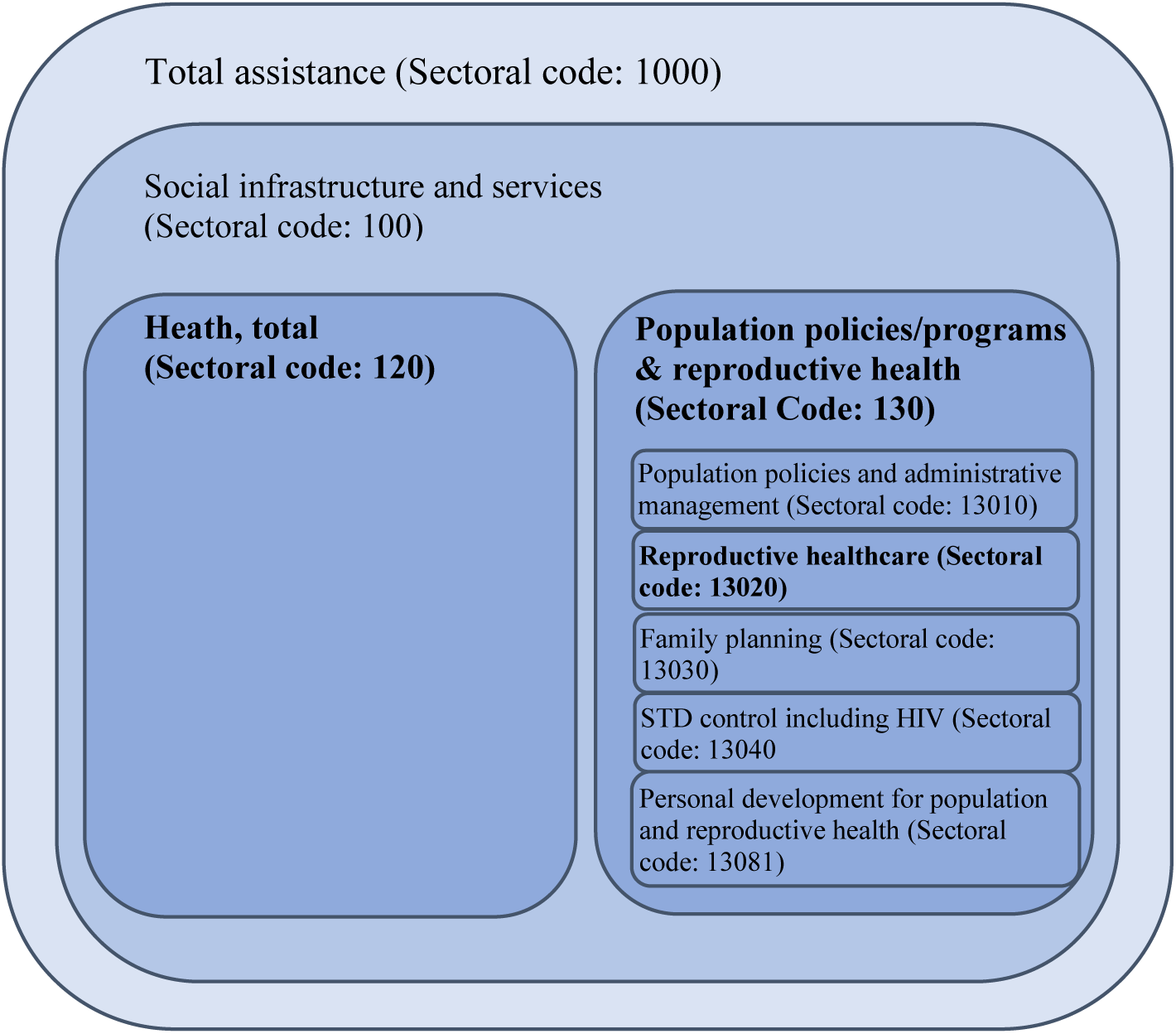
Health ODA explanatory variables and their corresponding sectoral code in the OECD-CRS database. ***Note.*** Figure A1 is an illustration of the sectoral structure of the OECD-CRS database. The bolded sectors represent the health ODA variables used in this study. The combination of sectors 120 and sector 130 is considered total Health ODA. The size of the individual sectors in the schema is not representative of the amount of ODA channelled towards these domains. ***Source***. Author.

## Appendix B. Descriptive Statistics

**Table B1.** Descriptive statistics for the sample used in contraceptive prevalence analyses.

| VARIABLES | N | mean | sd | min | max |
| --- | --- | --- | --- | --- | --- |
| Contraceptive prevalence | 384 | 36.40 | 20.20 | 3.700 | 84 |
| Total Health ODA pc | 384 | 8.281 | 10.22 | 0.0317 | 69.68 |
| SRH ODA pc | 382 | 3.648 | 6.870 | 0.00282 | 57.78 |
| RH service ODA pc | 378 | 0.504 | 0.784 | -0.0254 | 6.037 |
| Population density | 384 | 119.0 | 163.7 | 1.589 | 1,655 |
| GDP pc (PPP) | 384 | 5,671 | 4,948 | 554.9 | 34,280 |
| GHE/CHE (%) | 384 | 37.06 | 17.89 | 4.342 | 78.35 |
| Government effectiveness | 384 | -0.548 | 0.471 | -1.746 | 0.693 |
| Control of corruption | 384 | -0.614 | 0.458 | -1.638 | 0.913 |
| Adolescent fertility | 384 | 76.64 | 43.61 | 6.364 | 210.8 |
| Primary enrolment (%) | 384 | 104.7 | 16.45 | 48.99 | 150.0 |
| Female labour market (%) | 384 | 54.46 | 18.69 | 6.880 | 87.14 |
| Year | 384 | 2010 | 4.740 | 2002 | 2018 |
| Number of countries | 96 |  |  |  |  |

**Note.** Table B1 presents descriptive statistics for the sample of 96 LLMICs that had data on mCP and relevant controls for at least two time points between 2002-2019. No country had information for 2019. The unit of observation is country-year. There is a total of 384 observations. Regarding negative ODA values, loan repayments are recorded as negative and deducted from ODA and loans. In some cases, repayments are higher than new ODA and net ODA will show as a negative number.
**Data sources.** OECD-CRS, WDI, WGI.

**Table B2.** Descriptive statistics for the sample used in skilled birth attendance analyses.

| VARIABLES | N | mean | sd | min | max |
| --- | --- | --- | --- | --- | --- |
| Skilled Birth Attendance | 670 | 80.96 | 22.99 | 5.700 | 100 |
| Total Health ODA pc | 670 | 7.393 | 10.38 | 0.0312 | 124.4 |
| SRH ODA pc | 661 | 2.607 | 5.459 | 0.00290 | 57.78 |
| RH service ODA pc | 640 | 0.435 | 0.721 | -0.0254 | 6.037 |
| Population density | 670 | 111.4 | 172.1 | 1.589 | 1,655 |
| GDP pc (PPP) | 670 | 7,310 | 5,174 | 569.3 | 34,280 |
| GHE/CHE (%) | 670 | 43.21 | 18.91 | 4.342 | 80.50 |
| Government effectiveness | 670 | -0.458 | 0.493 | -1.746 | 0.945 |
| Control of corruption | 670 | -0.545 | 0.569 | -1.673 | 1.641 |
| Adolescent fertility | 670 | 63.43 | 39.19 | 6.364 | 210.8 |
| Primary enrolment (%) | 670 | 104.6 | 14.10 | 48.99 | 150.0 |
| Female labour market (%) | 670 | 51.56 | 15.92 | 6.880 | 86.71 |
| Gini index | 308 | 40.03 | 8.919 | 24 | 63 |
| Year | 670 | 2010 |  | 2002 | 2018 |
| Number of countries | 101 |  |  |  |  |

**Note.** Table B2 presents descriptive statistics for the sample of 101 LLMICs that had data on SBA and relevant controls for at least two time points between 2002-2019. No country had information for 2019. The unit of observation is country-year. There is a total of 670 country-year observations.
**Data sources.** OECD-CRS, WDI, WGI.

**Table B3.** Descriptive statistics for ART coverage sample LLMIC.

| VARIABLES | N | mean | sd | min | max |
| --- | --- | --- | --- | --- | --- |
| ART coverage | 1,295 | 21.49 | 20.22 | 0 | 94 |
| Total Health ODA pc | 1,295 | 8.144 | 10.13 | 0.0312 | 90.30 |
| SRH ODA pc | 1,295 | 3.882 | 7.672 | 0.0123 | 74.95 |
| RH service ODA pc | 1,282 | 0.492 | 0.807 | -0.00868 | 9.386 |
| Population density | 1,295 | 92.29 | 97.29 | 1.573 | 498.7 |
| GDP pc (PPP) | 1,295 | 5,955 | 5,044 | 441.8 | 38,408 |
| GHE/CHE (%) | 1,295 | 37.64 | 17.80 | 0.891 | 79.55 |
| Government effectiveness | 1,295 | -0.574 | 0.496 | -1.776 | 0.785 |
| Control of corruption | 1,295 | -0.611 | 0.503 | -1.766 | 1.641 |
| Adolescent fertility | 1,295 | 78.62 | 45.44 | 6.091 | 215.5 |
| Primary enrolment (%) | 1,295 | 103.5 | 16.96 | 39.54 | 150.0 |
| Female labour market (%) | 1,295 | 53.18 | 18.36 | 6.880 | 87.68 |
| Gini index | 469 | 40.70 | 9.098 | 24 | 64.80 |
| Year | 1,295 | 2010 |  | 2002 | 2018 |
| Number of countries | 90 |  |  |  |  |

**Note.** Table B3 presents descriptive statistics for the sample of 90 LLMICs that had data on ART coverage and relevant controls for at least two time points between 2002-2019. No country had information for 2019. The unit of observation is country-year. There is a total of 1,295 country-year observations.
**Data sources.** OECD-CRS, WDI, WGI.

## Appendix C. Main results

The tables below present the full estimation results, including the estimator selection procedures and relevant diagnostics. Note that in Tables A.1, A.2, A.5, and A.9, the variable Health ODA pc is a placeholder for the type of ODA specified in the column headings of each model.

**Table C.1:**
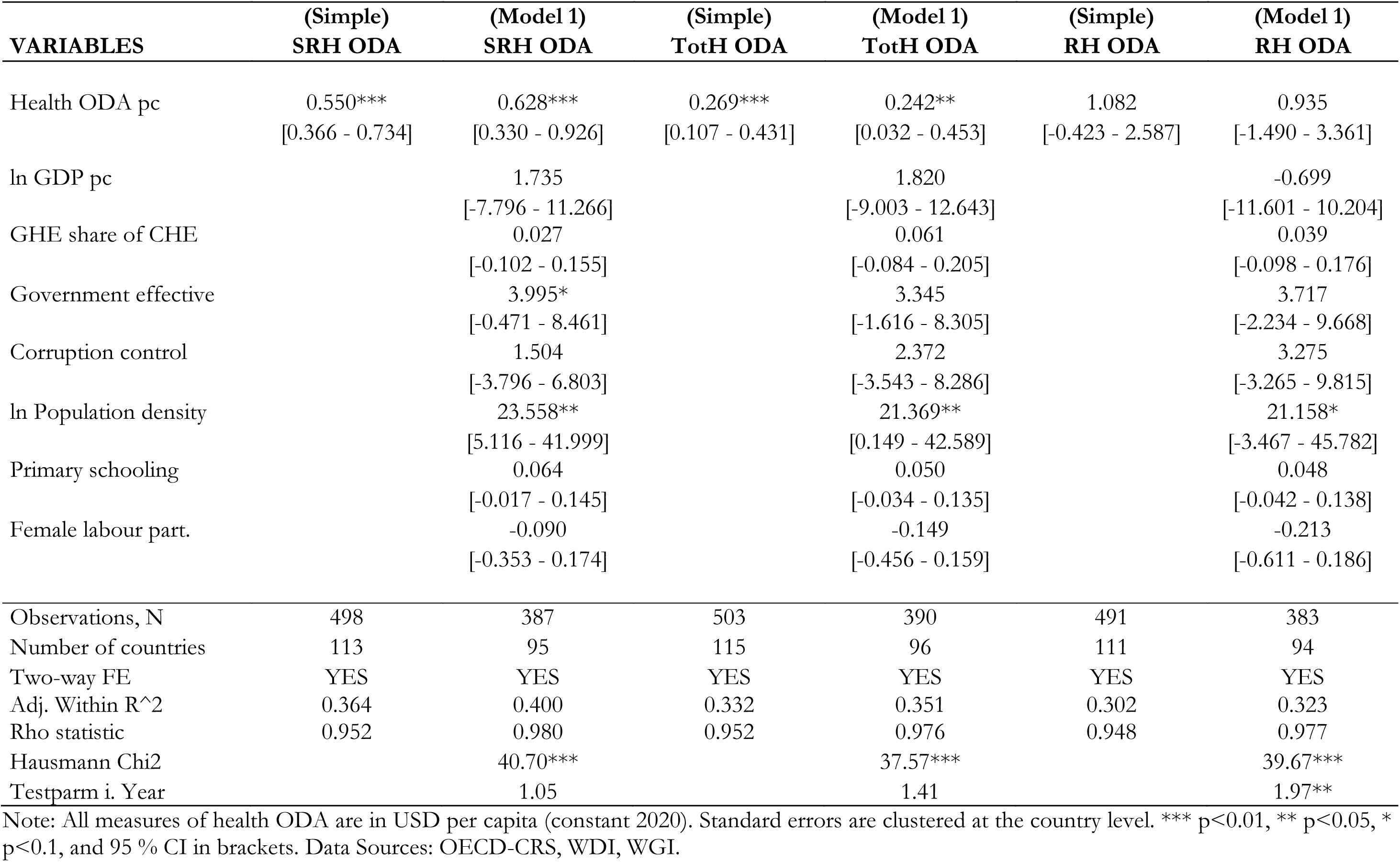
FE estimation of the effects of sectoral ODA on modern contraceptive prevalence.

| **VARIABLES** | **(Simple)**<br>**SRH ODA** | **(Model 1)**<br>**SRH ODA** | **(Simple)**<br>**TotH ODA** | **(Model 1)**<br>**TotH ODA** | **(Simple)**<br>**RH ODA** | **(Model 1)**<br>**RH ODA** |
| --- | --- | --- | --- | --- | --- | --- |
| Health ODA pc | 0.550***<br>[0.366 - 0.734] | 0.628***<br>[0.330 - 0.926] | 0.269***<br>[0.107 - 0.431] | 0.242**<br>[0.032 - 0.453] | 1.082<br>[-0.423 - 2.587] | 0.935<br>[-1.490 - 3.361] |
| ln GDP pc |  | 1.735<br>[-7.796 - 11.266] |  | 1.820<br>[-9.003 - 12.643] |  | -0.699<br>[-11.601 - 10.204] |
| GHE share of CHE |  | 0.027<br>[-0.102 - 0.155] |  | 0.061<br>[-0.084 - 0.205] |  | 0.039<br>[-0.098 - 0.176] |
| Government effective |  | 3.995*<br>[-0.471 - 8.461] |  | 3.345<br>[-1.616 - 8.305] |  | 3.717<br>[-2.234 - 9.668] |
| Corruption control |  | 1.504<br>[-3.796 - 6.803] |  | 2.372<br>[-3.543 - 8.286] |  | 3.275<br>[-3.265 - 9.815] |
| ln Population density |  | 23.558**<br>[5.116 - 41.999] |  | 21.369**<br>[0.149 - 42.589] |  | 21.158*<br>[-3.467 - 45.782] |
| Primary schooling |  | 0.064<br>[-0.017 - 0.145] |  | 0.050<br>[-0.034 - 0.135] |  | 0.048<br>[-0.042 - 0.138] |
| Female labour part. |  | -0.090<br>[-0.353 - 0.174] |  | -0.149<br>[-0.456 - 0.159] |  | -0.213<br>[-0.611 - 0.186] |
| Observations, N | 498 | 387 | 503 | 390 | 491 | 383 |
| Number of countries | 113 | 95 | 115 | 96 | 111 | 94 |
| Two-way FE | YES | YES | YES | YES | YES | YES |
| Adj. Within R<sup>2</sup> | 0.364 | 0.400 | 0.332 | 0.351 | 0.302 | 0.323 |
| Rho statistic | 0.952 | 0.980 | 0.952 | 0.976 | 0.948 | 0.977 |
| Hausmann Chi2 |  | 40.70*** |  | 37.57*** |  | 39.67*** |
| Testparm i. Year |  | 1.05 |  | 1.41 |  | 1.97** |

Note: All measures of health ODA are in USD per capita (constant 2020). Standard errors are clustered at the country level. \*\*\* p<0.01, \*\* p<0.05, \* p<0.1, and 95 % CI in brackets. Data Sources: OECD-CRS, WDI, WGI.

**Table C.2:**
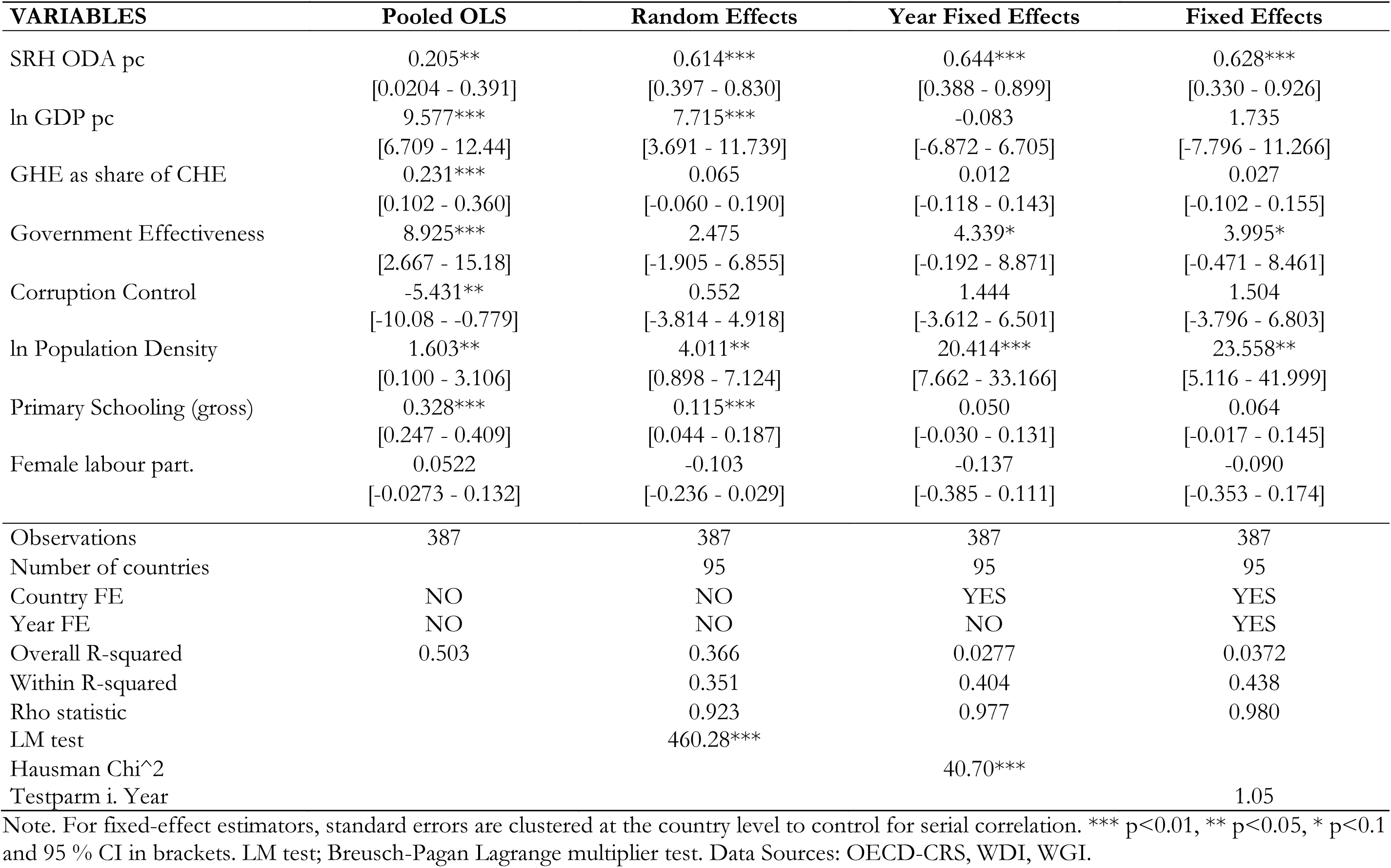
Effects of SRH ODA per capita on modern contraceptive prevalence.

| **VARIABLES** | **Pooled OLS** | **Random Effects** | **Year Fixed Effects** | **Fixed Effects** |
| --- | --- | --- | --- | --- |
| SRH ODA pc | 0.205**<br>[0.0204 - 0.391] | 0.614***<br>[0.397 - 0.830] | 0.644***<br>[0.388 - 0.899] | 0.628***<br>[0.330 - 0.926] |
| ln GDP pc | 9.577***<br>[6.709 - 12.44] | 7.715***<br>[3.691 - 11.739] | -0.083<br>[-6.872 - 6.705] | 1.735<br>[-7.796 - 11.266] |
| GHE as share of CHE | 0.231***<br>[0.102 - 0.360] | 0.065<br>[-0.060 - 0.190] | 0.012<br>[-0.118 - 0.143] | 0.027<br>[-0.102 - 0.155] |
| Government Effectiveness | 8.925***<br>[2.667 - 15.18] | 2.475<br>[-1.905 - 6.855] | 4.339*<br>[-0.192 - 8.871] | 3.995*<br>[-0.471 - 8.461] |
| Corruption Control | -5.431**<br>[-10.08 - -0.779] | 0.552<br>[-3.814 - 4.918] | 1.444<br>[-3.612 - 6.501] | 1.504<br>[-3.796 - 6.803] |
| ln Population Density | 1.603**<br>[0.100 - 3.106] | 4.011**<br>[0.898 - 7.124] | 20.414***<br>[7.662 - 33.166] | 23.558**<br>[5.116 - 41.999] |
| Primary Schooling (gross) | 0.328***<br>[0.247 - 0.409] | 0.115***<br>[0.044 - 0.187] | 0.050<br>[-0.030 - 0.131] | 0.064<br>[-0.017 - 0.145] |
| Female labour part. | 0.0522<br>[-0.0273 - 0.132] | -0.103<br>[-0.236 - 0.029] | -0.137<br>[-0.385 - 0.111] | -0.090<br>[-0.353 - 0.174] |
| Observations | 387 | 387 | 387 | 387 |
| Number of countries |  | 95 | 95 | 95 |
| Country FE | NO | NO | YES | YES |
| Year FE | NO | NO | NO | YES |
| Overall R-squared | 0.503 | 0.366 | 0.0277 | 0.0372 |
| Within R-squared |  | 0.351 | 0.404 | 0.438 |
| Rho statistic |  | 0.923 | 0.977 | 0.980 |
| LM test |  | 460.28*** |  |  |
| Hausman Chi<sup>2</sup> |  |  | 40.70*** |  |
| Testparm i. Year |  |  |  | 1.05 |

Note. For fixed-effect estimators, standard errors are clustered at the country level to control for serial correlation. \*\*\* p<0.01, \*\* p<0.05, \* p<0.1 and 95 % CI in brackets. LM test; Breusch-Pagan Lagrange multiplier test. Data Sources: OECD-CRS, WDI, WGI.

**Table C.3:** Effects of Total Health ODA per capita on modern contraceptive prevalence.

| **VARIABLES** | **Pooled OLS** | **Random Effects** | **Year Fixed Effects** | **Fixed Effects** |
| --- | --- | --- | --- | --- |
| Total Health ODA pc | -0.0828<br>[-0.254 - 0.0888] | 0.272***<br>[0.095 - 0.449] | 0.269**<br>[0.057 - 0.481] | 0.242**<br>[0.032 - 0.453] |
| ln GDP pc | 8.922***<br>[6.020 - 11.82] | 7.630***<br>[3.303 - 11.957] | 0.985<br>[-6.792 - 8.763] | 1.820<br>[-9.003 - 12.643] |
| GHE as share of CHE | 0.203***<br>[0.0782 - 0.327] | 0.078<br>[-0.056 - 0.212] | 0.039<br>[-0.106 - 0.184] | 0.061<br>[-0.084 - 0.205] |
| Government Effectiveness | 9.149***<br>[3.028 - 15.27] | 2.480<br>[-2.047 - 7.007] | 3.849<br>[-1.135 - 8.833] | 3.345<br>[-1.616 - 8.305] |
| Corruption Control | -4.312*<br>[-9.423 - 0.799] | 0.857<br>[-3.894 - 5.608] | 1.932<br>[-3.532 - 7.396] | 2.372<br>[-3.543 - 8.286] |
| ln Population Density | 1.154<br>[-0.308 - 2.617] | 3.526**<br>[0.454 - 6.598] | 18.000**<br>[4.182 - 31.818] | 21.369**<br>[0.149 - 42.589] |
| Primary Schooling (gross) | 0.360***<br>[0.279 - 0.442] | 0.109***<br>[0.034 - 0.184] | 0.044<br>[-0.041 - 0.129] | 0.050<br>[-0.034 - 0.135] |
| Female labour part. | 0.0559<br>[-0.0239 - 0.136] | -0.104<br>[-0.244 - 0.037] | -0.185<br>[-0.475 - 0.104] | -0.149<br>[-0.456 - 0.159] |
| Observations, N | 390 | 390 | 390 | 390 |
| Number of countries |  | 96 | 96 | 96 |
| Country FE | NO | NO | YES | YES |
| Year FE | NO | NO | NO | YES |
| Overall R-squared | 0.485 | 0.345 | 0.0340 | 0.0381 |
| Within R-squared |  | 0.310 | 0.353 | 0.392 |
| Rho statistic |  | 0.920 | 0.972 | 0.976 |
| LM test |  | 420.23*** |  |  |
| Hausmann Chi2 |  |  | 37.57*** |  |
| Testparm i. Year |  |  |  | 1.41 |

Note. For fixed-effect estimators, standard errors are clustered at the country level to control for serial correlation. \*\*\* p<0.01, \*\* p<0.05, \* p<0.1 and 95 % CI in brackets. LM test; Breusch-Pagan Lagrange multiplier test. Data Sources: OECD-CRS, WDI, WGI.

**Table C.4:** Effects of RH care ODA per capita on modern contraceptive prevalence.

| **VARIABLES** | **Pooled OLS** | **Random Effects** | **Country Fixed Effects** | **Fixed Effects** |
| --- | --- | --- | --- | --- |
| RH care ODA pc | -1.348<br>[-3.074 - 0.379] | 1.225<br>[-0.948 - 3.397] | 1.117<br>[-1.287 - 3.522] | 0.935<br>[-1.490 - 3.361] |
| ln GDP pc (PPP, constant) | 9.181***<br>[6.336 - 12.03] | 8.545***<br>[4.147 - 12.943] | 1.114<br>[-7.347 - 9.575] | -0.699<br>[-11.601 - 10.204] |
| GHE as share of CHE | 0.214***<br>[0.0868 - 0.341] | 0.069<br>[-0.057 - 0.195] | 0.014<br>[-0.121 - 0.150] | 0.039<br>[-0.098 - 0.176] |
| Government Effectiveness | 8.714***<br>[2.539 - 14.89] | 2.472<br>[-2.929 - 7.873] | 4.480<br>[-1.785 - 10.745] | 3.717<br>[-2.234 - 9.668] |
| Corruption Control | -3.869<br>[-8.770 - 1.032] | 1.610<br>[-3.872 - 7.091] | 2.268<br>[-3.812 - 8.348] | 3.275<br>[-3.265 - 9.815] |
| ln Population Density | 1.261*<br>[-0.172 - 2.695] | 3.475**<br>[0.355 - 6.596] | 21.223***<br>[6.764 - 35.682] | 21.158*<br>[-3.467 - 45.782] |
| Primary Schooling (gross) | 0.356***<br>[0.278 - 0.434] | 0.129***<br>[0.049 - 0.209] | 0.046<br>[-0.040 - 0.133] | 0.048<br>[-0.042 - 0.138] |
| Female labour part. | 0.0438<br>[-0.0393 - 0.127] | -0.098<br>[-0.248 - 0.053] | -0.237<br>[-0.617 - 0.143] | -0.213<br>[-0.611 - 0.186] |
| Observations | 383 | 383 | 383 | 383 |
| Number of countries |  | 94 | 94 | 94 |
| Country FE | NO | NO | YES | YES |
| Year FE | NO | NO | YES | YES |
| Overall R-squared | 0.508 | 0.394 | 0.0345 | 0.0261 |
| Within R-squared |  | 0.252 | 0.316 | 0.367 |
| Rho statistic |  | 0.912 | 0.976 | 0.977 |
| LM test |  | 410.10*** |  |  |
| Hausmann Chi2 |  |  | 39.67*** |  |
| Testparm i. Year |  |  |  | 1.97** |

Note. For fixed-effect estimators, standard errors are clustered at the country level to control for serial correlation. \*\*\* p<0.01, \*\* p<0.05, \* p<0.1 and 95 % CI in brackets. LM test; Breusch-Pagan Lagrange multiplier test. Data Sources: OECD-CRS, WDI, WGI.

**Table C.5:**
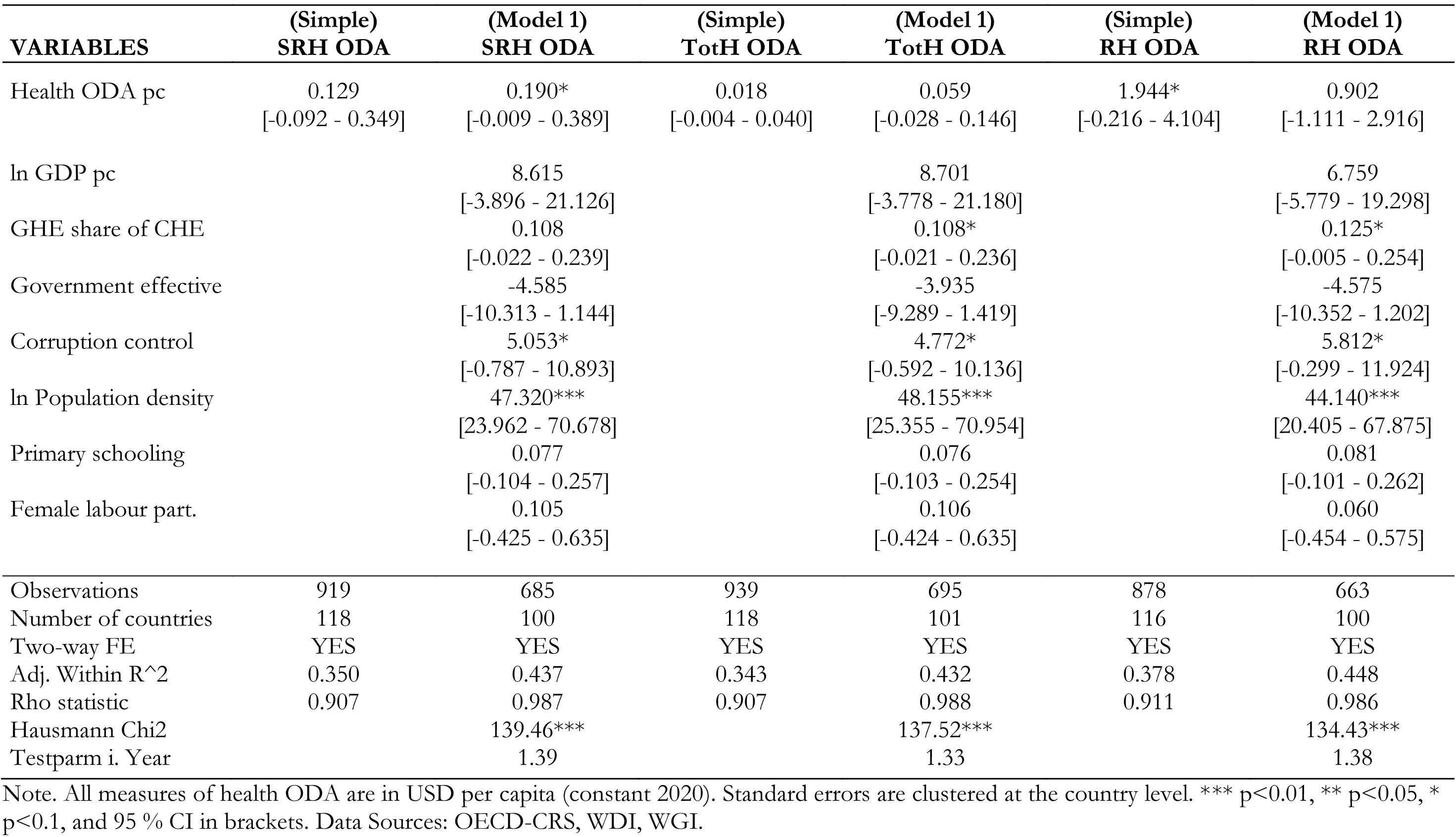
FE estimation of the effects of sectoral ODA on skilled birth attendance.

**Table C.6:** Effects of SRH ODA per capita on skilled birth attendance.

| **VARIABLES** | **Pooled OLS** | **Random Effects** | **Country Fixed Effects** | **Fixed Effects** |
| --- | --- | --- | --- | --- |
| SRH ODA pc | 0.0462<br>[-0.104 - 0.196] | 0.198*<br>[-0.000 - 0.395] | 0.104<br>[-0.107 - 0.315] | 0.190*<br>[-0.009 - 0.389] |
| ln GDP pc | 15.91***<br>[13.49 - 18.32] | 20.237***<br>[14.189 - 26.285] | 7.914<br>[-1.772 - 17.601] | 8.615<br>[-3.896 - 21.126] |
| GHE share of CHE | 0.198***<br>[0.111 - 0.286] | 0.166**<br>[0.033 - 0.300] | 0.109*<br>[-0.014 - 0.232] | 0.108<br>[-0.022 - 0.239] |
| Government effective | 5.397**<br>[1.280 - 9.514] | -5.054**<br>[-10.108 - -0.001] | -3.036<br>[-8.401 - 2.328] | -4.585<br>[-10.313 - 1.144] |
| Corruption control | -4.941***<br>[-8.147 - -1.735] | 2.982<br>[-3.430 - 9.393] | 4.848<br>[-1.326 - 11.022] | 5.053*<br>[-0.787 - 10.893] |
| ln Population density | 0.232<br>[-0.840 - 1.305] | 5.504***<br>[2.340 - 8.668] | 51.833***<br>[35.542 - 68.124] | 47.320***<br>[23.962 - 70.678] |
| Primary schooling | 0.193***<br>[0.0998 - 0.287] | 0.147*<br>[-0.015 - 0.309] | 0.061<br>[-0.118 - 0.240] | 0.077<br>[-0.104 - 0.257] |
| Female labour part. | -0.0418<br>[-0.134 - 0.0500] | 0.034<br>[-0.203 - 0.272] | 0.106<br>[-0.430 - 0.642] | 0.105<br>[-0.425 - 0.635] |
| Observations | 685 | 685 | 685 | 685 |
| Number of countries |  | 100 | 100 | 100 |
| Country FE | NO | NO | YES | YES |
| Year FE | NO | NO | NO | YES |
| Overall R-squared | 0.531 | 0.470 | 0.00330 | 0.00490 |
| Within R-squared |  | 0.310 | 0.427 | 0.458 |
| Rho statistic |  | 0.820 | 0.989 | 0.987 |
| LM test |  | 1436.30*** |  |  |
| Hausmann Chi2 |  |  | 139.46*** |  |
| Testparm i. Year |  |  |  | 1.39 |

Note. For fixed-effect estimators, standard errors are clustered at the country level to control for serial correlation. \*\*\* p<0.01, \*\* p<0.05, \* p<0.1 and 95 % CI in brackets. LM test; Breusch-Pagan Lagrange multiplier.

**Table C.7:** Effects of Total Health ODA per capita on skilled birth attendance.

| **VARIABLES** | **Pooled OLS** | **Random Effects** | **Country Fixed Effects** | **Fixed Effects** |
| --- | --- | --- | --- | --- |
| Tot Health ODA pc | 0.217***<br>[0.108 - 0.326] | 0.136**<br>[0.013 - 0.259] | 0.044<br>[-0.035 - 0.123] | 0.059<br>[-0.028 - 0.146] |
| ln GDP pc | 16.90***<br>[14.36 - 19.44] | 19.994***<br>[14.069 - 25.919] | 7.917<br>[-1.622 - 17.456] | 8.701<br>[-3.778 - 21.180] |
| GHE share of CHE | 0.192***<br>[0.106 - 0.278] | 0.161**<br>[0.029 - 0.293] | 0.109*<br>[-0.013 - 0.231] | 0.108*<br>[-0.021 - 0.236] |
| Government effective | 5.152**<br>[1.126 - 9.178] | -4.577*<br>[-9.394 - 0.239] | -2.691<br>[-7.711 - 2.329] | -3.935<br>[-9.289 - 1.419] |
| Corruption control | -5.576***<br>[-8.687 - -2.466] | 3.075<br>[-2.782 - 8.933] | 4.535<br>[-1.123 - 10.193] | 4.772*<br>[-0.592 - 10.136] |
| ln Population density | 0.445<br>[-0.596 - 1.486] | 5.228***<br>[2.163 - 8.292] | 51.506***<br>[35.306 - 67.705] | 48.155***<br>[25.355 - 70.954] |
| Primary schooling | 0.170***<br>[0.0778 - 0.263] | 0.140*<br>[-0.020 - 0.300] | 0.060<br>[-0.117 - 0.238] | 0.076<br>[-0.103 - 0.254] |
| Female labour part. | -0.0297<br>[-0.121 - 0.0618] | 0.034<br>[-0.200 - 0.268] | 0.103<br>[-0.434 - 0.640] | 0.106<br>[-0.424 - 0.635] |
| Observations | 695 | 695 | 695 | 695 |
| Number of countries |  | 101 | 101 | 101 |
| Country FE | NO | NO | YES | YES |
| Year FE | NO | NO | NO | YES |
| Overall R-squared | 0.539 | 0.480 | 0.00451 | 0.00629 |
| Within R-squared |  | 0.307 | 0.423 | 0.452 |
| Rho statistic |  | 0.817 | 0.989 | 0.988 |
| LM test |  | 1458.89*** |  |  |
| Hausmann Chi2 |  |  | 137.52*** |  |
| Testparm i. Year |  |  |  | 1.33 |

Note. For fixed-effect estimators, standard errors are clustered at the country level to control for serial correlation. \*\*\* p<0.01, \*\* p<0.05, \* p<0.1 and 95 % CI in brackets. LM test; Breusch-Pagan Lagrange multiplier test. Data Sources: OECD-CRS, WDI, WGI.

**Table C.8:** Effects of RH Care ODA per capita on skilled birth attendance.

| **VARIABLES** | **Pooled OLS** | **Random Effects** | **Country Fixed Effects** | **Fixed Effects** |
| --- | --- | --- | --- | --- |
| RH care ODA pc | 0.00677<br>[-1.814 - 1.827] | 1.494<br>[-0.751 - 3.740] | 0.798<br>[-1.341 - 2.938] | 0.902<br>[-1.111 - 2.916] |
| ln GDP pc | 16.31***<br>[13.77 - 18.84] | 20.299***<br>[14.259 - 26.339] | 7.951<br>[-1.862 - 17.764] | 6.759<br>[-5.779 - 19.298] |
| GHE share of CHE | 0.176***<br>[0.0853 - 0.267] | 0.182***<br>[0.050 - 0.314] | 0.127**<br>[0.004 - 0.249] | 0.125*<br>[-0.005 - 0.254] |
| Government effective | 4.794**<br>[0.625 - 8.964] | -5.293**<br>[-10.457 - -0.128] | -3.132<br>[-8.553 - 2.289] | -4.575<br>[-10.352 - 1.202] |
| Corruption control | -4.201**<br>[-7.527 - -0.876] | 3.901<br>[-2.832 - 10.634] | 5.735*<br>[-0.686 - 12.156] | 5.812*<br>[-0.299 - 11.924] |
| ln Population density | 0.0696<br>[-1.015 - 1.154] | 5.187***<br>[2.091 - 8.283] | 51.485***<br>[35.177 - 67.794] | 44.140***<br>[20.405 - 67.875] |
| Primary schooling | 0.196***<br>[0.103 - 0.288] | 0.151*<br>[-0.012 - 0.313] | 0.063<br>[-0.117 - 0.242] | 0.081<br>[-0.101 - 0.262] |
| Female labour part. | -0.0355<br>[-0.130 - 0.0590] | 0.033<br>[-0.200 - 0.267] | 0.082<br>[-0.447 - 0.612] | 0.060<br>[-0.454 - 0.575] |
| Observations | 663 | 663 | 663 | 663 |
| Number of countries |  | 100 | 100 | 100 |
| Country FE | NO | NO | YES | YES |
| Year FE | NO | NO | NO | YES |
| Overall R-squared | 0.530 | 0.474 | 0.00374 | 0.00416 |
| Within R-squared |  | 0.324 | 0.439 | 0.469 |
| Rho statistic |  | 0.820 | 0.989 | 0.986 |
| LM test |  | 1439.71*** |  |  |
| Hausmann Chi2 |  |  | 134.43*** |  |
| Testparm i. Year |  |  |  | 1.38 |

Note. For fixed-effect estimators, standard errors are clustered at the country level to control for serial correlation. \*\*\* p<0.01, \*\* p<0.05, \* p<0.1 and 95 % CI in brackets. LM test; Breusch-Pagan Lagrange multiplier test. Data Sources: OECD-CRS, WDI, WGI.

**Table C.9:**
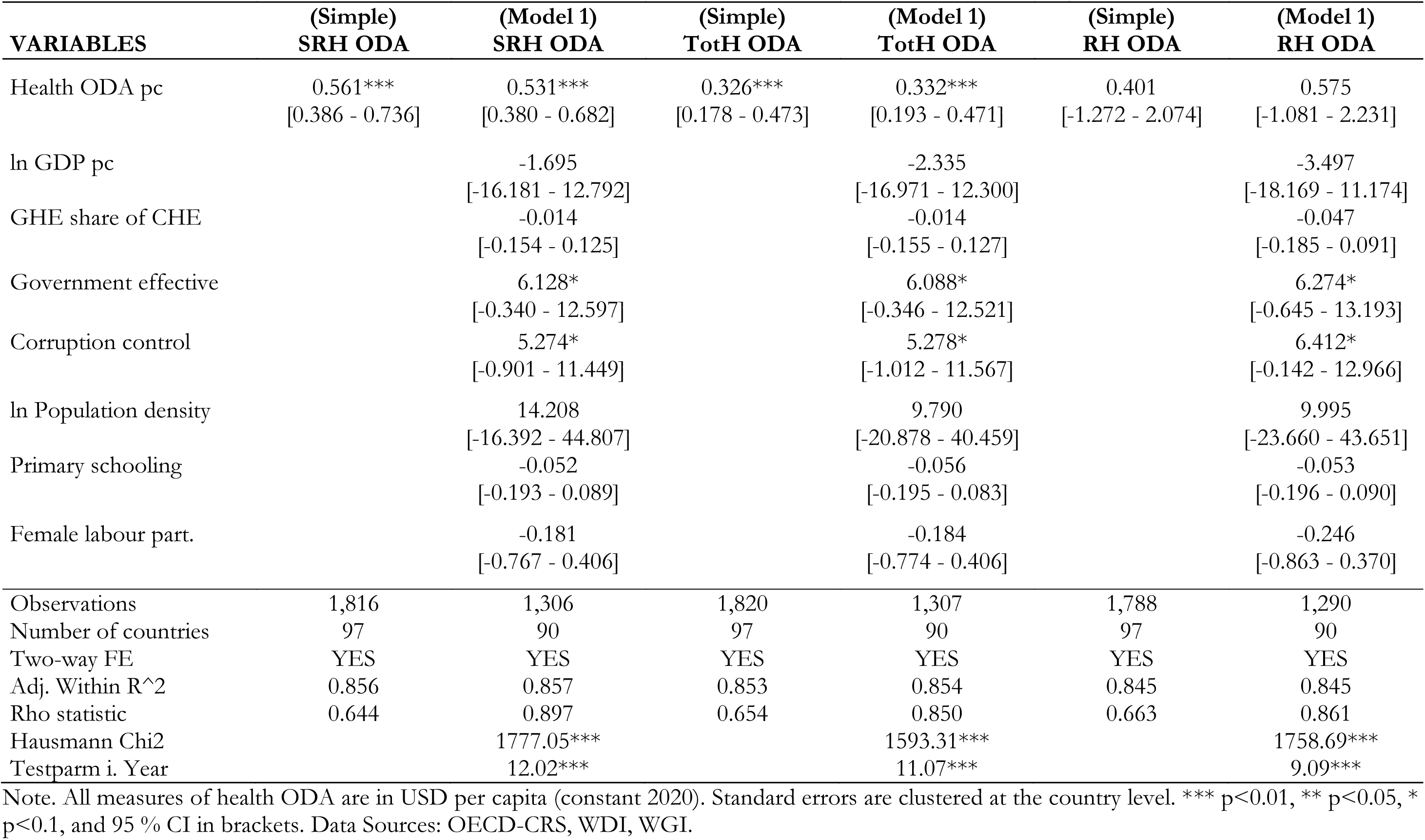
FE estimation of the effects of sectoral ODA on ART coverage.

| **VARIABLES** | **(Simple)**<br>**SRH ODA** | **(Model 1)**<br>**SRH ODA** | **(Simple)**<br>**TotH ODA** | **(Model 1)**<br>**TotH ODA** | **(Simple)**<br>**RH ODA** | **(Model 1)**<br>**RH ODA** |
| --- | --- | --- | --- | --- | --- | --- |
| Health ODA pc | 0.561***<br>[0.386 - 0.736] | 0.531***<br>[0.380 - 0.682] | 0.326***<br>[0.178 - 0.473] | 0.332***<br>[0.193 - 0.471] | 0.401<br>[-1.272 - 2.074] | 0.575<br>[-1.081 - 2.231] |
| ln GDP pc |  | -1.695<br>[-16.181 - 12.792] |  | -2.335<br>[-16.971 - 12.300] |  | -3.497<br>[-18.169 - 11.174] |
| GHE share of CHE |  | -0.014<br>[-0.154 - 0.125] |  | -0.014<br>[-0.155 - 0.127] |  | -0.047<br>[-0.185 - 0.091] |
| Government effective |  | 6.128*<br>[-0.340 - 12.597] |  | 6.088*<br>[-0.346 - 12.521] |  | 6.274*<br>[-0.645 - 13.193] |
| Corruption control |  | 5.274*<br>[-0.901 - 11.449] |  | 5.278*<br>[-1.012 - 11.567] |  | 6.412*<br>[-0.142 - 12.966] |
| ln Population density |  | 14.208<br>[-16.392 - 44.807] |  | 9.790<br>[-20.878 - 40.459] |  | 9.995<br>[-23.660 - 43.651] |
| Primary schooling |  | -0.052<br>[-0.193 - 0.089] |  | -0.056<br>[-0.195 - 0.083] |  | -0.053<br>[-0.196 - 0.090] |
| Female labour part. |  | -0.181<br>[-0.767 - 0.406] |  | -0.184<br>[-0.774 - 0.406] |  | -0.246<br>[-0.863 - 0.370] |
| Observations | 1,816 | 1,306 | 1,820 | 1,307 | 1,788 | 1,290 |
| Number of countries | 97 | 90 | 97 | 90 | 97 | 90 |
| Two-way FE | YES | YES | YES | YES | YES | YES |
| Adj. Within R<sup>2</sup> | 0.856 | 0.857 | 0.853 | 0.854 | 0.845 | 0.845 |
| Rho statistic | 0.644 | 0.897 | 0.654 | 0.850 | 0.663 | 0.861 |
| Hausmann Chi2 |  | 1777.05*** |  | 1593.31*** |  | 1758.69*** |
| Testparm i. Year |  | 12.02*** |  | 11.07*** |  | 9.09*** |

Note. All measures of health ODA are in USD per capita (constant 2020). Standard errors are clustered at the country level. \*\*\* p<0.01, \*\* p<0.05, \* p<0.1, and 95 % CI in brackets. Data Sources: OECD-CRS, WDI, WGI.

**Table C.10:** Effects of SRH ODA per capita on ART coverage.

| **VARIABLES** | **Pooled OLS** | **Random Effects** | **Country Fixed Effects** | **Fixed Effects** |
| --- | --- | --- | --- | --- |
| SRH ODA pc | 0.713***<br>[0.554 - 0.872] | 0.907***<br>[0.609 - 1.205] | 0.534***<br>[0.243 - 0.826] | 0.531***<br>[0.380 - 0.682] |
| ln GDP pc | 5.111***<br>[3.180 - 7.042] | 35.033***<br>[26.154 - 43.912] | 31.172***<br>[17.930 - 44.413] | -1.695<br>[-16.181 - 12.792] |
| GHE share of CHE | 0.0248<br>[-0.0633 - 0.113] | 0.143<br>[-0.127 - 0.414] | 0.110<br>[-0.086 - 0.306] | -0.014<br>[-0.154 - 0.125] |
| Government effective | 3.033*<br>[-0.487 - 6.554] | 2.013<br>[-6.457 - 10.483] | 10.537***<br>[2.833 - 18.241] | 6.128*<br>[-0.340 - 12.597] |
| Corruption control | 1.146<br>[-2.463 - 4.755] | 1.873<br>[-6.520 - 10.266] | 4.557<br>[-2.580 - 11.695] | 5.274*<br>[-0.901 - 11.449] |
| ln Population density | 2.207***<br>[1.283 - 3.132] | 16.346***<br>[10.537 - 22.156] | 109.281***<br>[87.292 - 131.270] | 14.208<br>[-16.392 - 44.807] |
| Primary schooling | 0.0817**<br>[0.0141 - 0.149] | 0.159<br>[-0.066 - 0.385] | -0.124<br>[-0.302 - 0.054] | -0.052<br>[-0.193 - 0.089] |
| Female labour part. | 0.190***<br>[0.122 - 0.258] | 0.513***<br>[0.139 - 0.887] | 0.312<br>[-0.398 - 1.021] | -0.181<br>[-0.767 - 0.406] |
| Observations | 1,306 | 1,306 | 1,306 | 1,306 |
| Number of countries |  | 90 | 90 | 90 |
| Country FE | NO | NO | YES | YES |
| Year FE | NO | NO | NO | YES |
| Overall R-squared | 0.168 | 0.100 | 0.0205 | 0.356 |
| Within R-squared |  | 0.611 | 0.759 | 0.860 |
| Rho statistic |  | 0.593 | 0.995 | 0.897 |
| LM test |  | 63.51*** |  |  |
| Hausmann Chi2 |  |  | 1777.05*** |  |
| Testparm i. Year |  |  |  | 12.02*** |

Note. For fixed-effect estimators, standard errors are clustered at the country level to control for serial correlation. \*\*\* p<0.01, \*\* p<0.05, \* p<0.1 and 95 % CI in brackets. LM test; Breusch-Pagan Lagrange multiplier test. Data Sources: OECD-CRS, WDI, WGI.

**Table C.11:**
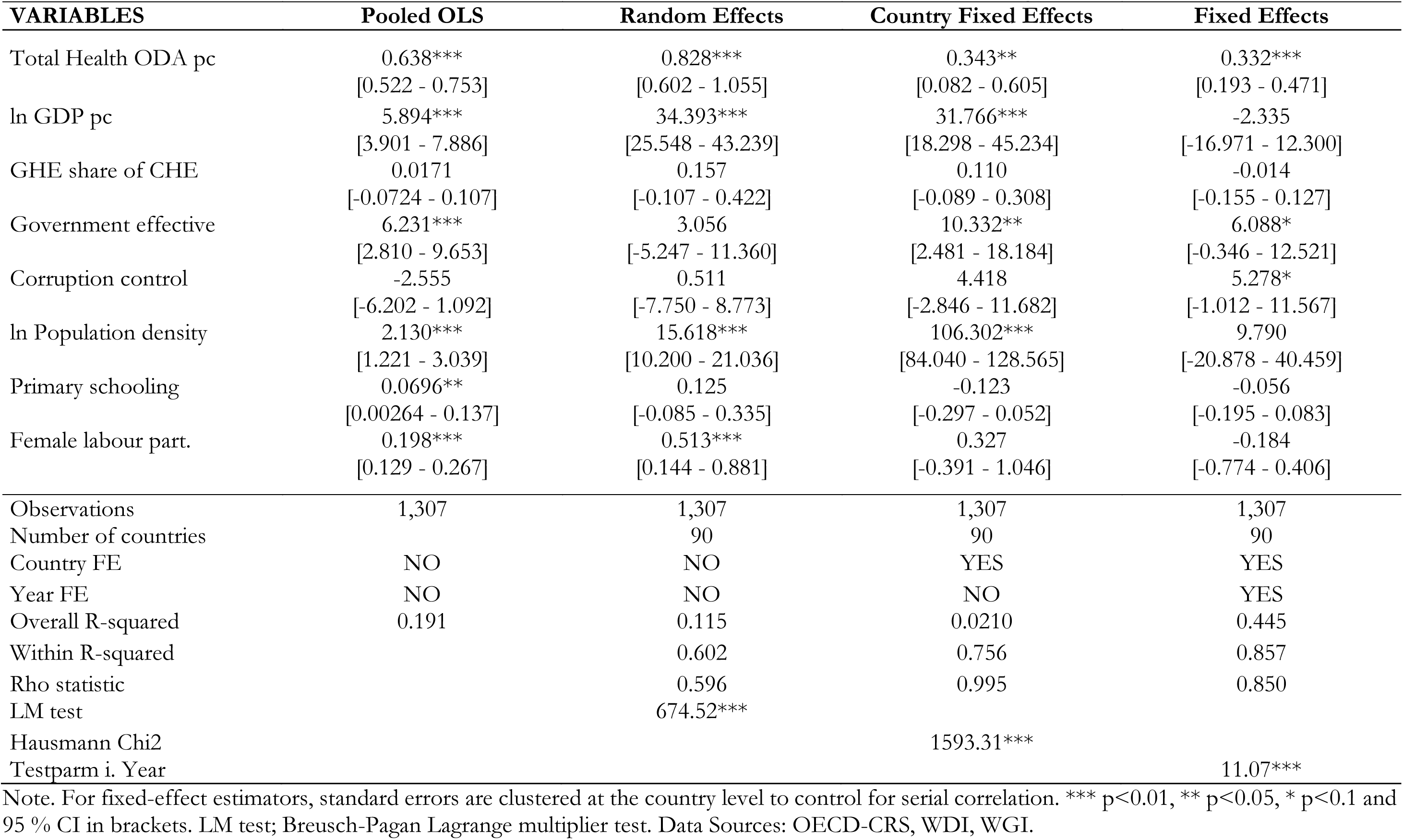
Effects of Total health ODA per capita on ART coverage.

| **VARIABLES** | **Pooled OLS** | **Random Effects** | **Country Fixed Effects** | **Fixed Effects** |
| --- | --- | --- | --- | --- |
| Total Health ODA pc | 0.638***<br>[0.522 - 0.753] | 0.828***<br>[0.602 - 1.055] | 0.343**<br>[0.082 - 0.605] | 0.332***<br>[0.193 - 0.471] |
| ln GDP pc | 5.894***<br>[3.901 - 7.886] | 34.393***<br>[25.548 - 43.239] | 31.766***<br>[18.298 - 45.234] | -2.335<br>[-16.971 - 12.300] |
| GHE share of CHE | 0.0171<br>[-0.0724 - 0.107] | 0.157<br>[-0.107 - 0.422] | 0.110<br>[-0.089 - 0.308] | -0.014<br>[-0.155 - 0.127] |
| Government effective | 6.231***<br>[2.810 - 9.653] | 3.056<br>[-5.247 - 11.360] | 10.332**<br>[2.481 - 18.184] | 6.088*<br>[-0.346 - 12.521] |
| Corruption control | -2.555<br>[-6.202 - 1.092] | 0.511<br>[-7.750 - 8.773] | 4.418<br>[-2.846 - 11.682] | 5.278*<br>[-1.012 - 11.567] |
| ln Population density | 2.130***<br>[1.221 - 3.039] | 15.618***<br>[10.200 - 21.036] | 106.302***<br>[84.040 - 128.565] | 9.790<br>[-20.878 - 40.459] |
| Primary schooling | 0.0696**<br>[0.00264 - 0.137] | 0.125<br>[-0.085 - 0.335] | -0.123<br>[-0.297 - 0.052] | -0.056<br>[-0.195 - 0.083] |
| Female labour part. | 0.198***<br>[0.129 - 0.267] | 0.513***<br>[0.144 - 0.881] | 0.327<br>[-0.391 - 1.046] | -0.184<br>[-0.774 - 0.406] |
| Observations | 1,307 | 1,307 | 1,307 | 1,307 |
| Number of countries |  | 90 | 90 | 90 |
| Country FE | NO | NO | YES | YES |
| Year FE | NO | NO | NO | YES |
| Overall R-squared | 0.191 | 0.115 | 0.0210 | 0.445 |
| Within R-squared |  | 0.602 | 0.756 | 0.857 |
| Rho statistic |  | 0.596 | 0.995 | 0.850 |
| LM test |  | 674.52*** |  |  |
| Hausmann Chi2 |  |  | 1593.31*** |  |
| Testparm i. Year |  |  |  | 11.07*** |

Note. For fixed-effect estimators, standard errors are clustered at the country level to control for serial correlation. \*\*\* p<0.01, \*\* p<0.05, \* p<0.1 and 95 % CI in brackets. LM test; Breusch-Pagan Lagrange multiplier test. Data Sources: OECD-CRS, WDI, WGI.

**Table C.12:**
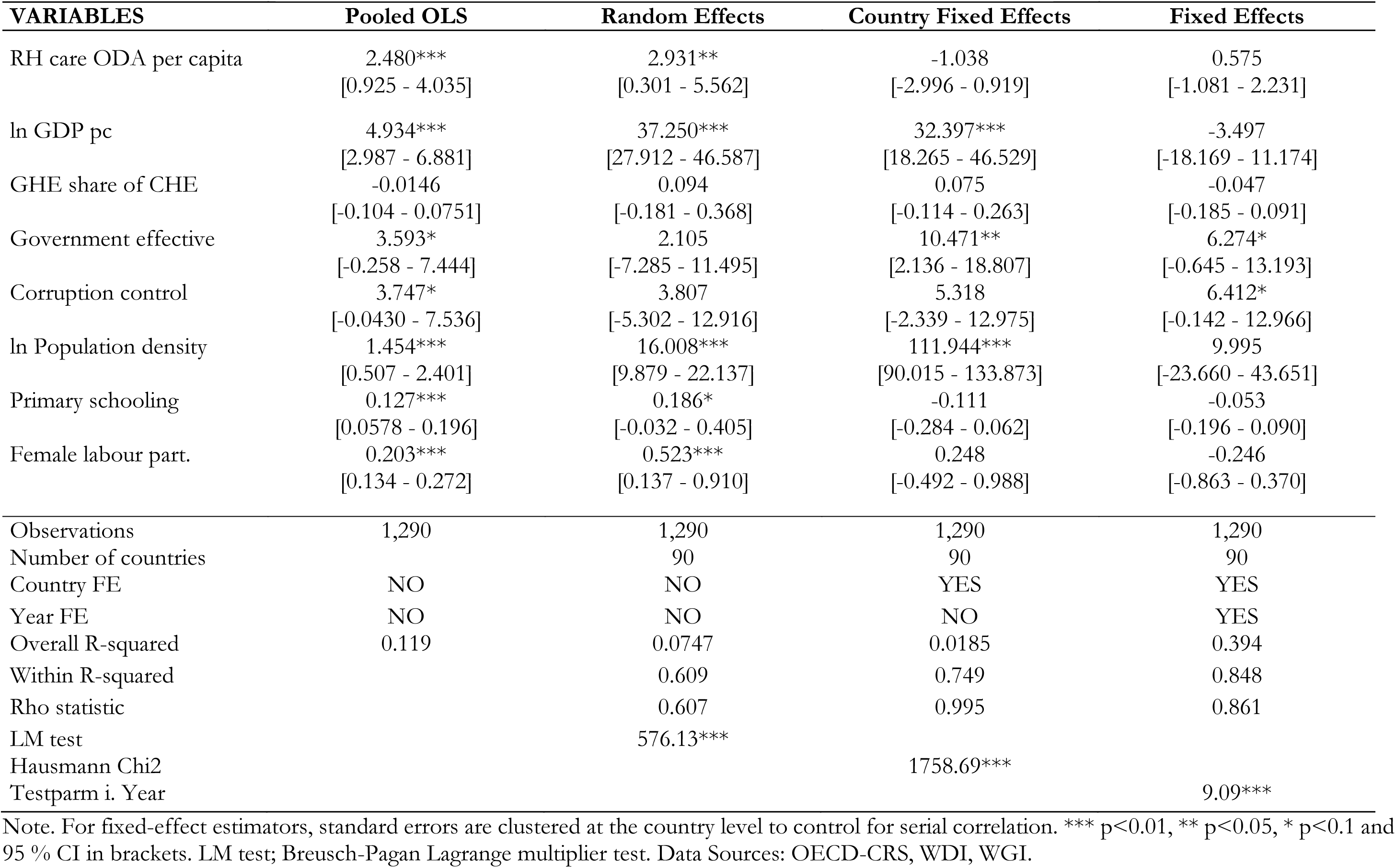
Effects of RH care ODA per capita on ART coverage.

| **VARIABLES** | **Pooled OLS** | **Random Effects** | **Country Fixed Effects** | **Fixed Effects** |
| --- | --- | --- | --- | --- |
| RH care ODA per capita | 2.480***<br>[0.925 - 4.035] | 2.931**<br>[0.301 - 5.562] | -1.038<br>[-2.996 - 0.919] | 0.575<br>[-1.081 - 2.231] |
| ln GDP pc | 4.934***<br>[2.987 - 6.881] | 37.250***<br>[27.912 - 46.587] | 32.397***<br>[18.265 - 46.529] | -3.497<br>[-18.169 - 11.174] |
| GHE share of CHE | -0.0146<br>[-0.104 - 0.0751] | 0.094<br>[-0.181 - 0.368] | 0.075<br>[-0.114 - 0.263] | -0.047<br>[-0.185 - 0.091] |
| Government effective | 3.593*<br>[-0.258 - 7.444] | 2.105<br>[-7.285 - 11.495] | 10.471**<br>[2.136 - 18.807] | 6.274*<br>[-0.645 - 13.193] |
| Corruption control | 3.747*<br>[-0.0430 - 7.536] | 3.807<br>[-5.302 - 12.916] | 5.318<br>[-2.339 - 12.975] | 6.412*<br>[-0.142 - 12.966] |
| ln Population density | 1.454***<br>[0.507 - 2.401] | 16.008***<br>[9.879 - 22.137] | 111.944***<br>[90.015 - 133.873] | 9.995<br>[-23.660 - 43.651] |
| Primary schooling | 0.127***<br>[0.0578 - 0.196] | 0.186*<br>[-0.032 - 0.405] | -0.111<br>[-0.284 - 0.062] | -0.053<br>[-0.196 - 0.090] |
| Female labour part. | 0.203***<br>[0.134 - 0.272] | 0.523***<br>[0.137 - 0.910] | 0.248<br>[-0.492 - 0.988] | -0.246<br>[-0.863 - 0.370] |
| Observations | 1,290 | 1,290 | 1,290 | 1,290 |
| Number of countries |  | 90 | 90 | 90 |
| Country FE | NO | NO | YES | YES |
| Year FE | NO | NO | NO | YES |
| Overall R-squared | 0.119 | 0.0747 | 0.0185 | 0.394 |
| Within R-squared |  | 0.609 | 0.749 | 0.848 |
| Rho statistic |  | 0.607 | 0.995 | 0.861 |
| LM test |  | 576.13*** |  |  |
| Hausmann Chi2 |  |  | 1758.69*** |  |
| Testparm i. Year |  |  |  | 9.09*** |

Note. For fixed-effect estimators, standard errors are clustered at the country level to control for serial correlation. \*\*\* p<0.01, \*\* p<0.05, \* p<0.1 and 95 % CI in brackets. LM test; Breusch-Pagan Lagrange multiplier test. Data Sources: OECD-CRS, WDI, WGI.

## Appendix D. Complementary Results

**Table D.1:** Effect of SRH ODApc on modern contraceptive prevalence (comparison of estimators)

| VARIABLES | Pooled OLS | Random Effects | Country<br>Fixed-Effects | Main Model 2A<br>Tow-way FE | Robust S<br>Estimator | Robust MM-Estimator | Cooks' D<br>Removed |
| --- | --- | --- | --- | --- | --- | --- | --- |
| SRH ODA pc | 0.226**<br>[0.048 - 0.403] | 0.604***<br>[0.371 - 0.837] | 0.627***<br>[0.352 - 0.903] | 0.587***<br>[0.278 - 0.896] | 0.367***<br>[0.236 - 0.499] | 0.357<br>[-0.073 - 0.787] | 0.626***<br>[0.393 - 0.859] |
| ln GDPpc | 7.925***<br>[5.100 - 10.750] | 2.877**<br>[0.141 - 5.614] | -0.848<br>[-4.871 - 3.176] | 0.188<br>[-6.640 - 7.016] | 3.150<br>[-5.144 - 11.444] | 5.522<br>[-16.776 - 27.820] | 0.740<br>[-4.922 - 6.403] |
| GHE/CHE (%) | 0.230***<br>[0.097 - 0.363] | 0.056<br>[-0.060 - 0.171] | 0.038<br>[-0.080 - 0.157] | 0.052<br>[-0.067 - 0.171] | 0.068<br>[-0.019 - 0.154] | 0.061<br>[-0.086 - 0.208] | 0.120***<br>[0.036 - 0.203] |
| Government effective | 10.915***<br>[5.130 - 16.700] | 3.419<br>[-0.674 - 7.512] | 4.395**<br>[0.035 - 8.755] | 3.920*<br>[-0.335 - 8.175] | -1.344<br>[-7.116 - 4.428] | 1.904<br>[-4.906 - 8.714] | 2.629*<br>[-0.447 - 5.705] |
| Corruption control | -6.983***<br>[-11.597 - -2.368] | 0.656<br>[-3.623 - 4.935] | 1.815<br>[-3.328 - 6.957] | 2.300<br>[-3.093 - 7.693] | -0.671<br>[-5.251 - 3.910] | -0.422<br>[-9.559 - 8.716] | -0.150<br>[-4.005 - 3.705] |
| Adolescent fertility | -0.044<br>[-0.096 - 0.009] | -0.188***<br>[-0.273 - -0.103] | -0.208**<br>[-0.371 - -0.044] | -0.200**<br>[-0.377 - -0.024] | -0.250***<br>[-0.379 - -0.121] | -0.205<br>[-1.224 - 0.813] | -0.154*<br>[-0.313 - 0.004] |
| ln Population density | 1.381*<br>[-0.228 - 2.989] | 1.660<br>[-1.391 - 4.711] | 10.190<br>[-4.669 - 25.048] | 12.260<br>[-6.881 - 31.402] | 13.590<br>[-13.199 - 40.378] | 17.009<br>[-11.380 - 45.399] | 12.082*<br>[-1.684 - 25.848] |
| School enrolment | 0.309***<br>[0.218 - 0.400] | 0.092***<br>[0.027 - 0.158] | 0.049<br>[-0.021 - 0.120] | 0.060<br>[-0.012 - 0.132] | 0.015<br>[-0.057 - 0.086] | 0.018<br>[-0.072 - 0.108] | 0.049<br>[-0.016 - 0.114] |
| Female labour | 0.060<br>[-0.023 - 0.142] | -0.050<br>[-0.191 - 0.092] | -0.220<br>[-0.519 - 0.079] | -0.224<br>[-0.552 - 0.104] | -0.030<br>[-0.252 - 0.193] | -0.112<br>[-0.490 - 0.265] | -0.117<br>[-0.322 - 0.087] |
| Constant | -74.821***<br>[-105.979 - -43.663] | 11.393<br>[-22.056 - 44.842] | 23.748<br>[-39.639 - 87.135] | 5.091<br>[-109.949 - 120.132] | 4.159***<br>[3.157 - 5.161] |  | -11.112<br>[-88.588 - 66.363] |

Table D.1: Continued
| VARIABLES | Pooled OLS | Random Effects | Country<br>Fixed-Effects | Main Model 2A<br>Two-way FE | Robust S<br>Estimator | Robust MM-Estimator | Cooks' D<br>Removed |
| --- | --- | --- | --- | --- | --- | --- | --- |
| Observations | 382 | 382 | 382 | 382 | 376 | 376 | 355 |
| R-squared | 0.521 | 0.384 | 0.449 | 0.476 |  |  | 0.539 |
| Number of countries |  | 95 | 95 | 95 | 89 | 89 | 89 |
| Within R-squared |  | 0.428 | 0.449 | 0.476 |  |  | 0.539 |
| Country FE |  |  | YES | YES | YES | YES | YES |
| Year FE |  |  | NO | YES | YES | YES | YES |
| Lagrange multiplier p |  | 0.000 |  |  |  |  |  |
| Hausman chi2 p |  |  | 0.7373 |  |  |  |  |
| Hausman S vs. FE p |  |  |  |  | 0.00520 |  |  |
| Hausman MM vs. S p |  |  |  |  |  | 0.967 |  |

**Note.** Table C1 presents regression estimates for the effect of SRH on mCP from different estimators. Diagnostics for choosing the preferred estimator are indicated. For fixed-effect models, standard errors are clustered at the country level.<sup>1</sup> Cooks' Distance outliers were defined as observations with Cooks' D > 4/(N-k-1), where N is the number of observations, k the number of predictors. \*\*\* p<0.01, \*\* p<0.05, \* p<0.1 and a 95 % CI in brackets. Lagrange multiplier p>0.05 indicates that differences between countries are not systematic and RE model could have been used.<sup>2</sup> **Data sources.** OECD-CRS, WDI, WGI.
<sup>1</sup> Breusch-Pagan Lagrange multiplier (LM) tests the null hypothesis that variances across observations in the panel is zero.
The null hypothesis of the Hausman chi2 test is that the preferred model is random effects, and it tests if country specific error terms are correlated with the regressor.

**Table D.2:** Effect of Total Health ODApc on modern contraceptive prevalence (comparison of estimators)

| VARIABLES | Pooled OLS | Random Effects | Country<br>Fixed-Effects | Main Model 2B<br>Tow-way FE | Robust S<br>Estimator | Robust MM-Estimator | Cooks' D<br>Removed |
| --- | --- | --- | --- | --- | --- | --- | --- |
| Total Health ODApc | -0.067<br>[-0.234 - 0.099] | 0.249**<br>[0.057 - 0.441] | 0.288***<br>[0.081 - 0.496] | 0.252**<br>[0.045 - 0.458] | 0.034<br>[-0.023 - 0.092] | 0.131<br>[-0.112 - 0.374] | 0.369***<br>[0.233 - 0.505] |
| Ln GDPpc | 7.965***<br>[5.102 - 10.829] | 3.323**<br>[0.268 - 6.378] | -0.155<br>[-4.779 - 4.469] | 0.541<br>[-6.601 - 7.683] | 4.702*<br>[-0.192 - 9.596] | 7.087<br>[-6.407 - 20.581] | 1.840<br>[-3.624 - 7.305] |
| GHE/CHE (%) | 0.203***<br>[0.070 - 0.336] | 0.066<br>[-0.059 - 0.192] | 0.069<br>[-0.066 - 0.204] | 0.091<br>[-0.045 - 0.228] | 0.050<br>[-0.017 - 0.117] | 0.086<br>[-0.058 - 0.230] | 0.111**<br>[0.024 - 0.198] |
| Government effective | 10.730***<br>[4.980 - 16.481] | 3.319<br>[-1.170 - 7.808] | 3.785<br>[-1.226 - 8.796] | 3.278<br>[-1.636 - 8.193] | -0.507<br>[-4.162 - 3.149] | 1.170<br>[-4.584 - 6.924] | 3.110**<br>[0.251 - 5.969] |
| Corruption control | -5.543**<br>[-10.554 - -<br>0.531] | 0.887<br>[-3.810 - 5.583] | 2.236<br>[-3.299 - 7.771] | 3.050<br>[-2.785 - 8.885] | -1.588<br>[-5.222 - 2.047] | 0.025<br>[-5.207 - 5.257] | 0.275<br>[-3.515 - 4.065] |
| Adolescent fertility | -0.034<br>[-0.086 - 0.018] | -0.183***<br>[-0.272 - -0.094] | -0.243***<br>[-0.418 - -0.068] | -0.237**<br>[-0.422 - -0.053] | -0.273***<br>[-0.389 - -0.157] | -0.259<br>[-0.891 - 0.372] | -0.171**<br>[-0.335 - -0.007] |
| Ln Population density | 1.028<br>[-0.570 - 2.627] | 1.049<br>[-1.980 - 4.078] | 5.622<br>[-9.866 - 21.110] | 8.996<br>[-12.032 -<br>30.025] | 17.784**<br>[1.942 - 33.625] | 15.322<br>[-14.172 -<br>44.815] | 10.419<br>[-3.708 - 24.547] |
| School enrolment | 0.342***<br>[0.251 - 0.433] | 0.088**<br>[0.018 - 0.158] | 0.042<br>[-0.032 - 0.116] | 0.045<br>[-0.030 - 0.120] | 0.005<br>[-0.072 - 0.083] | 0.012<br>[-0.077 - 0.102] | 0.024<br>[-0.043 - 0.092] |
| Female labour | 0.066<br>[-0.018 - 0.149] | -0.040<br>[-0.191 - 0.111] | -0.255<br>[-0.586 - 0.076] | -0.272<br>[-0.633 - 0.089] | -0.045<br>[-0.273 - 0.182] | -0.145<br>[-0.536 - 0.246] | -0.169<br>[-0.386 - 0.047] |
| Constant | -75.116***<br>[-106.635 - -<br>43.596] | 9.102<br>[-26.970 -<br>45.174] | 40.926<br>[-25.320 -<br>107.171] | 20.614<br>[-104.550 -<br>145.779] | 4.189***<br>[3.122 - 5.256] |  | -6.660<br>[-85.564 -<br>72.244] |

Table D.2: Continued
| VARIABLES | Pooled OLS | Random Effects | Country<br>Fixed-Effects | Main Model 2B<br>Tow-way FE | Robust S<br>Estimator | Robust MM-Estimator | Cooks' D<br>Removed |
| --- | --- | --- | --- | --- | --- | --- | --- |
| Observations | 384 | 384 | 384 | 384 | 378 | 378 | 358 |
| R-squared | 0.512 | 0.366 | 0.411 | 0.443 |  |  | 0.546 |
| Number of countries |  | 96 | 96 | 96 | 90 | 90 | 91 |
| Within R-squared |  | 0.388 | 0.411 | 0.443 |  |  | 0.546 |
| Country FE |  |  | YES | YES | YES | YES | YES |
| Year FE |  |  |  | YES | YES | YES | YES |
| Lagrange multiplier p |  | 0.000 |  |  |  |  |  |
| Hausman chi2 p |  |  | 0.000 |  |  |  |  |
| Hausman S vs. FE p |  |  |  |  | 0.00364 |  |  |
| Hausman MM vs. S p |  |  |  |  |  | 0.0936 |  |

**Note.** Table C2 presents regression estimates for the effect of TotH ODA on mCP from different estimators. Diagnostics for choosing the preferred estimator are indicated. For fixed effect models, standard errors are clustered at the country level to control for serial correlation. Cooks' Distance outliers were defined as observations with Cooks' $D > 4/N-k-1$ , where N is the number of observations, k the number of predictors. \*\*\* $p < 0.01$ , \*\* $p < 0.05$ , \* $p < 0.1$ and a 95 % CI in brackets. Of the 96 countries in the FE model, 6 were dropped due to singleton observation. **Data sources.** OECD-CRS, WDI, WGI.

**Table D.3:** Effect of RH care ODApc on modern contraceptive prevalence (comparison of estimators)

| VARIABLES | Pooled OLS | Random Effects | Country<br>Fixed-Effects | Main Model 2C<br>Tow-way FE | Robust S<br>Estimator | Robust MM-Estimator | Cooks' D<br>Removed |
| --- | --- | --- | --- | --- | --- | --- | --- |
| RH Health ODA pc | -1.163<br>[-2.912 - 0.587] | 0.846<br>[-1.269 - 2.962] | 0.855<br>[-1.397 - 3.108] | 0.759<br>[-1.509 - 3.027] | 0.363<br>[-0.419 - 1.146] | 1.116<br>[-0.298 - 2.530] | 1.591**<br>[0.198 - 2.985] |
| Ln GDPpc | 7.755***<br>[4.903 - 10.606] | 4.096**<br>[0.891 - 7.301] | 0.186<br>[-5.465 - 5.838] | -1.533<br>[-9.314 - 6.248] | 3.768<br>[-4.246 - 11.783] | 5.779<br>[-5.327 - 16.885] | -0.601<br>[-7.309 - 6.106] |
| GHE/CHE (%) | 0.215***<br>[0.081 - 0.349] | 0.052<br>[-0.069 - 0.174] | 0.041<br>[-0.086 - 0.168] | 0.066<br>[-0.065 - 0.198] | 0.043<br>[-0.037 - 0.124] | 0.075<br>[-0.050 - 0.200] | 0.110***<br>[0.029 - 0.190] |
| Government effective | 10.545***<br>[4.777 - 16.314] | 3.418<br>[-1.982 - 8.818] | 4.666<br>[-1.713 - 11.045] | 4.004<br>[-1.890 - 9.898] | -0.578<br>[-7.583 - 6.426] | 1.807<br>[-3.710 - 7.325] | 3.403**<br>[0.169 - 6.637] |
| Corruption control | -5.418**<br>[-10.299 - -<br>0.536] | 1.320<br>[-4.044 - 6.685] | 2.305<br>[-3.731 - 8.340] | 3.730<br>[-2.634 - 10.095] | -1.355<br>[-6.087 - 3.376] | 0.606<br>[-4.399 - 5.610] | 1.183<br>[-2.786 - 5.152] |
| Adolescent fertility | -0.040<br>[-0.092 - 0.011] | -0.184***<br>[-0.274 - -0.093] | -0.223**<br>[-0.405 - -0.040] | -0.223**<br>[-0.409 - -0.037] | -0.280***<br>[-0.455 - -0.105] | -0.263<br>[-0.746 - 0.220] | -0.171*<br>[-0.350 - 0.009] |
| ln Population density | 1.099<br>[-0.495 - 2.694] | 1.165<br>[-1.836 - 4.165] | 10.202<br>[-7.087 - 27.492] | 10.101<br>[-13.431 -<br>33.633] | 15.964<br>[-21.759 -<br>53.687] | 17.169<br>[-10.870 -<br>45.207] | 13.442*<br>[-1.027 - 27.910] |
| School enrolment | 0.338***<br>[0.251 - 0.425] | 0.107***<br>[0.034 - 0.180] | 0.047<br>[-0.031 - 0.125] | 0.045<br>[-0.034 - 0.125] | 0.004<br>[-0.105 - 0.112] | 0.004<br>[-0.077 - 0.086] | 0.022<br>[-0.048 - 0.092] |
| Female labour | 0.053<br>[-0.032 - 0.139] | -0.023<br>[-0.183 - 0.136] | -0.280<br>[-0.673 - 0.113] | -0.311<br>[-0.728 - 0.106] | -0.042<br>[-0.272 - 0.189] | -0.125<br>[-0.476 - 0.227] | -0.186<br>[-0.421 - 0.049] |
| Constant | -72.318***<br>[-103.966 - -<br>40.670] | 2.487<br>[-34.384 -<br>39.358] | 22.084<br>[-45.238 -<br>89.405] | 35.922<br>[-103.701 -<br>175.546] | 4.265***<br>[3.160 - 5.369] |  | 3.540<br>[-82.243 -<br>89.323] |

Table D.3: Continued
| *VARIABLES* | *Pooled OLS* | *Random Effects* | *Fixed Effects* | *Fixed Effects*<br>*Main Model 2C* | *Robust S*<br>*Estimator* | *Robust MM-*<br>*Estimator* | *Cooks'D*<br>*removed* |
| --- | --- | --- | --- | --- | --- | --- | --- |
| Observations | 378 | 378 | 378 | 378 | 372 | 372 | 354 |
| R-squared | 0.526 | 0.405 | 0.365 | 0.414 |  |  | 0.501 |
| Number of countries |  | 94 | 94 | 94 | 88 | 88 | 90 |
| Within R-squared |  | 0.334 | 0.365 | 0.414 |  |  | 0.501 |
| Country FE |  |  | YES | YES | YES | YES | YES |
| Year FE |  |  |  | YES | YES | YES | YES |
| Lagrange multiplier p |  | 0.000 |  |  |  |  |  |
| Hausman chi2 p |  |  | 0.000 |  |  |  |  |
| Hausman S vs. FE p |  |  |  |  | 0.000373 |  |  |
| Hausman MM vs. S p |  |  |  |  |  | 0.255 |  |

**Note.** Table C3 presents regression estimates for the effect of RH ODA on mCP from different estimators. Diagnostics for choosing the preferred estimator are indicated. For fixed effect models, standard errors are clustered at the country level to control for serial correlation. Cooks' Distance outliers were defined as observations with Cooks' $D > 4/N-k-1$ , where N is the number of observations, k the number of predictors. Of the 94 countries in the FE model, 6 do not contribute to estimates due to singleton observation leaving 88 countries. \*\*\* p<0.01, \*\* p<0.05, \* p<0.1 and a 95 % CI in brackets. **Data sources.** OECD-CRS, WDI, WGI.

**Table D.4:** Effect of SRH ODApc on skilled birth attendance (comparison of estimators)

| VARIABLES | Pooled OLS | Random Effects | Country<br>Fixed-Effects | Main Model 2A<br>Tow-way FE | Robust S<br>Estimator | Robust MM-Estimator | Cooks' D<br>Removed |
| --- | --- | --- | --- | --- | --- | --- | --- |
| SRH Health ODA pc | 0.139*<br>[-0.017 - 0.295] | 0.140<br>[-0.056 - 0.336] | 0.075<br>[-0.132 - 0.283] | 0.185*<br>[-0.022 - 0.392] | 0.295***<br>[0.215 - 0.375] | 0.136<br>[-0.040 - 0.313] | 0.215**<br>[0.039 - 0.391] |
| Ln GDPpc | 13.298***<br>[11.028 - 15.569] | 12.548***<br>[8.590 - 16.506] | 6.019**<br>[0.952 - 11.086] | 11.114***<br>[2.730 - 19.497] | 0.921<br>[-3.446 - 5.289] | 2.564<br>[-2.736 - 7.865] | 9.048***<br>[3.382 - 14.713] |
| GHE/CHE (%) | 0.136***<br>[0.053 - 0.219] | 0.081<br>[-0.020 - 0.181] | 0.085<br>[-0.020 - 0.191] | 0.067<br>[-0.042 - 0.175] | 0.018<br>[-0.015 - 0.052] | 0.076<br>[-0.026 - 0.179] | 0.064<br>[-0.021 - 0.148] |
| Government effective | 2.335<br>[-1.953 - 6.624] | -3.176<br>[-7.588 - 1.237] | -0.583<br>[-5.282 - 4.115] | -2.013<br>[-6.771 - 2.745] | -0.587<br>[-1.700 - 0.526] | -0.086<br>[-4.605 - 4.433] | -2.218<br>[-6.414 - 1.979] |
| Corruption control | -2.235<br>[-5.713 - 1.243] | 2.177<br>[-3.188 - 7.542] | 3.104<br>[-2.667 - 8.874] | 3.415<br>[-2.088 - 8.918] | 0.438<br>[-0.673 - 1.549] | 2.316<br>[-1.082 - 5.715] | 4.425*<br>[-0.101 - 8.950] |
| Adolescent fertility | -0.180***<br>[-0.219 - -0.140] | -0.301***<br>[-0.375 - -0.227] | -0.354***<br>[-0.511 - -0.197] | -0.351***<br>[-0.491 - -0.211] | -0.023<br>[-0.136 - 0.091] | -0.307***<br>[-0.506 - -0.109] | -0.346***<br>[-0.481 - -0.212] |
| lnPopulation density | -0.256<br>[-1.236 - 0.725] | 0.956<br>[-1.626 - 3.538] | 26.809***<br>[8.508 - 45.110] | 29.730***<br>[8.794 - 50.666] | 4.312<br>[-13.656 - 22.280] | 35.708***<br>[19.565 - 51.852] | 35.825***<br>[22.387 - 49.263] |
| School enrolment | 0.141***<br>[0.046 - 0.235] | 0.114<br>[-0.030 - 0.258] | 0.072<br>[-0.091 - 0.236] | 0.075<br>[-0.090 - 0.240] | 0.008<br>[-0.027 - 0.043] | 0.013<br>[-0.133 - 0.160] | 0.005<br>[-0.105 - 0.116] |
| Female labour | 0.052<br>[-0.038 - 0.142] | 0.110<br>[-0.095 - 0.314] | -0.021<br>[-0.469 - 0.427] | -0.033<br>[-0.468 - 0.403] | 0.059<br>[-0.047 - 0.166] | -0.081<br>[-0.400 - 0.237] | -0.026<br>[-0.436 - 0.384] |
| Constant | -44.934***<br>[-69.581 - -20.286] | -34.565<br>[-76.293 - 7.163] | -66.475<br>[-146.763 - 13.813] | -115.472*<br>[-239.548 - 8.605] | 3.772***<br>[1.516 - 6.029] |  | -113.356***<br>[-184.722 - -41.990] |

Table D.4: Continued
| *VARIABLES* | *Pooled OLS* | *Random Effects* | *Country*<br>*Fixed-Effects* | *Main Model 2A*<br>*Tow-way FE* | *Robust S*<br>*Estimator* | *Robust MM-*<br>*Estimator* | *Cooks' D*<br>*Removed* |
| --- | --- | --- | --- | --- | --- | --- | --- |
| Observations | 661 | 661 | 661 | 661 | 657 | 657 | 592 |
| R-squared | 0.599 | 0.574 | 0.471 | 0.510 |  |  | 0.616 |
| Number of countries |  | 101 | 101 | 101 | 97 | 97 | 89 |
| Within R-squared |  | 0.430 | 0.471 | 0.510 |  |  | 0.616 |
| Country FE |  |  | YES | YES | YES | YES | YES |
| Year FE |  |  | NO | YES | YES | YES | YES |
| Lagrange multiplier p |  | 0.000 |  |  |  |  |  |
| Hausman chi2 p |  |  | 0.000 |  |  |  |  |
| Hausman S vs. FE p |  |  |  |  | 0.000 |  |  |
| Hausman MM vs. S p |  |  |  |  |  | 1.37e-07 |  |

**Note.** Table C4 presents regression estimates for the effect of SRH ODA on SBA from different estimators. Diagnostics for choosing the preferred estimator are indicated. For fixed effect models, standard errors are clustered at the country level to control for serial correlation. Cooks' Distance outliers were defined as observations with Cooks' D $> 4/N-k-1$ , where N is the number of observations, k the number of predictors. Of the 101 countries in the FE model, 4 do not contribute to estimates due to singleton observation leaving 97 countries. \*\*\* p<0.01, \*\* p<0.05, \* p<0.1 and a 95 % CI in brackets. **Data sources.** OECD-CRS, WDI, WGI.

**Table D.5:** Effect of Total health ODApc on skilled birth attendance (comparison of estimators)

| VARIABLES | Pooled OLS | Random Effects | Country<br>Fixed-Effects | Main Model 2B<br>Tow-way FE | Robust S<br>Estimator | Robust MM-Estimator | Cooks' D<br>Removed |
| --- | --- | --- | --- | --- | --- | --- | --- |
| Health ODA pc | 0.214***<br>[0.106 - 0.322] | 0.094*<br>[-0.008 - 0.196] | 0.041<br>[-0.032 - 0.114] | 0.073*<br>[-0.008 - 0.154] | -0.009<br>[-0.038 - 0.020] | 0.017<br>[-0.028 - 0.062] | 0.022<br>[-0.042 - 0.086] |
| Ln GDPpc | 14.055***<br>[11.701 - 16.409] | 12.356***<br>[8.462 - 16.249] | 5.857**<br>[0.921 - 10.794] | 11.414***<br>[3.126 - 19.702] | 1.456<br>[-2.195 - 5.108] | 2.848<br>[-2.181 - 7.878] | 9.461***<br>[3.881 - 15.041] |
| GHE/CHE (%) | 0.124***<br>[0.043 - 0.205] | 0.080<br>[-0.017 - 0.178] | 0.087<br>[-0.017 - 0.190] | 0.070<br>[-0.037 - 0.178] | 0.015<br>[-0.022 - 0.052] | 0.078<br>[-0.025 - 0.182] | 0.065<br>[-0.018 - 0.147] |
| Government effective | 2.612<br>[-1.572 - 6.797] | -2.763<br>[-7.078 - 1.552] | -0.306<br>[-4.782 - 4.169] | -1.571<br>[-6.126 - 2.984] | -0.233<br>[-1.402 - 0.936] | 0.103<br>[-3.816 - 4.023] | -1.548<br>[-5.518 - 2.421] |
| Corruption control | -2.909*<br>[-6.302 - 0.485] | 2.247<br>[-2.891 - 7.384] | 3.039<br>[-2.546 - 8.624] | 3.511<br>[-1.764 - 8.786] | 0.227<br>[-1.106 - 1.561] | 2.271<br>[-0.929 - 5.472] | 4.291**<br>[0.000 - 8.582] |
| Adolescent fertility | -0.179***<br>[-0.219 - -0.140] | -0.298***<br>[-0.373 - -0.224] | -0.354***<br>[-0.509 - -0.199] | -0.360***<br>[-0.500 - -0.220] | -0.028<br>[-0.131 - 0.074] | -0.315***<br>[-0.488 - -0.143] | -0.352***<br>[-0.482 - -0.221] |
| lnPopulation density | -0.145<br>[-1.099 - 0.810] | 0.815<br>[-1.674 - 3.303] | 26.786***<br>[8.632 - 44.940] | 30.645***<br>[10.072 - 51.217] | 5.750<br>[-6.439 - 17.939] | 36.992***<br>[21.702 - 52.281] | 38.772***<br>[26.022 - 51.522] |
| School enrolment | 0.123**<br>[0.029 - 0.217] | 0.110<br>[-0.032 - 0.253] | 0.071<br>[-0.091 - 0.232] | 0.072<br>[-0.091 - 0.235] | 0.017<br>[-0.009 - 0.042] | 0.020<br>[-0.121 - 0.161] | -0.005<br>[-0.112 - 0.103] |
| Female labour | 0.064<br>[-0.025 - 0.153] | 0.108<br>[-0.095 - 0.311] | -0.022<br>[-0.471 - 0.426] | -0.028<br>[-0.460 - 0.405] | 0.069<br>[-0.047 - 0.184] | -0.076<br>[-0.401 - 0.249] | -0.015<br>[-0.417 - 0.387] |
| Constant | -51.542***<br>[-76.823 - -26.262] | -32.128<br>[-73.271 - 9.016] | -65.094<br>[-145.871 - 15.682] | -121.245*<br>[-243.113 - 0.624] | 3.736***<br>[1.554 - 5.918] |  | -127.238***<br>[-196.073 - -58.403] |

Table D.5: Continued
| VARIABLES | Pooled OLS | Random Effects | Country<br>Fixed-Effects | Main Model 2B<br>Two-way FE | Robust S<br>Estimator | Robust MM-Estimator | Cooks' D<br>Removed |
| --- | --- | --- | --- | --- | --- | --- | --- |
| Observations | 670 | 670 | 670 | 670 | 666 | 666 | 605 |
| R-squared | 0.606 | 0.579 | 0.467 | 0.507 |  |  | 0.882 |
| Number of countries |  | 101 | 101 | 101 | 97 | 97 | 89 |
| Within R-squared |  | 0.425 | 0.467 | 0.507 |  |  | 0.882 |
| Country FE |  |  | YES | YES | YES | YES | YES |
| Year FE |  |  | NO | YES | YES | YES | YES |
| Lagrange multiplier p |  | 0.000 |  |  |  |  |  |
| Hausman chi2 p |  |  | 0.000 |  |  |  |  |
| Hausman S vs. FE p |  |  |  |  | 0.000 |  |  |
| Hausman MM vs. S p |  |  |  |  |  | 0.000 |  |

**Table D.6:** Effect of RH care ODApc on skilled birth attendance (comparison of estimators)

| VARIABLES | Pooled OLS | Random Effects | Country<br>Fixed-Effects | Main Model 2C<br>Tow-way FE | Robust S<br>Estimator | Robust MM-Estimator | Cooks' D<br>Removed |
| --- | --- | --- | --- | --- | --- | --- | --- |
| RH Health ODA pc | -0.527<br>[-2.477 - 1.423] | 1.271<br>[-0.578 - 3.120] | 0.985<br>[-0.622 - 2.591] | 1.039<br>[-0.402 - 2.480] | 0.421<br>[-0.136 - 0.977] | 0.551<br>[-0.803 - 1.905] | 0.482<br>[-0.545 - 1.509] |
| Ln GDPpc | 13.473***<br>[11.047 - 15.899] | 12.931***<br>[8.935 - 16.927] | 6.318**<br>[1.091 - 11.544] | 9.835**<br>[1.368 - 18.302] | 1.372<br>[-3.342 - 6.087] | 1.554<br>[-4.553 - 7.660] | 9.043***<br>[3.294 - 14.792] |
| GHE/CHE (%) | 0.116***<br>[0.029 - 0.202] | 0.095*<br>[-0.005 - 0.195] | 0.105*<br>[-0.004 - 0.213] | 0.089<br>[-0.022 - 0.200] | 0.021<br>[-0.025 - 0.067] | 0.086*<br>[-0.013 - 0.185] | 0.077*<br>[-0.007 - 0.160] |
| Government effective | 1.971<br>[-2.337 - 6.278] | -3.076<br>[-7.593 - 1.440] | -0.112<br>[-4.889 - 4.665] | -1.500<br>[-6.259 - 3.259] | -0.439<br>[-1.924 - 1.046] | 0.283<br>[-3.998 - 4.564] | -1.681<br>[-5.593 - 2.232] |
| Corruption control | -1.534<br>[-5.091 - 2.023] | 2.673<br>[-3.007 - 8.354] | 3.598<br>[-2.456 - 9.652] | 3.938<br>[-1.763 - 9.640] | 0.504<br>[-0.851 - 1.860] | 2.700<br>[-0.698 - 6.099] | 4.741**<br>[0.125 - 9.357] |
| Adolescent fertility | -0.173***<br>[-0.214 - -0.133] | -0.297***<br>[-0.370 - -0.223] | -0.365***<br>[-0.526 - -0.205] | -0.366***<br>[-0.512 - -0.219] | -0.045<br>[-0.199 - 0.108] | -0.317***<br>[-0.502 - -0.131] | -0.367***<br>[-0.498 - -0.236] |
| ln Population density | -0.455<br>[-1.468 - 0.557] | 0.774<br>[-1.734 - 3.283] | 25.430***<br>[7.022 - 43.838] | 26.443**<br>[6.039 - 46.847] | 5.227<br>[-7.159 - 17.613] | 36.078***<br>[20.357 - 51.799] | 32.192***<br>[16.526 - 47.857] |
| School enrolment | 0.153***<br>[0.059 - 0.247] | 0.113<br>[-0.032 - 0.258] | 0.067<br>[-0.098 - 0.232] | 0.074<br>[-0.093 - 0.241] | 0.018<br>[-0.013 - 0.048] | 0.008<br>[-0.153 - 0.168] | 0.011<br>[-0.102 - 0.125] |
| Female labour | 0.053<br>[-0.040 - 0.145] | 0.117<br>[-0.085 - 0.319] | -0.047<br>[-0.506 - 0.411] | -0.069<br>[-0.513 - 0.376] | 0.071<br>[-0.068 - 0.210] | -0.081<br>[-0.403 - 0.242] | -0.077<br>[-0.471 - 0.318] |
| Constant | -45.799***<br>[-72.060 - -19.538] | -38.054*<br>[-80.140 - 4.032] | -61.311<br>[-141.546 - 18.924] | -89.502<br>[-208.355 - 29.351] | 3.933***<br>[1.563 - 6.304] |  | -94.469**<br>[-173.057 - -15.881] |

Table D.6: Continued
| VARIABLES | Pooled OLS | Random Effects | Country<br>Fixed-Effects | Main Model 2C<br>Two-way FE | Robust S<br>Estimator | Robust MM-Estimator | Cooks' D<br>Removed |
| --- | --- | --- | --- | --- | --- | --- | --- |
| Observations | 640 | 640 | 640 | 640 | 635 | 635 | 582 |
| R-squared | 0.596 | 0.570 | 0.488 | 0.524 |  |  | 0.615 |
| Number of countries |  | 101 | 101 | 101 | 96 | 96 | 97 |
| Within R-squared |  | 0.449 | 0.488 | 0.524 |  |  | 0.615c |
| Country FE |  |  | YES | YES | YES | YES | YES |
| Year FE |  |  | NO | YES | YES | YES | YES |
| Lagrange multiplier p |  | 0.000 |  |  |  |  |  |
| Hausman chi2 p |  |  | 0.000 |  |  |  |  |
| Hausman S vs. FE p |  |  |  |  | 0.000 |  |  |
| Hausman MM vs. S p |  |  |  |  |  | 0.000 |  |

**Table D.7:** Effect of SRH ODApc on ART coverage (comparison of estimators)

| VARIABLES | Pooled OLS | Random Effects | Country<br>Fixed-Effects | Main Model 2A<br>Tow-way FE | Robust S<br>Estimator | Robust MM-Estimator | Cooks' D<br>Removed |
| --- | --- | --- | --- | --- | --- | --- | --- |
| SRH ODA pc | 0.753***<br>[0.596 - 0.910] | 0.849***<br>[0.567 - 1.131] | 0.604***<br>[0.333 - 0.875] | 0.573***<br>[0.412 - 0.734] | 0.519***<br>[0.400 - 0.637] | 0.586***<br>[0.384 - 0.789] | 0.635***<br>[0.489 - 0.781] |
| ln GDPpc | 8.329***<br>[6.560 - 10.098] | 32.848***<br>[26.431 - 39.265] | 28.341***<br>[20.435 - 36.246] | -2.905<br>[-14.683 - 8.873] | -12.794**<br>[-25.583 - -<br>0.005] | -5.621<br>[-56.529 -<br>45.287] | -6.710<br>[-16.045 - 2.626] |
| GHE/CHE (%) | -0.016<br>[-0.097 - 0.064] | -0.040<br>[-0.239 - 0.160] | 0.081<br>[-0.116 - 0.277] | -0.026<br>[-0.164 - 0.111] | 0.092<br>[-0.069 - 0.253] | 0.040<br>[-0.177 - 0.257] | 0.009<br>[-0.099 - 0.116] |
| Government effective | -1.605<br>[-4.951 - 1.741] | -1.837<br>[-8.681 - 5.007] | 10.847***<br>[4.699 - 16.994] | 6.456**<br>[0.992 - 11.919] | -1.288<br>[-6.323 - 3.747] | 3.962<br>[-3.232 - 11.157] | 6.200***<br>[1.591 - 10.809] |
| Control of corruption | 2.856<br>[-0.565 - 6.276] | 0.890<br>[-6.762 - 8.541] | 2.178<br>[-4.147 - 8.503] | 3.715<br>[-3.097 - 10.527] | -0.136<br>[-5.299 - 5.026] | 3.135<br>[-4.029 - 10.299] | 3.742<br>[-1.312 - 8.795] |
| School enrolment | 0.009<br>[-0.022 - 0.041] | -0.052<br>[-0.214 - 0.110] | -0.081<br>[-0.451 - 0.288] | -0.005<br>[-0.275 - 0.265] | -0.130<br>[-0.333 - 0.073] | -0.162<br>[-0.918 - 0.594] | -0.023<br>[-0.197 - 0.152] |
| ln Population density | 2.501***<br>[1.604 - 3.399] | 10.964***<br>[6.395 - 15.534] | 72.051***<br>[40.902 -<br>103.200] | -0.082<br>[-27.318 -<br>27.155] | -4.374<br>[-47.981 -<br>39.232] | -11.294<br>[-57.723 -<br>35.135] | -2.602<br>[-22.031 -<br>16.827] |
| Female labour | 0.079**<br>[0.016 - 0.142] | 0.075<br>[-0.115 - 0.266] | -0.090<br>[-0.252 - 0.073] | -0.009<br>[-0.141 - 0.123] | 0.041<br>[-0.107 - 0.189] | 0.057<br>[-0.136 - 0.249] | 0.046<br>[-0.051 - 0.143] |
| Adolescent fertility | 0.221***<br>[0.156 - 0.286] | 0.649***<br>[0.313 - 0.985] | 0.220<br>[-0.455 - 0.895] | -0.090<br>[-0.631 - 0.452] | -0.318**<br>[-0.622 - -0.014] | -0.129<br>[-0.716 - 0.459] | -0.035<br>[-0.403 - 0.332] |
| Constant | -79.971***<br>[-96.490 - -<br>63.453] | -335.955***<br>[-402.593 - -<br>269.317] | -493.162***<br>[-617.621 - -<br>368.703] | 36.432<br>[-126.156 -<br>199.020] | 6.970***<br>[5.922 - 8.018] |  | 68.482<br>[-51.422 -<br>188.386] |

Table D.7: Continued
| VARIABLES | Pooled OLS | Random Effects | Country<br>Fixed-Effects | Main Model 2A<br>Two-way FE | Robust S<br>Estimator | Robust MM-Estimator | Cooks' D<br>Removed |
| --- | --- | --- | --- | --- | --- | --- | --- |
| Observations | 1,295 | 1,295 | 1,295 | 1,295 | 1,295 | 1,295 | 1,225 |
| R-squared | 0.227 | 0.157 | 0.740 | 0.850 |  |  | 0.895 |
| Number of countries |  | 90 | 90 | 90 | 90 | 90 | 90 |
| Within R-squared |  | 0.654 | 0.740 | 0.850 |  |  | 0.895 |
| Country FE |  |  | YES | YES | YES | YES | YES |
| Year FE |  |  | NO | YES | YES | YES | YES |
| Lagrange multiplier p |  | 0.000 |  |  |  |  |  |
| Hausman chi2 p |  |  | 0.000 |  |  |  |  |
| Hausman S vs. FE p |  |  |  |  | 0.00175 |  |  |
| Hausman MM vs. S p |  |  |  |  |  | 0.0125 |  |

**Note.** Table C7 presents regression estimates for the effect of SRH ODA on ART coverage from different estimators. Diagnostics for choosing the preferred estimator are indicated. For fixed effect models, standard errors are clustered at the country level to control for serial correlation. Cooks' Distance outliers were defined as observations with Cooks' $D > 4/N-k-1$ , where N is the number of observations, k the number of predictors. \*\*\* $p < 0.01$ , \*\* $p < 0.05$ , \* $p < 0.1$ and a 95 % CI in brackets. **Data sources.** OECD-CRS, WDI, WGI.

**Table D.8:**
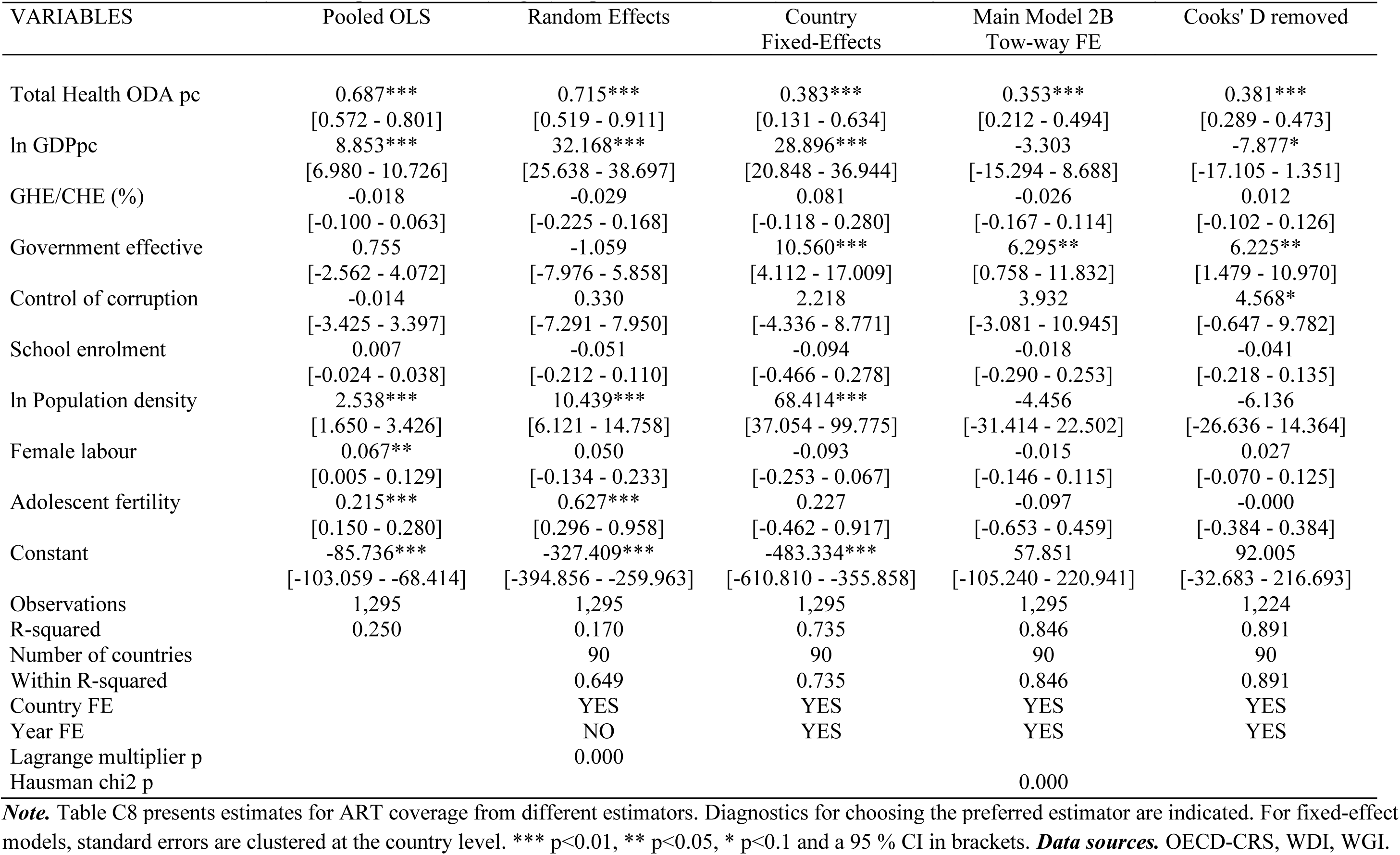
Effect of Total Health ODApc on ART coverage (comparison of estimators)

**Table D.9:**
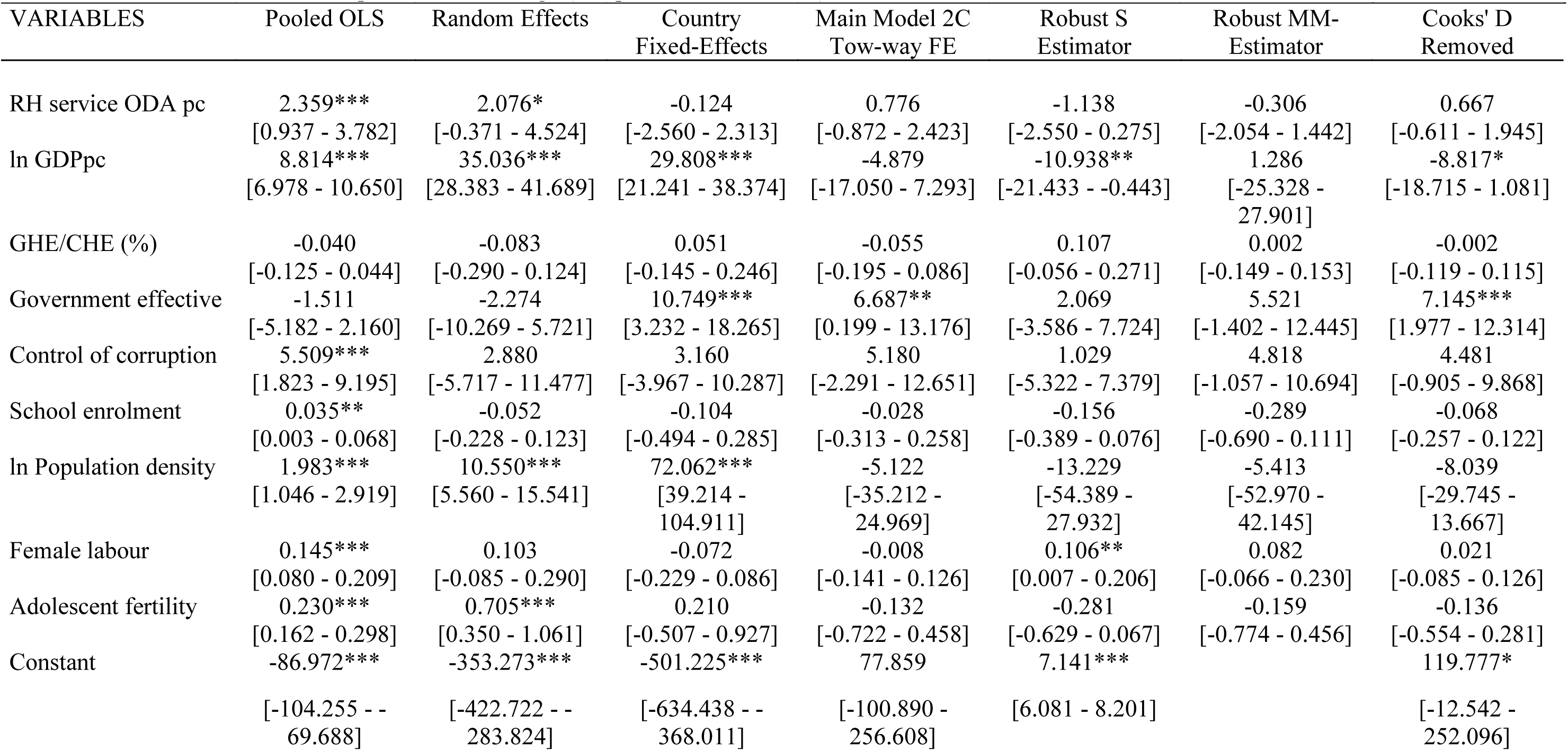
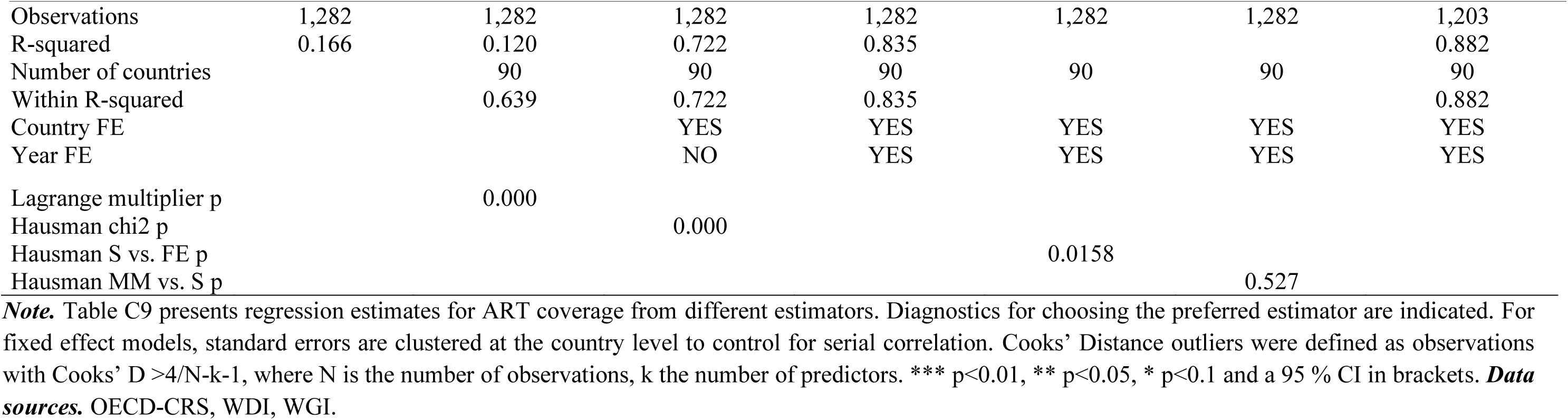
Effect of RH care ODApc on ART coverage (comparison of estimators)

| VARIABLES | Pooled OLS | Random Effects | Country<br>Fixed-Effects | Main Model 2C<br>Tow-way FE | Robust S<br>Estimator | Robust MM-Estimator | Cooks' D<br>Removed |
| --- | --- | --- | --- | --- | --- | --- | --- |
| RH service ODA pc | 2.359***<br>[0.937 - 3.782] | 2.076*<br>[-0.371 - 4.524] | -0.124<br>[-2.560 - 2.313] | 0.776<br>[-0.872 - 2.423] | -1.138<br>[-2.550 - 0.275] | -0.306<br>[-2.054 - 1.442] | 0.667<br>[-0.611 - 1.945] |
| ln GDPpc | 8.814***<br>[6.978 - 10.650] | 35.036***<br>[28.383 - 41.689] | 29.808***<br>[21.241 - 38.374] | -4.879<br>[-17.050 - 7.293] | -10.938**<br>[-21.433 - -0.443] | 1.286<br>[-25.328 - 27.901] | -8.817*<br>[-18.715 - 1.081] |
| GHE/CHE (%) | -0.040<br>[-0.125 - 0.044] | -0.083<br>[-0.290 - 0.124] | 0.051<br>[-0.145 - 0.246] | -0.055<br>[-0.195 - 0.086] | 0.107<br>[-0.056 - 0.271] | 0.002<br>[-0.149 - 0.153] | -0.002<br>[-0.119 - 0.115] |
| Government effective | -1.511<br>[-5.182 - 2.160] | -2.274<br>[-10.269 - 5.721] | 10.749***<br>[3.232 - 18.265] | 6.687**<br>[0.199 - 13.176] | 2.069<br>[-3.586 - 7.724] | 5.521<br>[-1.402 - 12.445] | 7.145***<br>[1.977 - 12.314] |
| Control of corruption | 5.509***<br>[1.823 - 9.195] | 2.880<br>[-5.717 - 11.477] | 3.160<br>[-3.967 - 10.287] | 5.180<br>[-2.291 - 12.651] | 1.029<br>[-5.322 - 7.379] | 4.818<br>[-1.057 - 10.694] | 4.481<br>[-0.905 - 9.868] |
| School enrolment | 0.035**<br>[0.003 - 0.068] | -0.052<br>[-0.228 - 0.123] | -0.104<br>[-0.494 - 0.285] | -0.028<br>[-0.313 - 0.258] | -0.156<br>[-0.389 - 0.076] | -0.289<br>[-0.690 - 0.111] | -0.068<br>[-0.257 - 0.122] |
| ln Population density | 1.983***<br>[1.046 - 2.919] | 10.550***<br>[5.560 - 15.541] | 72.062***<br>[39.214 - 104.911] | -5.122<br>[-35.212 - 24.969] | -13.229<br>[-54.389 - 27.932] | -5.413<br>[-52.970 - 42.145] | -8.039<br>[-29.745 - 13.667] |
| Female labour | 0.145***<br>[0.080 - 0.209] | 0.103<br>[-0.085 - 0.290] | -0.072<br>[-0.229 - 0.086] | -0.008<br>[-0.141 - 0.126] | 0.106**<br>[0.007 - 0.206] | 0.082<br>[-0.066 - 0.230] | 0.021<br>[-0.085 - 0.126] |
| Adolescent fertility | 0.230***<br>[0.162 - 0.298] | 0.705***<br>[0.350 - 1.061] | 0.210<br>[-0.507 - 0.927] | -0.132<br>[-0.722 - 0.458] | -0.281<br>[-0.629 - 0.067] | -0.159<br>[-0.774 - 0.456] | -0.136<br>[-0.554 - 0.281] |
| Constant | -86.972***<br>[-104.255 - -69.688] | -353.273***<br>[-422.722 - -283.824] | -501.225***<br>[-634.438 - -368.011] | 77.859<br>[-100.890 - 256.608] | 7.141***<br>[6.081 - 8.201] |  | 119.777*<br>[-12.542 - 252.096] |

Table D.9: Continued
| VARIABLES | Pooled OLS | Random Effects | Country<br>Fixed-Effects | Main Model 2C<br>Tow-way FE | Robust S<br>Estimator | Robust MM-Estimator | Cooks' D<br>Removed |
| --- | --- | --- | --- | --- | --- | --- | --- |
| Observations | 1,282 | 1,282 | 1,282 | 1,282 | 1,282 | 1,282 | 1,203 |
| R-squared | 0.166 | 0.120 | 0.722 | 0.835 |  |  | 0.882 |
| Number of countries |  | 90 | 90 | 90 | 90 | 90 | 90 |
| Within R-squared |  | 0.639 | 0.722 | 0.835 |  |  | 0.882 |
| Country FE |  |  | YES | YES | YES | YES | YES |
| Year FE |  |  | NO | YES | YES | YES | YES |
| Lagrange multiplier p |  | 0.000 |  |  |  |  |  |
| Hausman chi2 p |  |  | 0.000 |  |  |  |  |
| Hausman S vs. FE p |  |  |  |  | 0.0158 |  |  |
| Hausman MM vs. S p |  |  |  |  |  | 0.527 |  |

**Note.** Table C9 presents regression estimates for ART coverage from different estimators. Diagnostics for choosing the preferred estimator are indicated. For fixed effect models, standard errors are clustered at the country level to control for serial correlation. Cooks' Distance outliers were defined as observations with Cooks' $D > 4/N-k-1$ , where N is the number of observations, k the number of predictors. \*\*\* $p < 0.01$ , \*\* $p < 0.05$ , \* $p < 0.1$ and a 95 % CI in brackets. **Data sources.** OECD-CRS, WDI, WGI.

**Table D.10:**
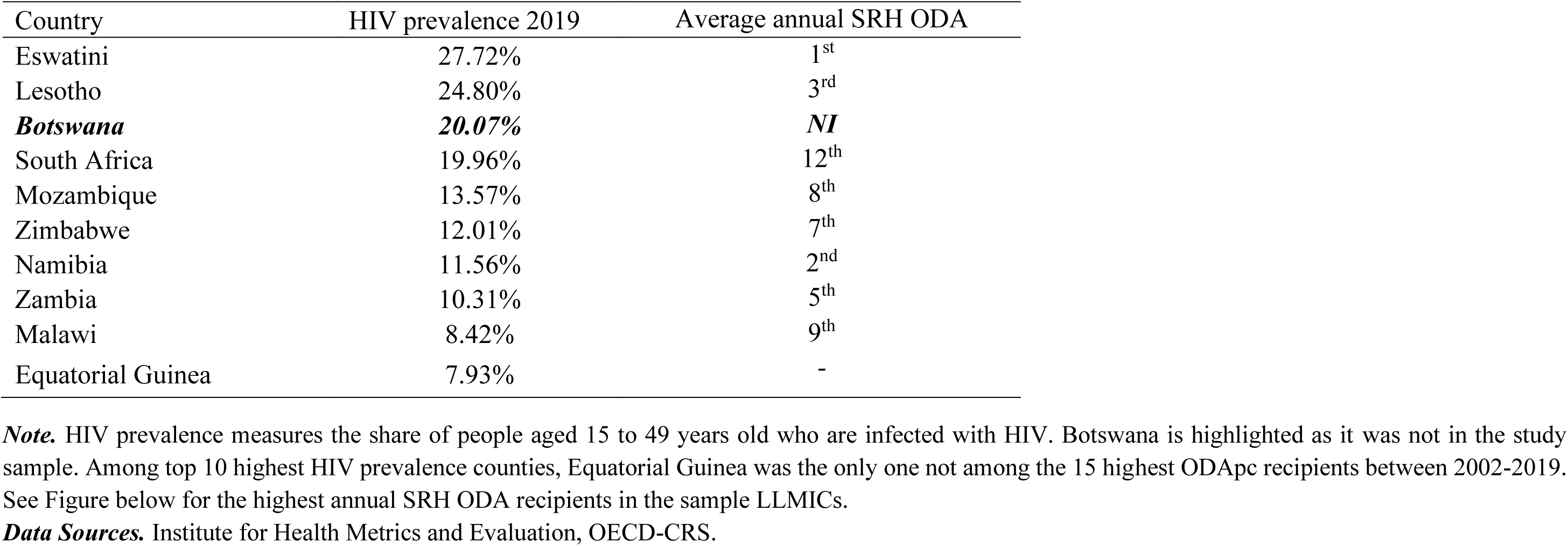
10 countries with highest HIV prevalence (in 2019) compared to highest average annual SRH ODA (2019, US$ pc) recipients in the sample LLMICs.

| Country | HIV prevalence 2019 | Average annual SRH ODA |
| --- | --- | --- |
| Eswatini | 27.72% | 1<sup>st</sup> |
| Lesotho | 24.80% | 3<sup>rd</sup> |
| **Botswana** | **20.07%** | **NI** |
| South Africa | 19.96% | 12<sup>th</sup> |
| Mozambique | 13.57% | 8<sup>th</sup> |
| Zimbabwe | 12.01% | 7<sup>th</sup> |
| Namibia | 11.56% | 2<sup>nd</sup> |
| Zambia | 10.31% | 5<sup>th</sup> |
| Malawi | 8.42% | 9<sup>th</sup> |
| Equatorial Guinea | 7.93% | - |

**Note.** HIV prevalence measures the share of people aged 15 to 49 years old who are infected with HIV. Botswana is highlighted as it was not in the study sample. Among top 10 highest HIV prevalence counties, Equatorial Guinea was the only one not among the 15 highest ODApc recipients between 2002-2019. See Figure below for the highest annual SRH ODA recipients in the sample LLMICs.
**Data Sources.** Institute for Health Metrics and Evaluation, OECD-CRS.

**Figure D.1:**
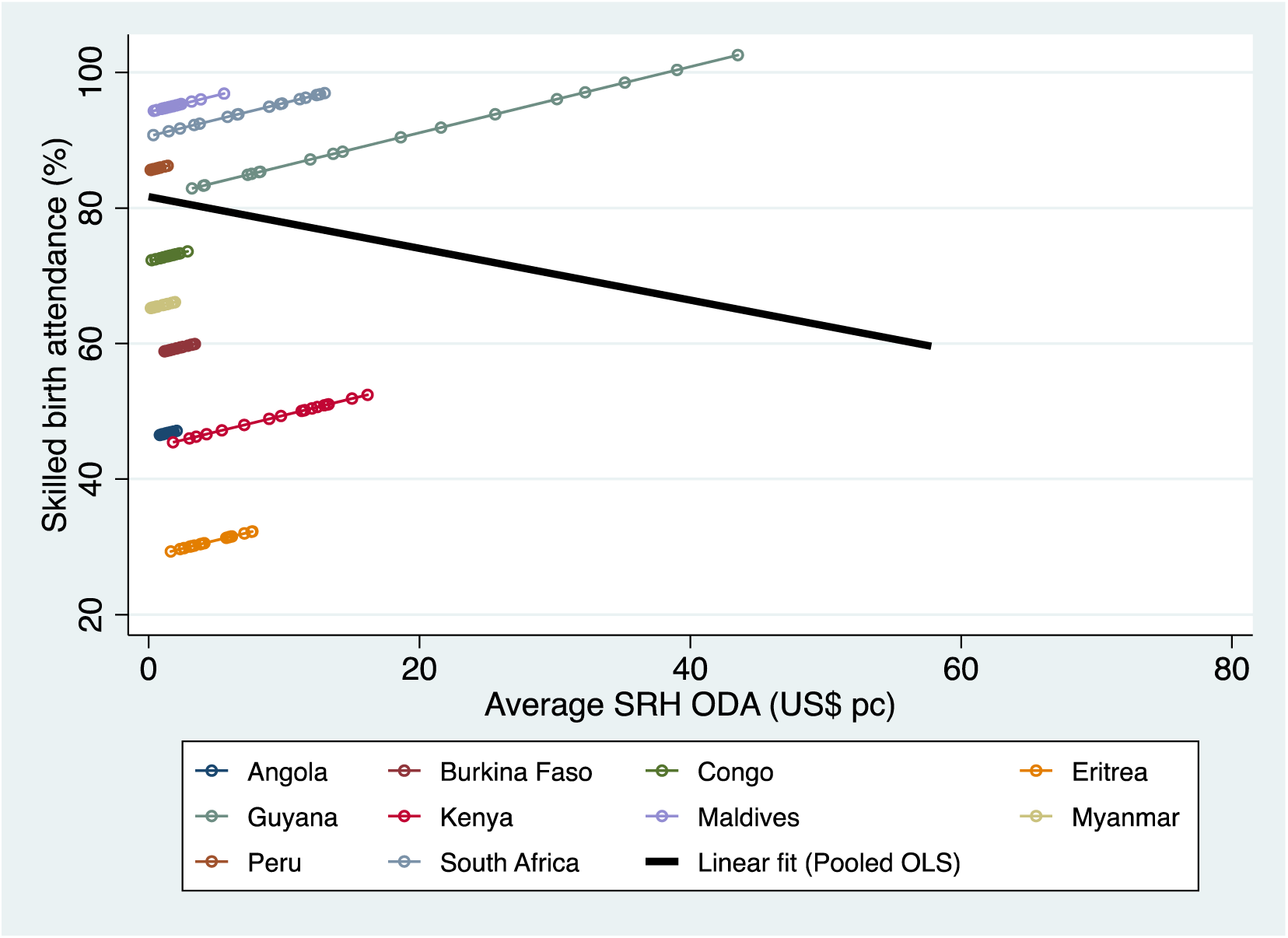
Difference between bivariate linear prediction (pooled OLS) and bivariate country-fixed effect estimation of SBA by SRH ODA. *Note.* Figure C1 illustrates the discrepancy between fitting a FE OLS and a pooled OLS to the panel data. The estimates are based on the entire sample of countries. A random selection of countries is presented in the graph to avoid overcrowding. The FE estimation allows for heterogeneities between individual countries, reflected in country-specific intercepts. Using FE an average positive effect of SRH ODA across countries is estimated, represented by the common upwards slope. In contrast, the linear fit indicates that the effect estimate of SRH ODA from the bivariate OLS is negative (*b*=-.382, [-.677, -.086]).^3^ *Data sources.* OECD-CRS, WDI. ^3^ The divergence in the slopes from the country-FE OLS compared to the pooled OLS suggests that country level factors play a large role in the effect of SRH ODA on SBA. This was confirmed by appropriate tests.

**Figure D.2:**
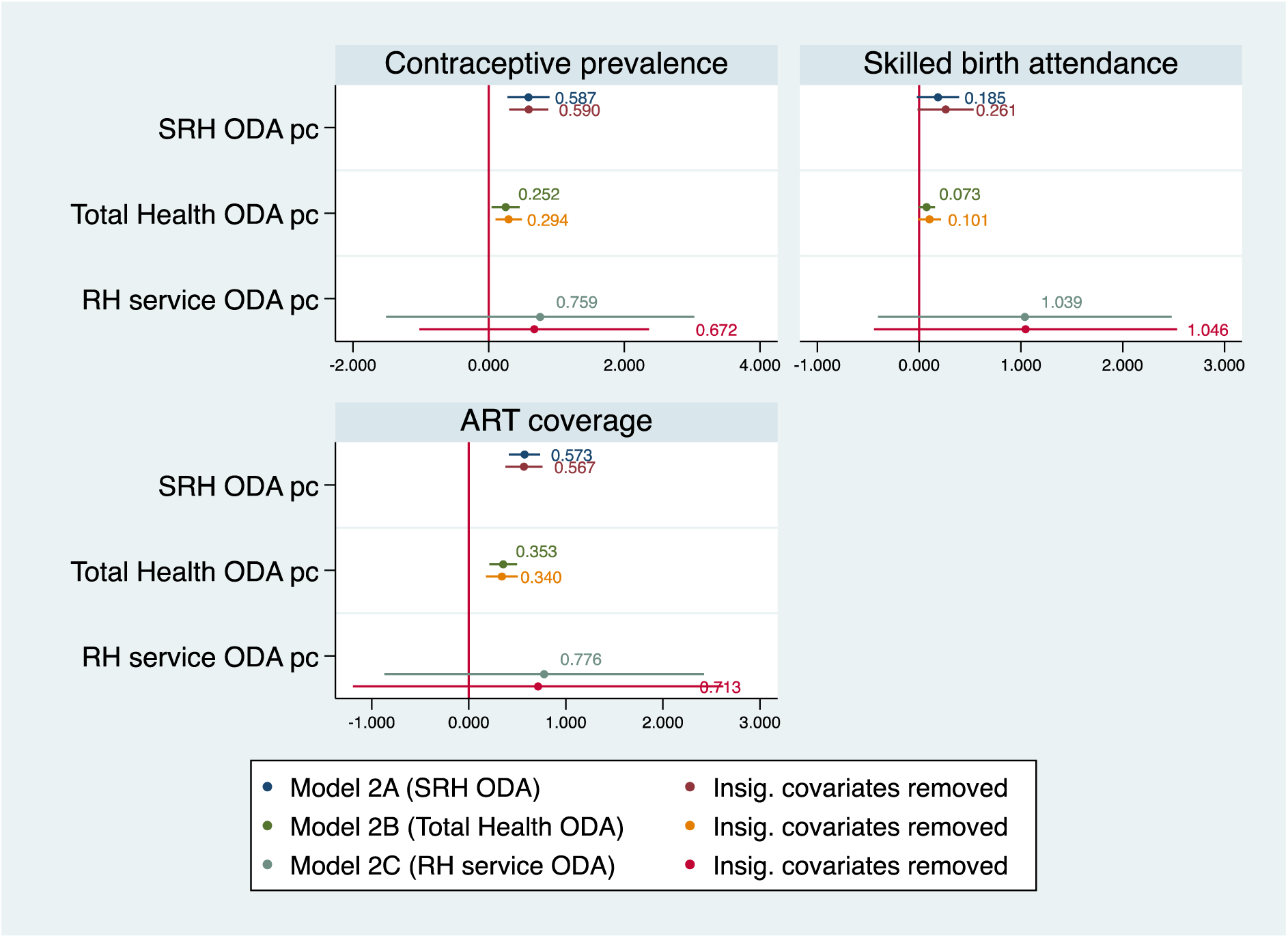
Main effect estimates compared to effect estimates from models including only significant controls. ***Note.*** Figure C2 displays the main effect estimates for health ODA on service coverage next to effect estimates from models that only included controls significant at p<0.1. All models included country and year FE. The models for mCP controlled for adolescent fertiltiy and government effectivenss. Models for SBA inclueded GDP pc (ln), population densitiy (ln) and adolescent fertility. Models on ART coverage inclueded only population desnitiy (ln). The rope-ladder plot show markers for point estimates, and spikes for confidence intervals at 95% levels. Those spikes crossing the reference line at zero show coefficients that are not significantly different from zero. ***Data sources.*** OECD-CRS, WDI, WGI.

**Figure D.3:**
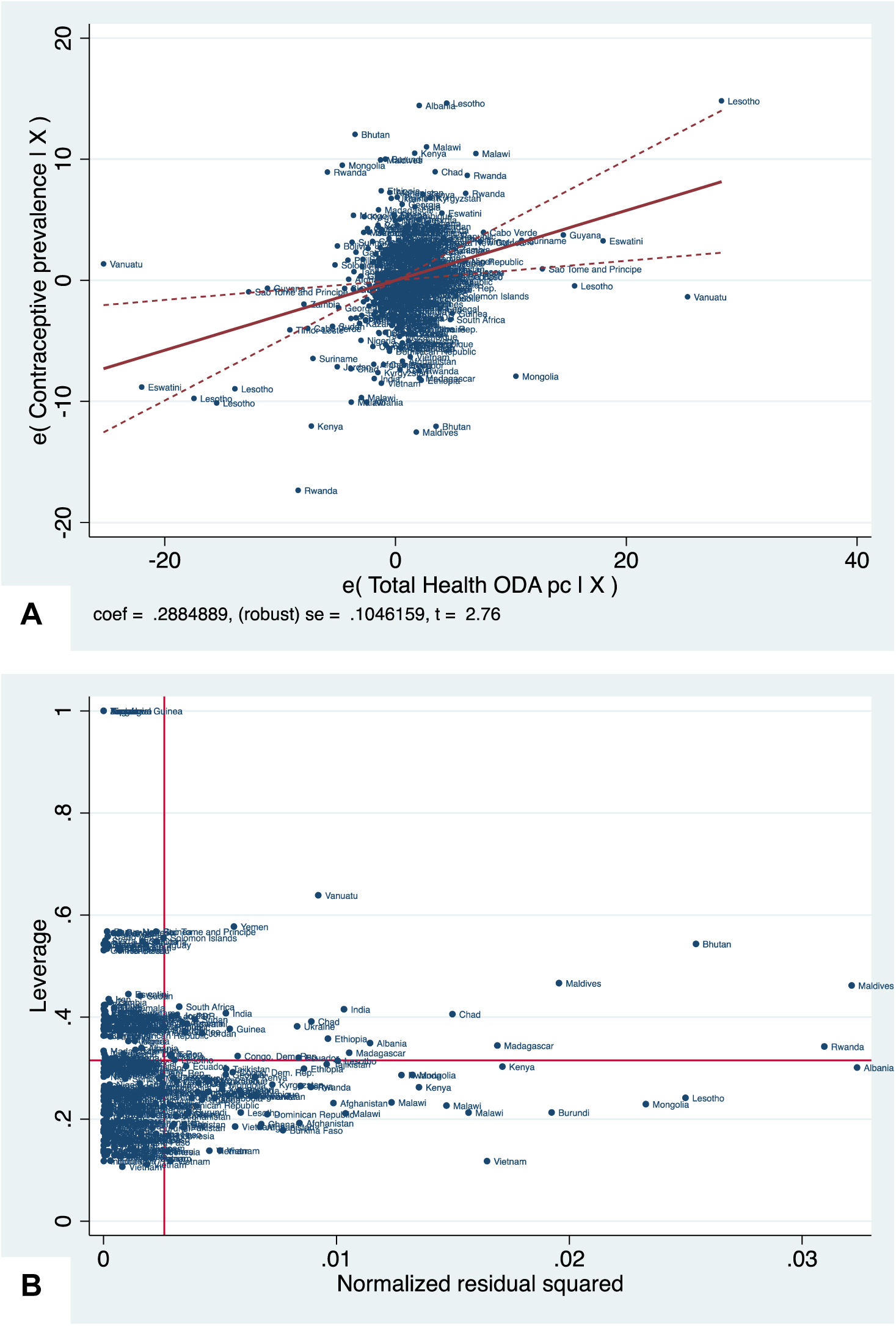
Outlier identification graphs for estimation of mCP by TotH ODApc. ***Note.*** Panel A shows the added variable plot for TotH ODApc from the FE estimation of mCP. Panel B shows the leverage vs. standardized residuals plot. Observation with high leverage and high normalised residuals are potentially influential. Removing 27 observation with cook’s distance>4/N-k-1 from different countries, decreasing the sample from 90 to 84 countries.^4^ TotH ODA effect estimate remained positive significant, GHE/CHE and government effectiveness became significant predictors of mCP (at 95% CI). ***Data sources.*** OECD-CRS, WDI, WGI ^4^ Outliers included observations from LICs (Bhutan, Chad, Ethiopia, India, Kenya, Lesotho, Madagascar, Mongolia, Rwanda, Salomon Islands, Yemen) and LMICs (Albania, Maldives, Ukraine, Vanuatu).

**Figure D.4:**
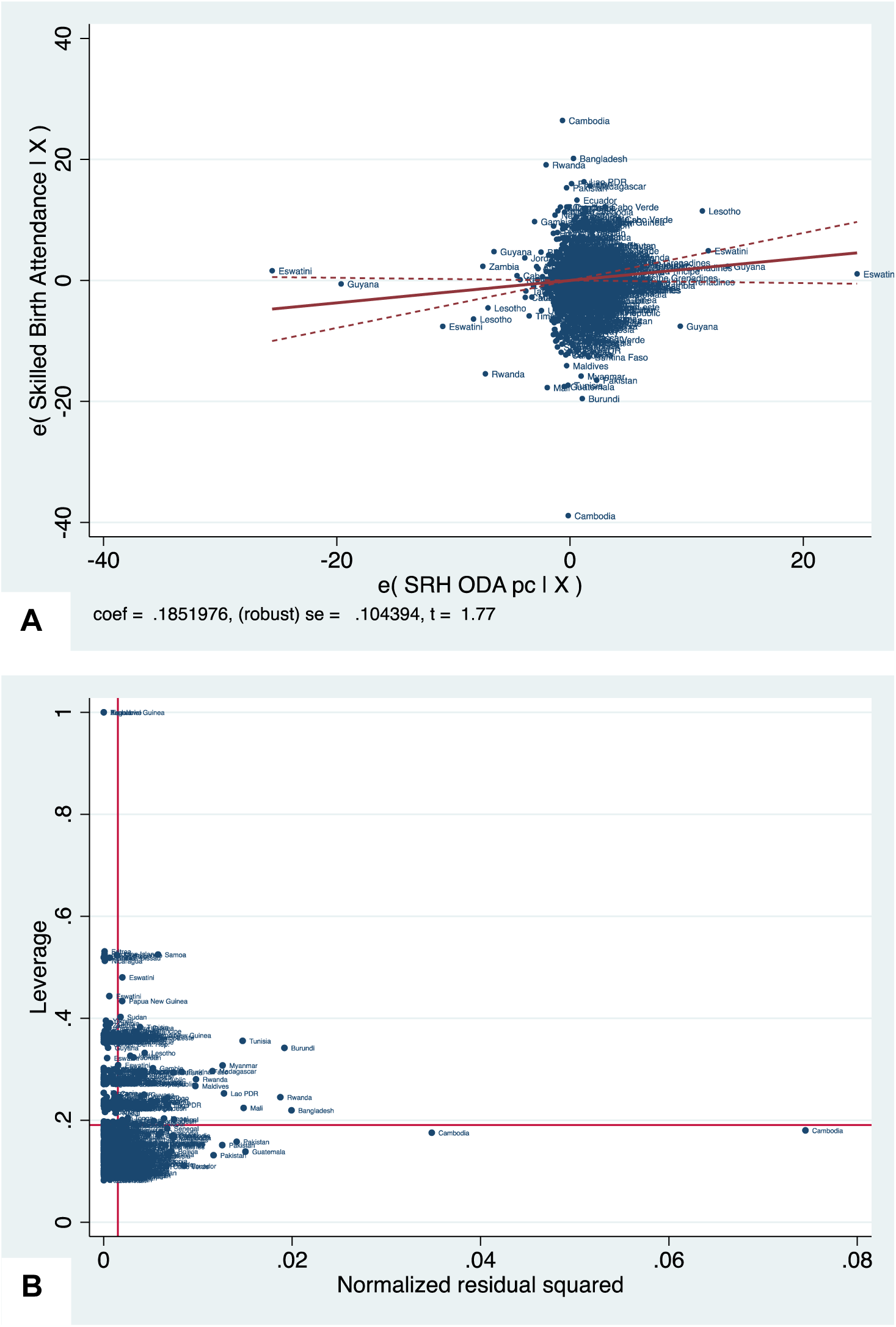
Outlier identification graphs for estimation of SBA by SRH ODApc. ***Note.*** Panel A shows the added variable plot for SRH ODApc from the FE estimation of SBA. Panel B shows the leverage vs. standardized residuals plot, observation with high leverage and high normalised residuals are potentially influential. Removing 72 observations with cook’s distance>4/N-k-1 from different countries, decreased the analysis sample from 97 to 89 countries. The effect of SRH ODA effect became positive significant (*b*=.215, 95% CI [.039, .391]. ***Data sources.*** OECD-CRS, WDI, WGI

**Figure D.5:**
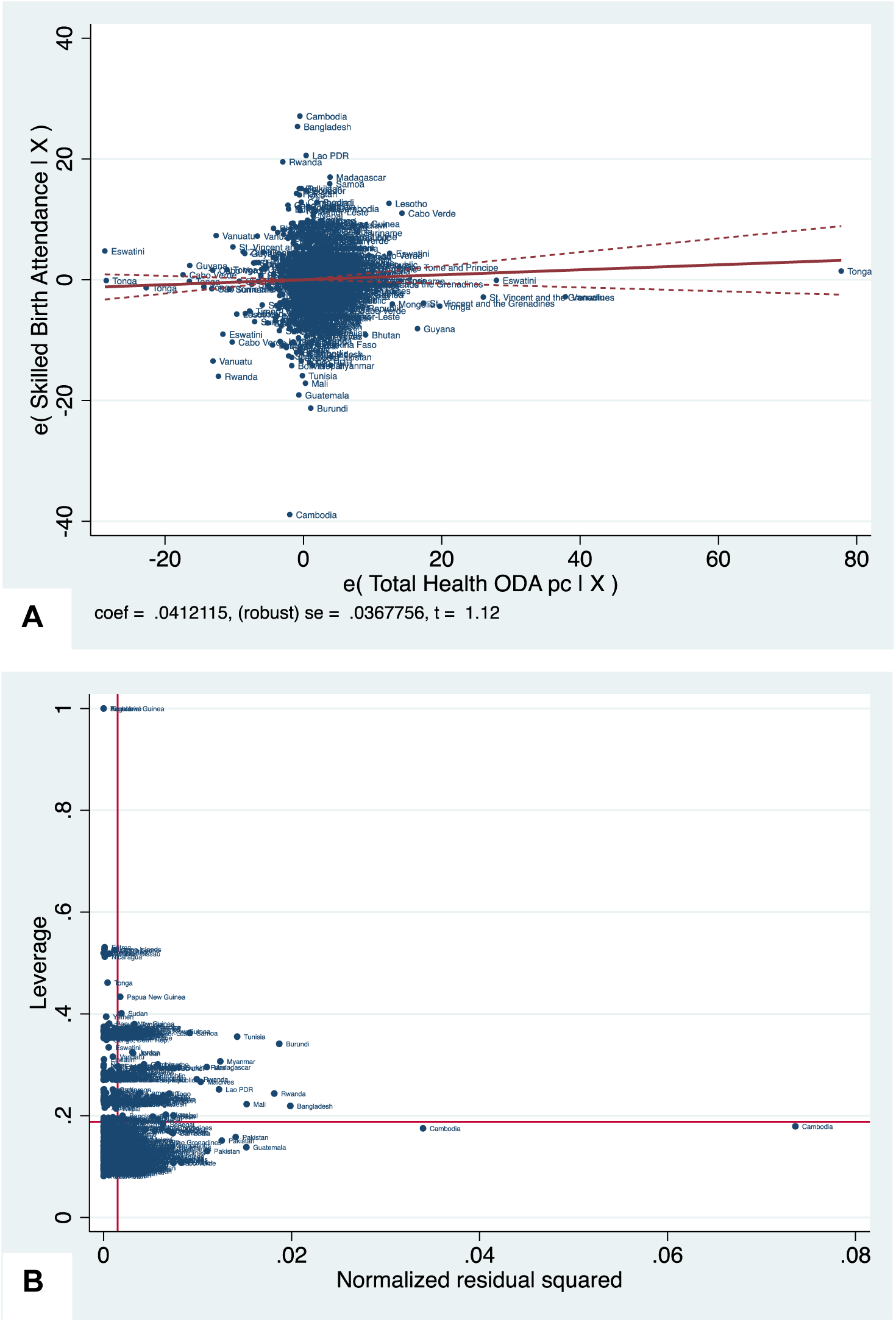
Outlier identification graphs for estimation of SBA by TotH ODApc. ***Note.*** Panel A shows the added variable plot of TotH ODApc from the main model. Panel B shows the leverage vs. standardized residuals plot. Observation with high leverage and high normalised residuals are potentially influential. Removing 70 observations with cook’s distance>4/N-k-1 from different countries, reduced the sample from 97 to 89 countries. The effect of TotH ODA in LMICs was no longer observed. ***Data sources.*** OECD-CRS, WDI, WGI

**Figure D.6:**
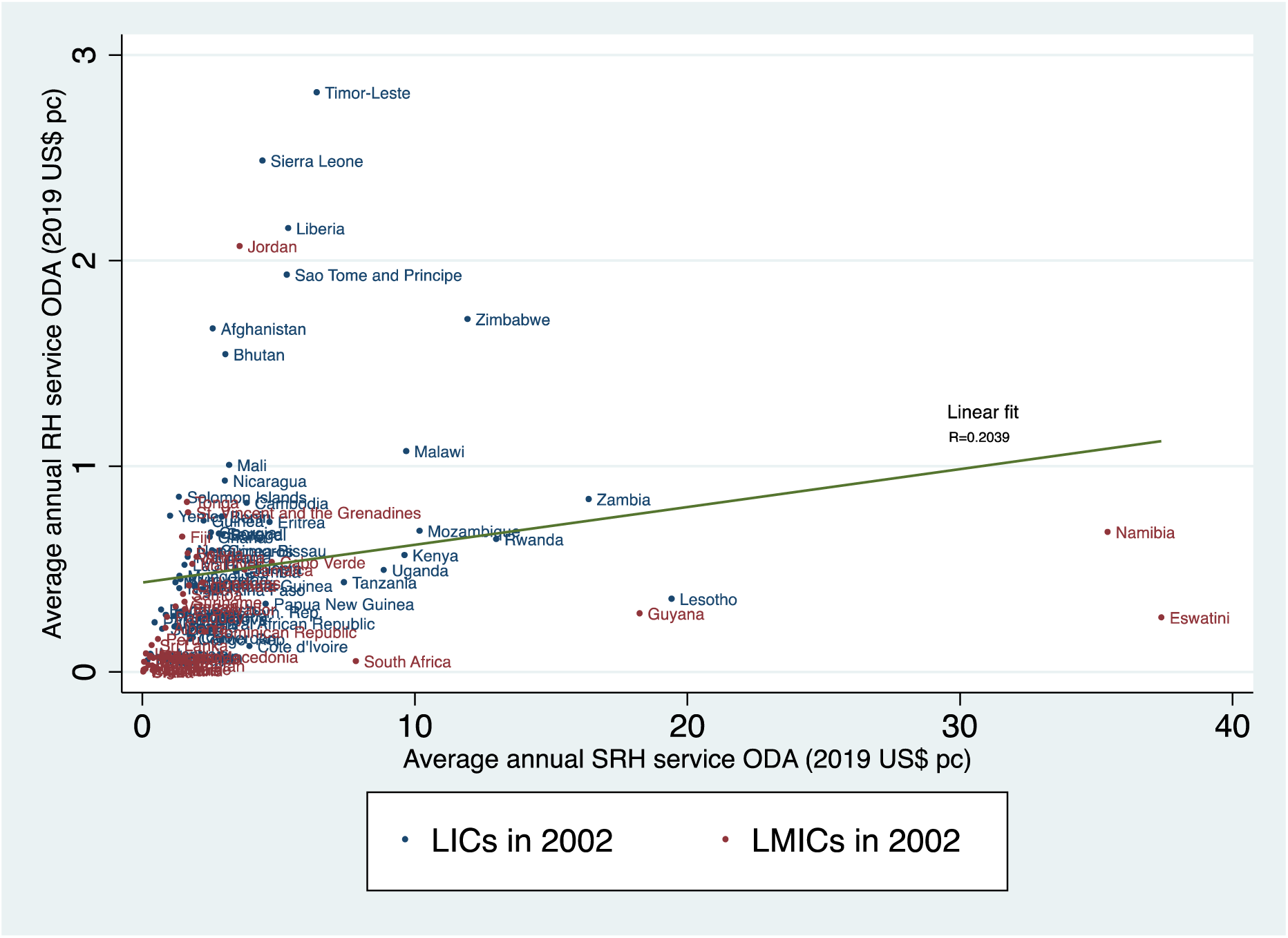
Scatterplot of average annual SRH ODA against average annual RH care ODA (2019 US$ pc) received by sample countries over 2002-2019. ***Note.*** Figure C6 looks at whether the same countries that received significant amounts of annual RH care ODApc were also high SRH ODApc recipients. *Data sources.* OECD-CRS.

**Figure D.7:**
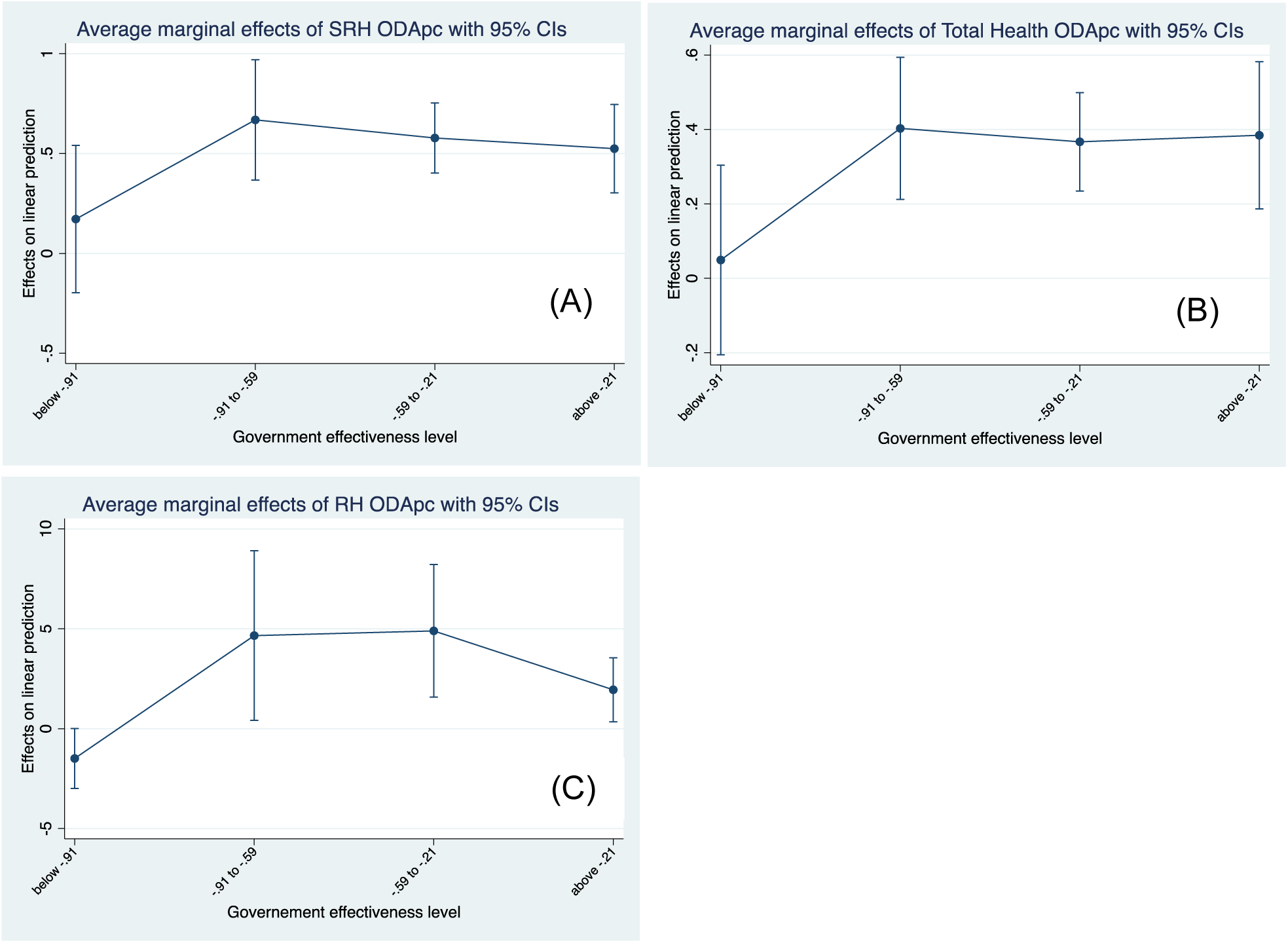
Marginal effect of health ODA (2019 US$, pc) on ART coverage by level of government effectiveness. ***Note.*** Categories of government effectiveness reflect the average rating of countries over the period 2002-2019. The levels were defined using quartile values in the sample of countries used in the ART analysis. All measures of health ODA had a significant impact on ART coverage in countries with an average government effectiveness rating above -0.91 (25 percentile value in the sample countries). *Data sources.* OECD-CRS, WDI, WGI.

**Figure D.8:**
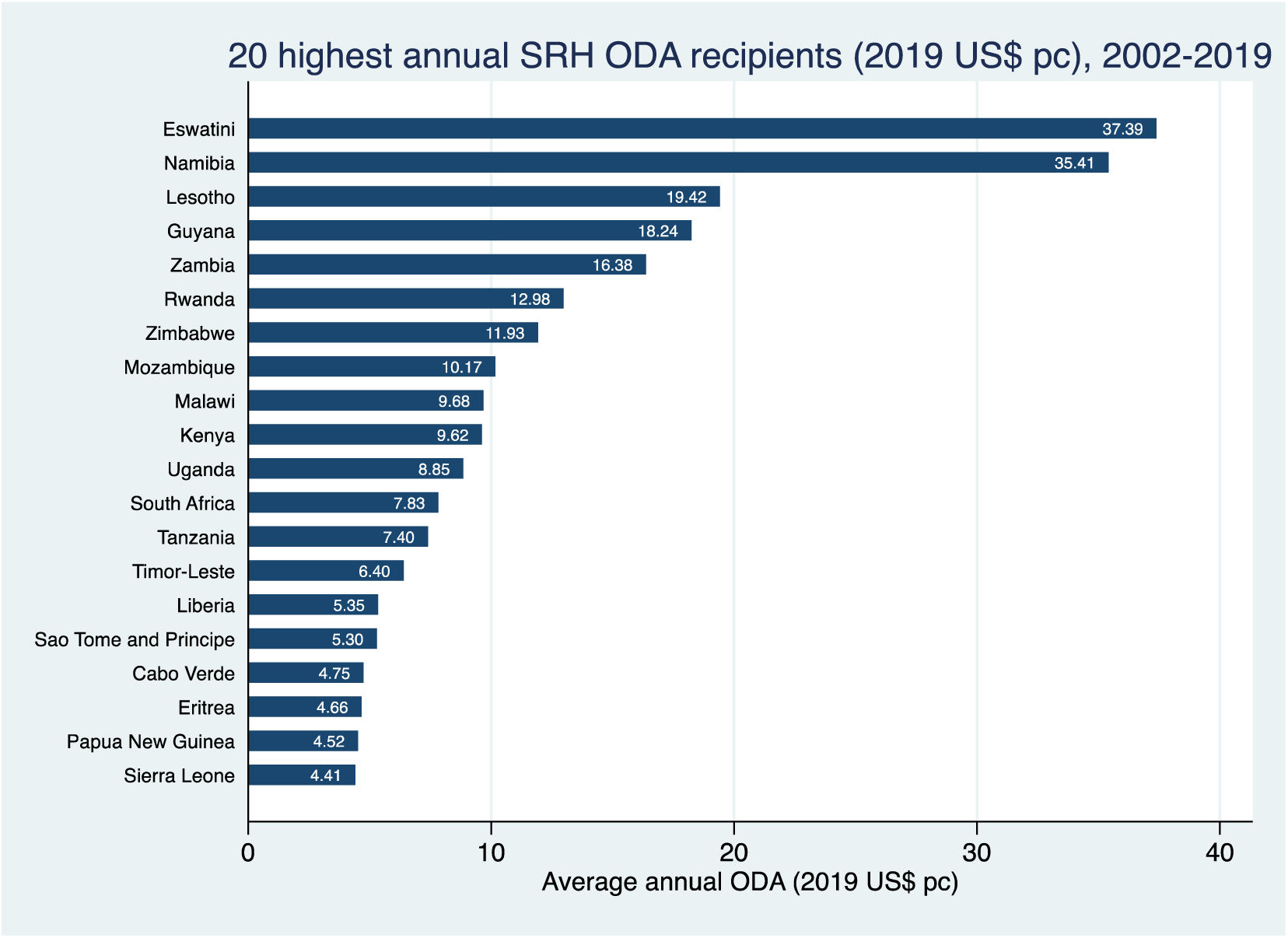
20 highest SRH ODA recipients, 2002-2019.

## Appendix E. Sensitivity analysis

Our sensitivity analyses aim to account for shortcomings of the data and the main FE estimation. For each of these approaches, we first provide a summary table showing the ODA effect estimates from the main model next to the effect estimates from the sensitivity analyses. Each summary table is then followed by complete tables, including parameter estimates and robust standard errors.

### E.1: Sensitivity to missing data and measurement error

**Table E.1:** The effects of health ODA on SRH services’ summary of sensitivity analysis for missing data and measurement error.

| Sectoral ODA | SRH service outcomes |  |  |
| --- | --- | --- | --- |
|  | Contraceptive Prevalence | Skilled birth attendance | ART coverage |
| **SRH ODA** |  |  |  |
| Main FE Estimator | 0.628*** | 0.190* | 0.531*** |
| FE Interpolated Variables | 0.482*** | 0.247*** | 0.545*** |
| FE 3-Year-Averaged Variables | 0.531*** | 0.314** | 0.593*** |
| **Total Health ODA** |  |  |  |
| Main FE Estimator | 0.242** | 0.059 | 0.332*** |
| FE Interpolated Variables | 0.196*** | 0.080* | 0.331*** |
| FE 3-Year-Averaged Variables | 0.258** | 0.115 | 0.370*** |
| **RH care ODA** |  |  |  |
| Main FE Estimator | 0.935 | 0.902 | 0.575 |
| FE Interpolated Variables | 1.113** | 1.413*** | 0.649 |
| FE 3-Year-Averaged Variables | 1.481 | 0.649 | 1.181 |

*Note.* All estimators include country and time fixed effects. All measures of health ODA are in USD per capita (constant 2020). Standard errors are clustered at the country level. \*\*\* $p < 0.01$ , \*\* $p < 0.05$ , \* $p < 0.1$ . In the Interpolated Variables model, all covariates and the dependent variable (SRH service indicators) are interpolated with STATA command *ipolate*. In the 3-Year-Averaged Variables model, all variables are averaged over three years resulting in six 3-year periods.

**Table E.2:** Sensitivity for missing data - Effects of sectoral ODA per capita on modern contraceptive prevalence.

| VARIABLES | SRH ODA |  |  | Total Health ODA |  |  | RH care ODA |  |  |
| --- | --- | --- | --- | --- | --- | --- | --- | --- | --- |
|  | Main | Interpolate | 3-year-avg | Main | Interpolate | 3-year-avg | Main | Interpolate | 3-year-avg |
| Health ODA pc | 0.628***<br>(0.150) | 0.482***<br>(0.129) | 0.531***<br>(0.163) | 0.242**<br>(0.106) | 0.196***<br>(0.065) | 0.258**<br>(0.120) | 0.935<br>(1.221) | 1.113**<br>(0.455) | 1.480<br>(1.487) |
| ln GDP pc | 1.735<br>(4.800) | 4.156<br>(4.849) | 5.914<br>(5.060) | 1.820<br>(5.452) | 4.414<br>(5.102) | 5.939<br>(5.725) | -0.699<br>(5.490) | 3.250<br>(5.016) | 3.999<br>(5.561) |
| GHE share of CHE | 0.027<br>(0.065) | 0.023<br>(0.052) | 0.028<br>(0.073) | 0.061<br>(0.073) | 0.031<br>(0.054) | 0.044<br>(0.080) | 0.039<br>(0.069) | 0.003<br>(0.053) | -0.010<br>(0.079) |
| Government effective | 3.995*<br>(2.249) | 1.563<br>(2.169) | 8.048***<br>(3.063) | 3.345<br>(2.499) | 1.367<br>(2.386) | 7.964**<br>(3.297) | 3.717<br>(2.997) | 1.158<br>(2.748) | 8.665**<br>(3.551) |
| Corruption control | 1.504<br>(2.669) | 2.568<br>(2.263) | 0.070<br>(3.179) | 2.372<br>(2.979) | 2.712<br>(2.396) | -0.227<br>(3.353) | 3.275<br>(3.294) | 3.177<br>(2.583) | 0.120<br>(3.557) |
| ln Population density | 23.558**<br>(9.288) | 22.457**<br>(10.869) | 32.531***<br>(10.512) | 21.369**<br>(10.689) | 19.836*<br>(11.828) | 30.499***<br>(10.918) | 21.158*<br>(12.400) | 18.839<br>(12.611) | 29.369**<br>(11.839) |
| Primary schooling | 0.064<br>(0.041) | 0.037<br>(0.046) | 0.103**<br>(0.044) | 0.050<br>(0.043) | 0.034<br>(0.045) | 0.094**<br>(0.045) | 0.048<br>(0.045) | 0.032<br>(0.044) | 0.089**<br>(0.043) |
| Female labour part. | -0.090<br>(0.133) | -0.072<br>(0.125) | 0.062<br>(0.153) | -0.149<br>(0.155) | -0.129<br>(0.145) | 0.037<br>(0.157) | -0.213<br>(0.201) | -0.175<br>(0.189) | -0.041<br>(0.195) |
| Observations | 387 | 1,152 | 328 | 390 | 1,157 | 328 | 383 | 1,132 | 327 |
| N. of countries | 95 | 101 | 102 | 96 | 101 | 102 | 94 | 101 | 102 |
| Two-way FE | YES | YES | YES | YES | YES | YES | YES | YES | YES |
| Adj. within R-squared | 0.400 | 0.380 | 0.416 | 0.351 | 0.348 | 0.392 | 0.323 | 0.327 | 0.367 |
| Rho | 0.980 | 0.987 | 0.987 | 0.976 | 0.984 | 0.985 | 0.977 | 0.983 | 0.984 |

*Note.* All measures of health ODA are in USD per capita (constant 2020). Standard errors are clustered at the country level. \*\*\* p<0.01, \*\* p<0.05, \* p<0.1, and SE in parenthesis.

**Table E.3:** Sensitivity for missing data - Effects of sectoral ODA per capita on skilled birth attendance.

| VARIABLES | SRH ODA |  |  | Total Health ODA |  |  | RH care ODA |  |  |
| --- | --- | --- | --- | --- | --- | --- | --- | --- | --- |
|  | Main | Interpolate | 3-year-avg | Main | Interpolate | 3-year-avg | Main | Interpolate | 3-year-avg |
| Health ODA pc | 0.190*<br>(0.100) | 0.247***<br>(0.075) | 0.314**<br>(0.120) | 0.059<br>(0.044) | 0.080*<br>(0.043) | 0.115<br>(0.083) | 0.902<br>(1.015) | 1.413***<br>(0.523) | 2.032*<br>(1.130) |
| ln GDP pc | 8.615<br>(6.305) | 16.344***<br>(6.153) | 20.513***<br>(7.694) | 8.701<br>(6.290) | 16.435***<br>(6.214) | 20.689***<br>(7.833) | 6.759<br>(6.319) | 14.031**<br>(6.037) | 18.546**<br>(7.654) |
| GHE share of CHE | 0.108<br>(0.066) | 0.110*<br>(0.056) | 0.199**<br>(0.084) | 0.108*<br>(0.065) | 0.105*<br>(0.056) | 0.199**<br>(0.084) | 0.125*<br>(0.065) | 0.110*<br>(0.056) | 0.191**<br>(0.083) |
| Government effective | -4.585<br>(2.887) | -3.432<br>(2.556) | -3.482<br>(4.554) | -3.935<br>(2.698) | -2.985<br>(2.469) | -2.860<br>(4.391) | -4.575<br>(2.911) | -3.187<br>(2.655) | -3.507<br>(4.590) |
| Corruption control | 5.053*<br>(2.943) | 3.459<br>(2.874) | 4.118<br>(4.303) | 4.772*<br>(2.704) | 3.274<br>(2.758) | 3.696<br>(4.327) | 5.812*<br>(3.080) | 4.213<br>(3.049) | 4.917<br>(4.480) |
| ln Population density | 47.320***<br>(11.772) | 47.544***<br>(12.927) | 54.167***<br>(13.699) | 48.155***<br>(11.492) | 47.295***<br>(12.718) | 54.373***<br>(13.284) | 44.140***<br>(11.962) | 42.268***<br>(12.830) | 48.259***<br>(13.677) |
| Primary schooling | 0.077<br>(0.091) | 0.035<br>(0.083) | 0.131<br>(0.103) | 0.076<br>(0.090) | 0.036<br>(0.081) | 0.127<br>(0.102) | 0.081<br>(0.091) | 0.036<br>(0.081) | 0.119<br>(0.103) |
| Female labour part. | 0.105<br>(0.267) | 0.127<br>(0.234) | 0.309<br>(0.267) | 0.106<br>(0.267) | 0.110<br>(0.233) | 0.303<br>(0.264) | 0.060<br>(0.259) | 0.066<br>(0.242) | 0.236<br>(0.273) |
| Observations | 685 | 1,347 | 416 | 695 | 1,360 | 417 | 663 | 1,308 | 412 |
| N. of countries | 100 | 103 | 104 | 101 | 103 | 104 | 100 | 103 | 104 |
| Two-way FE | YES | YES | YES | YES | YES | YES | YES | YES | YES |
| Adj. within R-squared | 0.437 | 0.496 | 0.508 | 0.432 | 0.490 | 0.505 | 0.448 | 0.506 | 0.511 |
| Rho | 0.987 | 0.989 | 0.987 | 0.988 | 0.989 | 0.987 | 0.986 | 0.986 | 0.983 |

*Note.* All measures of health ODA are in USD per capita (constant 2020). Standard errors are clustered at the country level. \*\*\* p<0.01, \*\* p<0.05, \* p<0.1, and SE in parenthesis.

**Table E.4:** Sensitivity for missing data - Effects of sectoral ODA per capita on ART coverage.

| VARIABLES | SRH ODA |  |  | Total Health ODA |  |  | RH care ODA |  |  |
| --- | --- | --- | --- | --- | --- | --- | --- | --- | --- |
|  | Main | Interpolate | 3-year-avg | Main | Interpolate | 3-year-avg | Main | Interpolate | 3-year-avg |
| Health ODA pc | 0.531***<br>(0.076) | 0.545***<br>(0.079) | 0.593***<br>(0.124) | 0.332***<br>(0.070) | 0.331***<br>(0.070) | 0.370***<br>(0.104) | 0.575<br>(0.833) | 0.649<br>(0.767) | 1.181<br>(1.254) |
| ln GDP pc | -1.695<br>(7.291) | 1.798<br>(7.087) | 3.339<br>(7.839) | -2.335<br>(7.366) | 1.464<br>(7.219) | 2.914<br>(8.023) | -3.497<br>(7.384) | 0.347<br>(7.196) | 1.830<br>(7.982) |
| GHE share of CHE | -0.014<br>(0.070) | -0.029<br>(0.068) | 0.002<br>(0.091) | -0.014<br>(0.071) | -0.028<br>(0.068) | 0.006<br>(0.093) | -0.047<br>(0.070) | -0.061<br>(0.068) | -0.037<br>(0.090) |
| Government effective | 6.128*<br>(3.255) | 5.172<br>(3.415) | 6.433<br>(4.760) | 6.088*<br>(3.238) | 5.201<br>(3.415) | 6.775<br>(4.862) | 6.274*<br>(3.482) | 5.391<br>(3.606) | 6.664<br>(4.930) |
| Corruption control | 5.274*<br>(3.108) | 4.836<br>(3.077) | 4.717<br>(3.831) | 5.278*<br>(3.165) | 4.635<br>(3.155) | 4.204<br>(3.965) | 6.412*<br>(3.298) | 5.951*<br>(3.252) | 5.280<br>(4.005) |
| ln Population density | 14.208<br>(15.400) | 15.592<br>(14.924) | 20.802<br>(14.748) | 9.790<br>(15.435) | 11.600<br>(15.038) | 17.279<br>(14.883) | 9.995<br>(16.938) | 11.923<br>(16.358) | 16.126<br>(15.732) |
| Primary schooling | -0.052<br>(0.071) | -0.038<br>(0.069) | -0.036<br>(0.075) | -0.056<br>(0.070) | -0.041<br>(0.068) | -0.045<br>(0.074) | -0.053<br>(0.072) | -0.037<br>(0.070) | -0.046<br>(0.075) |
| Female labour part. | -0.181<br>(0.295) | -0.121<br>(0.284) | -0.250<br>(0.326) | -0.184<br>(0.297) | -0.116<br>(0.289) | -0.247<br>(0.331) | -0.246<br>(0.310) | -0.192<br>(0.297) | -0.340<br>(0.331) |
| Observations | 1,306 | 1,434 | 486 | 1,307 | 1,436 | 486 | 1,290 | 1,415 | 486 |
| Number of countries | 90 | 90 | 90 | 90 | 90 | 90 | 90 | 90 | 90 |
| Two-way FE | YES | YES | YES | YES | YES | YES | YES | YES | YES |
| Adj. within R-squared | 0.857 | 0.856 | 0.867 | 0.854 | 0.853 | 0.864 | 0.845 | 0.844 | 0.858 |
| Rho | 0.897 | 0.906 | 0.934 | 0.850 | 0.866 | 0.914 | 0.861 | 0.878 | 0.913 |

*Note.* All measures of health ODA are in USD per capita (constant 2020). Standard errors are clustered at the country level. \*\*\* p<0.01, \*\* p<0.05, \* p<0.1, and SE in parenthesis.

### E.2: Influence of outlying observations

**Table E.5:**
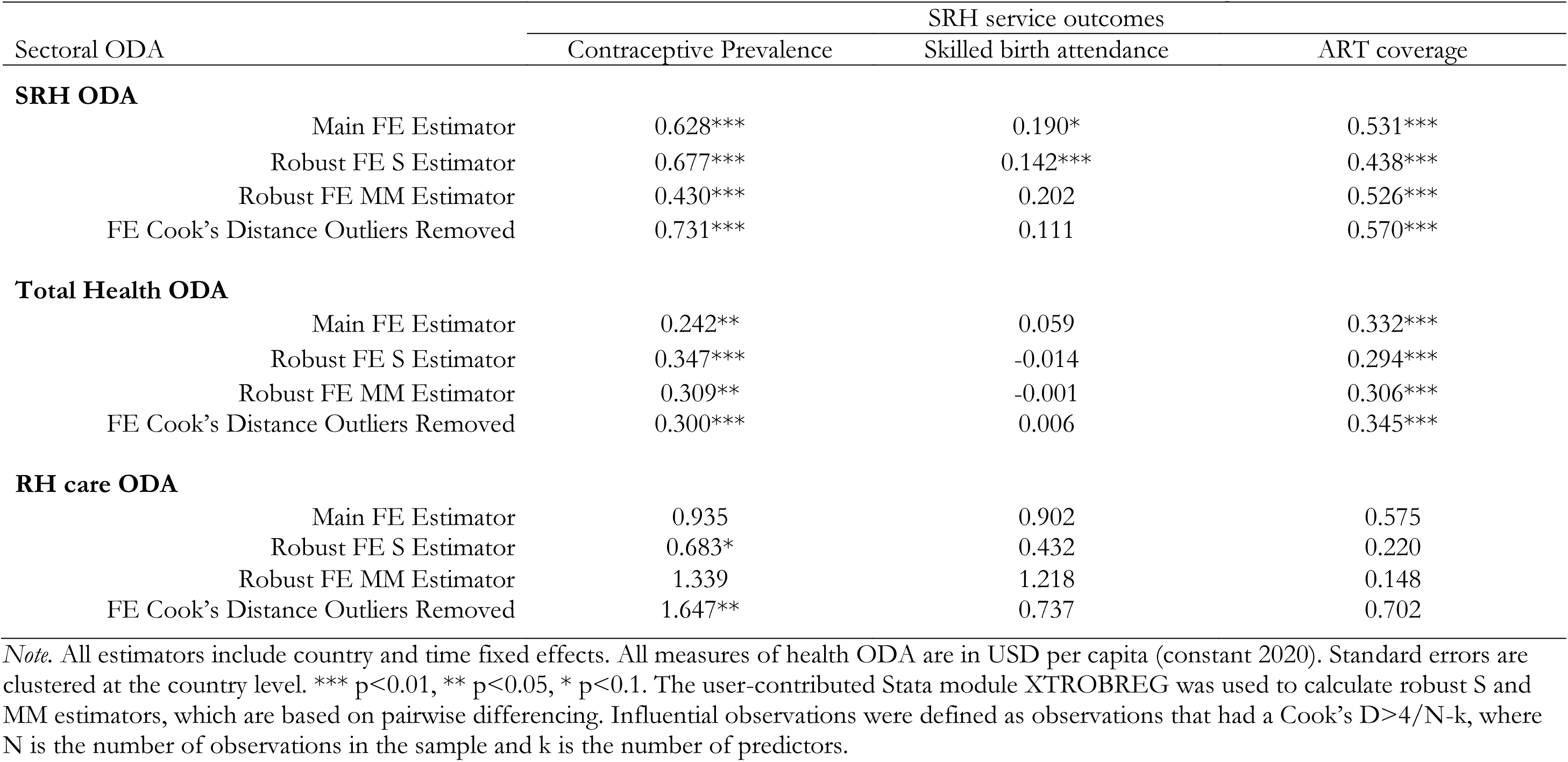
The effects of health ODA on SRH services’ summary of sensitivity analysis for outlying observations.

| Sectoral ODA | SRH service outcomes |  |  |
| --- | --- | --- | --- |
|  | Contraceptive Prevalence | Skilled birth attendance | ART coverage |
| **SRH ODA** |  |  |  |
| Main FE Estimator | 0.628*** | 0.190* | 0.531*** |
| Robust FE S Estimator | 0.677*** | 0.142*** | 0.438*** |
| Robust FE MM Estimator | 0.430*** | 0.202 | 0.526*** |
| FE Cook's Distance Outliers Removed | 0.731*** | 0.111 | 0.570*** |
| **Total Health ODA** |  |  |  |
| Main FE Estimator | 0.242** | 0.059 | 0.332*** |
| Robust FE S Estimator | 0.347*** | -0.014 | 0.294*** |
| Robust FE MM Estimator | 0.309** | -0.001 | 0.306*** |
| FE Cook's Distance Outliers Removed | 0.300*** | 0.006 | 0.345*** |
| **RH care ODA** |  |  |  |
| Main FE Estimator | 0.935 | 0.902 | 0.575 |
| Robust FE S Estimator | 0.683* | 0.432 | 0.220 |
| Robust FE MM Estimator | 1.339 | 1.218 | 0.148 |
| FE Cook's Distance Outliers Removed | 1.647** | 0.737 | 0.702 |

*Note.* All estimators include country and time fixed effects. All measures of health ODA are in USD per capita (constant 2020). Standard errors are clustered at the country level. \*\*\* $p < 0.01$ , \*\* $p < 0.05$ , \* $p < 0.1$ . The user-contributed Stata module XTROBREG was used to calculate robust S and MM estimators, which are based on pairwise differencing. Influential observations were defined as observations that had a Cook's $D > 4/(N-k)$ , where $N$ is the number of observations in the sample and $k$ is the number of predictors.

**Table E.6:**
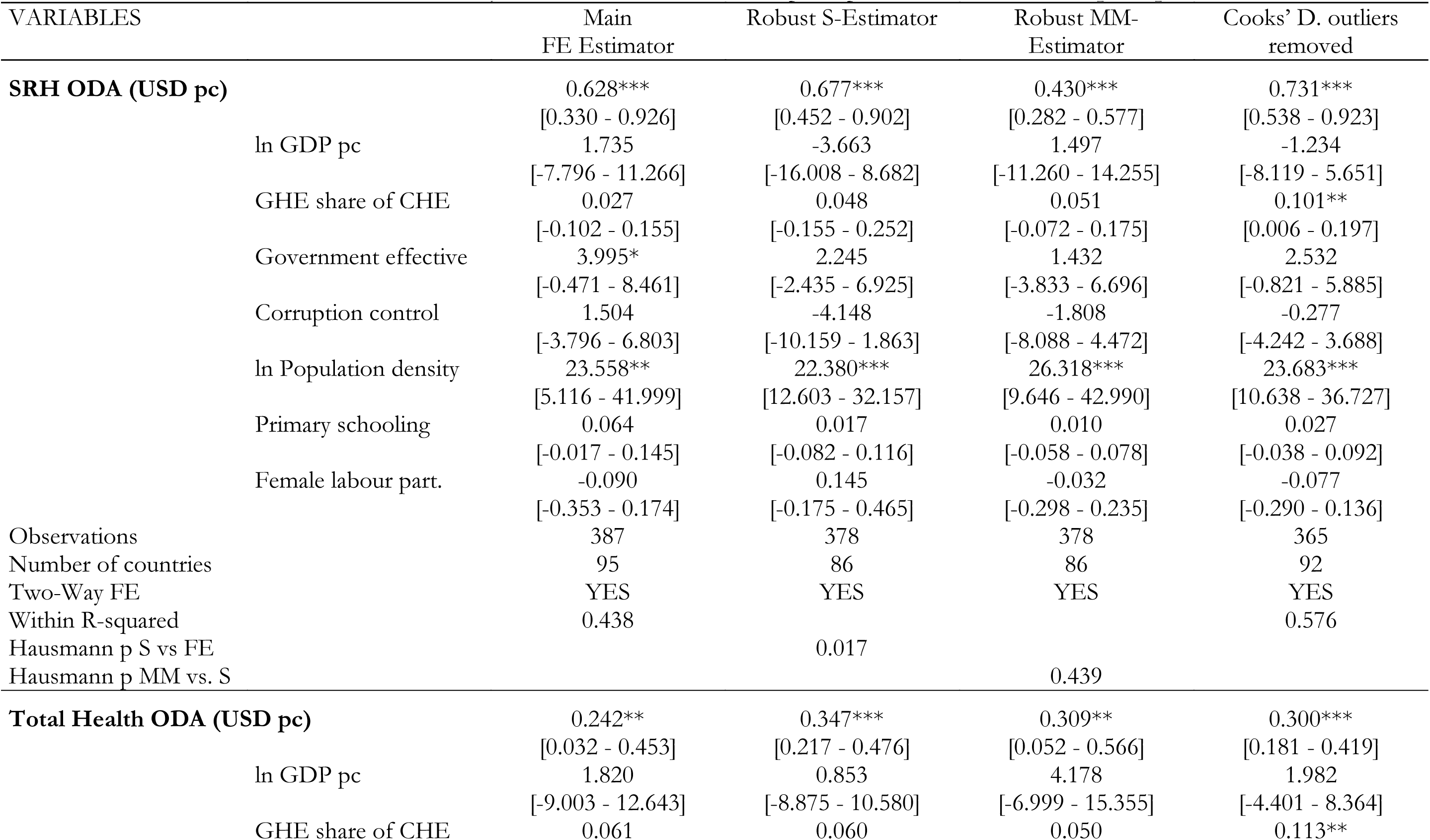
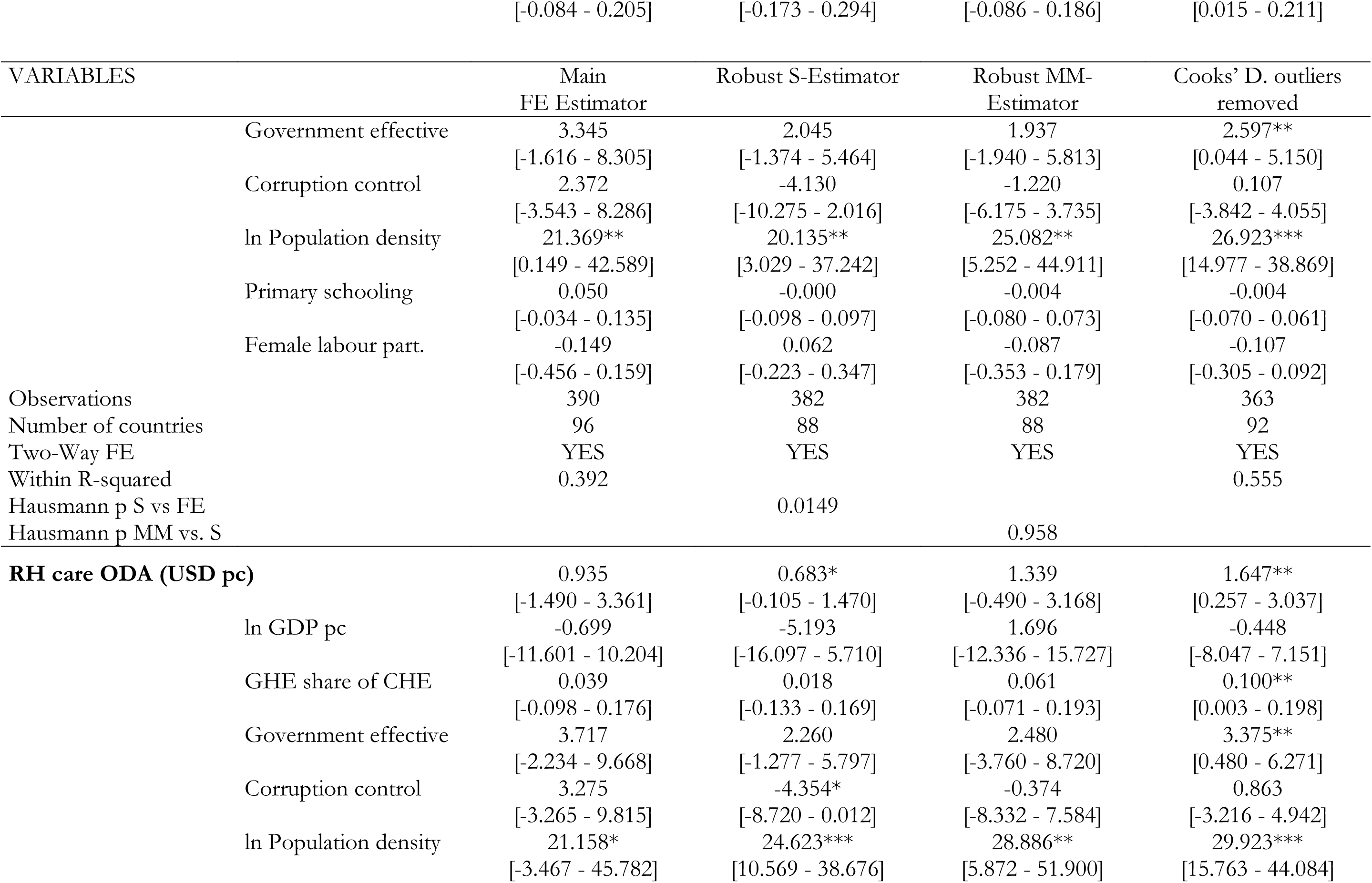
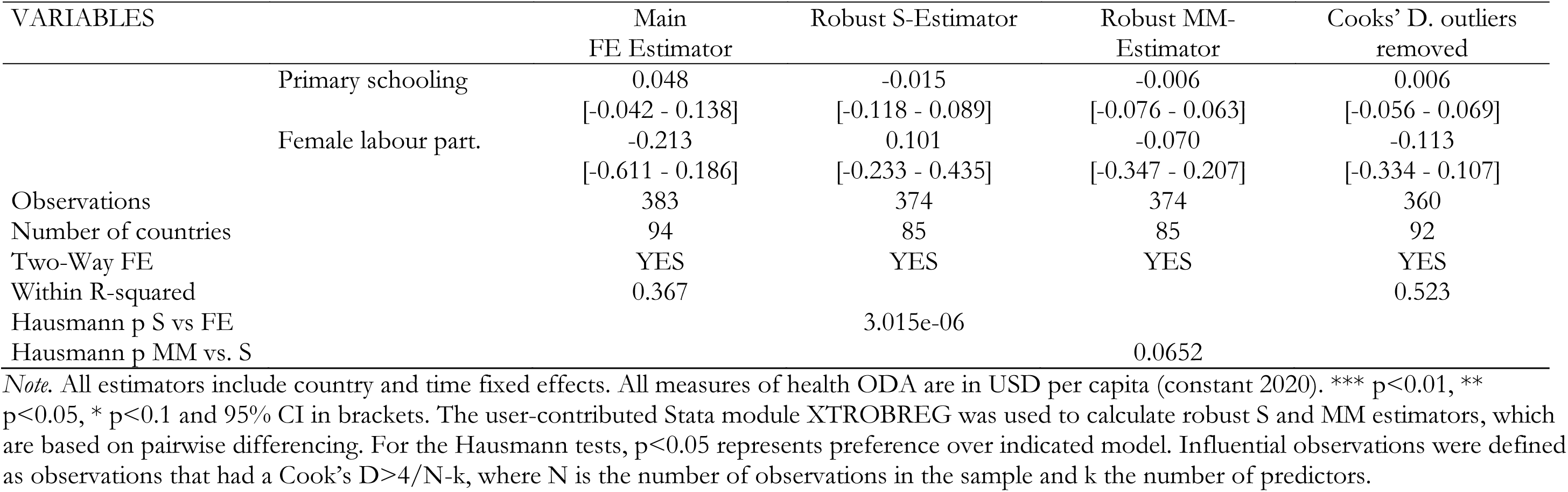
Outlier robustness analysis - Effects of sectoral ODA per capita on modern contraceptive prevalence.

| VARIABLES | Main<br>FE Estimator | Robust S-Estimator | Robust MM-Estimator | Cooks' D. outliers<br>removed |
| --- | --- | --- | --- | --- |
| **SRH ODA (USD pc)** | 0.628***<br>[0.330 - 0.926] | 0.677***<br>[0.452 - 0.902] | 0.430***<br>[0.282 - 0.577] | 0.731***<br>[0.538 - 0.923] |
| ln GDP pc | 1.735<br>[-7.796 - 11.266] | -3.663<br>[-16.008 - 8.682] | 1.497<br>[-11.260 - 14.255] | -1.234<br>[-8.119 - 5.651] |
| GHE share of CHE | 0.027<br>[-0.102 - 0.155] | 0.048<br>[-0.155 - 0.252] | 0.051<br>[-0.072 - 0.175] | 0.101**<br>[0.006 - 0.197] |
| Government effective | 3.995*<br>[-0.471 - 8.461] | 2.245<br>[-2.435 - 6.925] | 1.432<br>[-3.833 - 6.696] | 2.532<br>[-0.821 - 5.885] |
| Corruption control | 1.504<br>[-3.796 - 6.803] | -4.148<br>[-10.159 - 1.863] | -1.808<br>[-8.088 - 4.472] | -0.277<br>[-4.242 - 3.688] |
| ln Population density | 23.558**<br>[5.116 - 41.999] | 22.380***<br>[12.603 - 32.157] | 26.318***<br>[9.646 - 42.990] | 23.683***<br>[10.638 - 36.727] |
| Primary schooling | 0.064<br>[-0.017 - 0.145] | 0.017<br>[-0.082 - 0.116] | 0.010<br>[-0.058 - 0.078] | 0.027<br>[-0.038 - 0.092] |
| Female labour part. | -0.090<br>[-0.353 - 0.174] | 0.145<br>[-0.175 - 0.465] | -0.032<br>[-0.298 - 0.235] | -0.077<br>[-0.290 - 0.136] |
| Observations | 387 | 378 | 378 | 365 |
| Number of countries | 95 | 86 | 86 | 92 |
| Two-Way FE | YES | YES | YES | YES |
| Within R-squared | 0.438 |  |  | 0.576 |
| Hausmann p S vs FE |  | 0.017 |  |  |
| Hausmann p MM vs. S |  |  | 0.439 |  |
| **Total Health ODA (USD pc)** | 0.242**<br>[0.032 - 0.453] | 0.347***<br>[0.217 - 0.476] | 0.309**<br>[0.052 - 0.566] | 0.300***<br>[0.181 - 0.419] |
| ln GDP pc | 1.820<br>[-9.003 - 12.643] | 0.853<br>[-8.875 - 10.580] | 4.178<br>[-6.999 - 15.355] | 1.982<br>[-4.401 - 8.364] |
| GHE share of CHE | 0.061 | 0.060 | 0.050 | 0.113** |

|  |  | [-0.084 - 0.205] | [-0.173 - 0.294] | [-0.086 - 0.186] | [0.015 - 0.211] |
| --- | --- | --- | --- | --- | --- |
| *Table AIV.6 Continued* |  |  |  |  |  |
| VARIABLES |  | Main<br>FE Estimator | Robust S-Estimator | Robust MM-Estimator | Cooks' D. outliers<br>removed |
| Government effective |  | 3.345<br>[-1.616 - 8.305] | 2.045<br>[-1.374 - 5.464] | 1.937<br>[-1.940 - 5.813] | 2.597**<br>[0.044 - 5.150] |
| Corruption control |  | 2.372<br>[-3.543 - 8.286] | -4.130<br>[-10.275 - 2.016] | -1.220<br>[-6.175 - 3.735] | 0.107<br>[-3.842 - 4.055] |
| ln Population density |  | 21.369**<br>[0.149 - 42.589] | 20.135**<br>[3.029 - 37.242] | 25.082**<br>[5.252 - 44.911] | 26.923***<br>[14.977 - 38.869] |
| Primary schooling |  | 0.050<br>[-0.034 - 0.135] | -0.000<br>[-0.098 - 0.097] | -0.004<br>[-0.080 - 0.073] | -0.004<br>[-0.070 - 0.061] |
| Female labour part. |  | -0.149<br>[-0.456 - 0.159] | 0.062<br>[-0.223 - 0.347] | -0.087<br>[-0.353 - 0.179] | -0.107<br>[-0.305 - 0.092] |
| Observations |  | 390 | 382 | 382 | 363 |
| Number of countries |  | 96 | 88 | 88 | 92 |
| Two-Way FE |  | YES | YES | YES | YES |
| Within R-squared |  | 0.392 |  |  | 0.555 |
| Hausmann p S vs FE |  |  | 0.0149 |  |  |
| Hausmann p MM vs. S |  |  |  | 0.958 |  |
| **RH care ODA (USD pc)** |  | 0.935<br>[-1.490 - 3.361] | 0.683*<br>[-0.105 - 1.470] | 1.339<br>[-0.490 - 3.168] | 1.647**<br>[0.257 - 3.037] |
| ln GDP pc |  | -0.699<br>[-11.601 - 10.204] | -5.193<br>[-16.097 - 5.710] | 1.696<br>[-12.336 - 15.727] | -0.448<br>[-8.047 - 7.151] |
| GHE share of CHE |  | 0.039<br>[-0.098 - 0.176] | 0.018<br>[-0.133 - 0.169] | 0.061<br>[-0.071 - 0.193] | 0.100**<br>[0.003 - 0.198] |
| Government effective |  | 3.717<br>[-2.234 - 9.668] | 2.260<br>[-1.277 - 5.797] | 2.480<br>[-3.760 - 8.720] | 3.375**<br>[0.480 - 6.271] |
| Corruption control |  | 3.275<br>[-3.265 - 9.815] | -4.354*<br>[-8.720 - 0.012] | -0.374<br>[-8.332 - 7.584] | 0.863<br>[-3.216 - 4.942] |
| ln Population density |  | 21.158*<br>[-3.467 - 45.782] | 24.623***<br>[10.569 - 38.676] | 28.886**<br>[5.872 - 51.900] | 29.923***<br>[15.763 - 44.084] |
| Primary schooling | 0.048<br>[-0.042 - 0.138] | -0.015<br>[-0.118 - 0.089] | -0.006<br>[-0.076 - 0.063] | 0.006<br>[-0.056 - 0.069] |
| Female labour part. | -0.213<br>[-0.611 - 0.186] | 0.101<br>[-0.233 - 0.435] | -0.070<br>[-0.347 - 0.207] | -0.113<br>[-0.334 - 0.107] |
| Observations | 383 | 374 | 374 | 360 |
| Number of countries | 94 | 85 | 85 | 92 |
| Two-Way FE | YES | YES | YES | YES |
| Within R-squared | 0.367 |  |  | 0.523 |
| Hausmann p S vs FE |  | 3.015e-06 |  |  |
| Hausmann p MM vs. S |  |  | 0.0652 |  |

*Note.* All estimators include country and time fixed effects. All measures of health ODA are in USD per capita (constant 2020). \*\*\* $p < 0.01$ , \*\* $p < 0.05$ , \* $p < 0.1$ and 95% CI in brackets. The user-contributed Stata module XTROBREG was used to calculate robust S and MM estimators, which are based on pairwise differencing. For the Hausmann tests, $p < 0.05$ represents preference over indicated model. Influential observations were defined as observations that had a Cook's $D > 4/N-k$ , where N is the number of observations in the sample and k the number of predictors.

**Table E.7:**
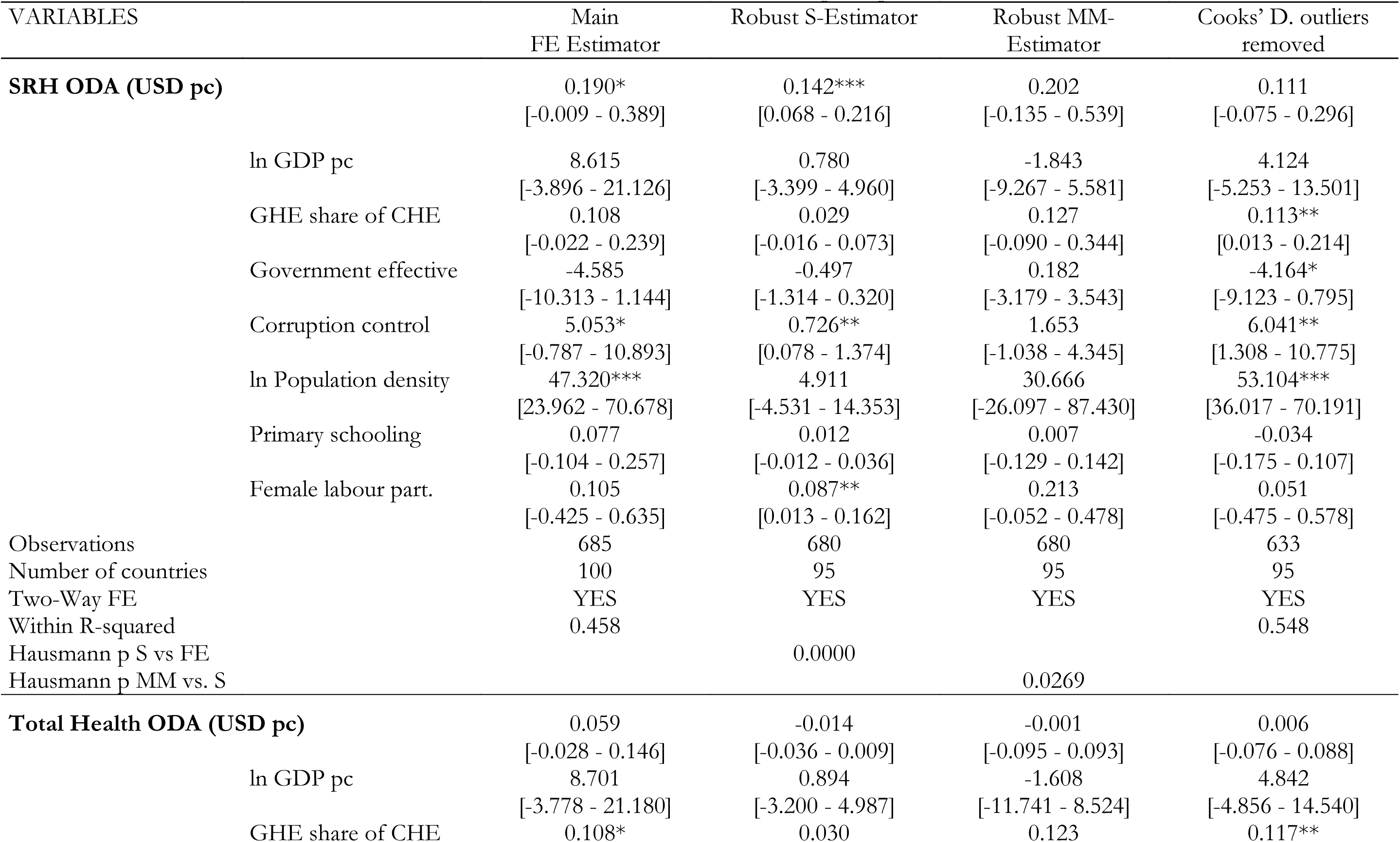
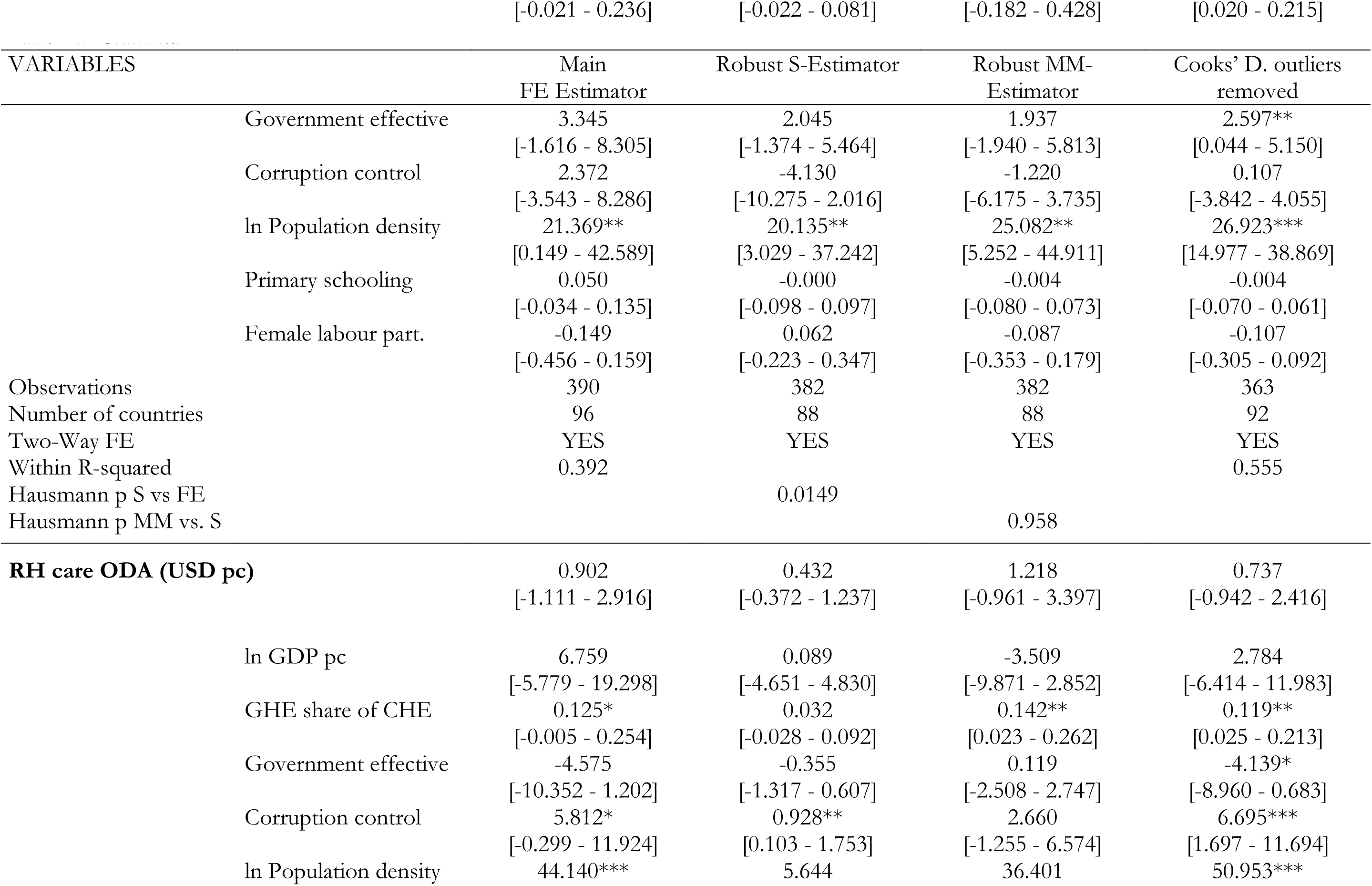
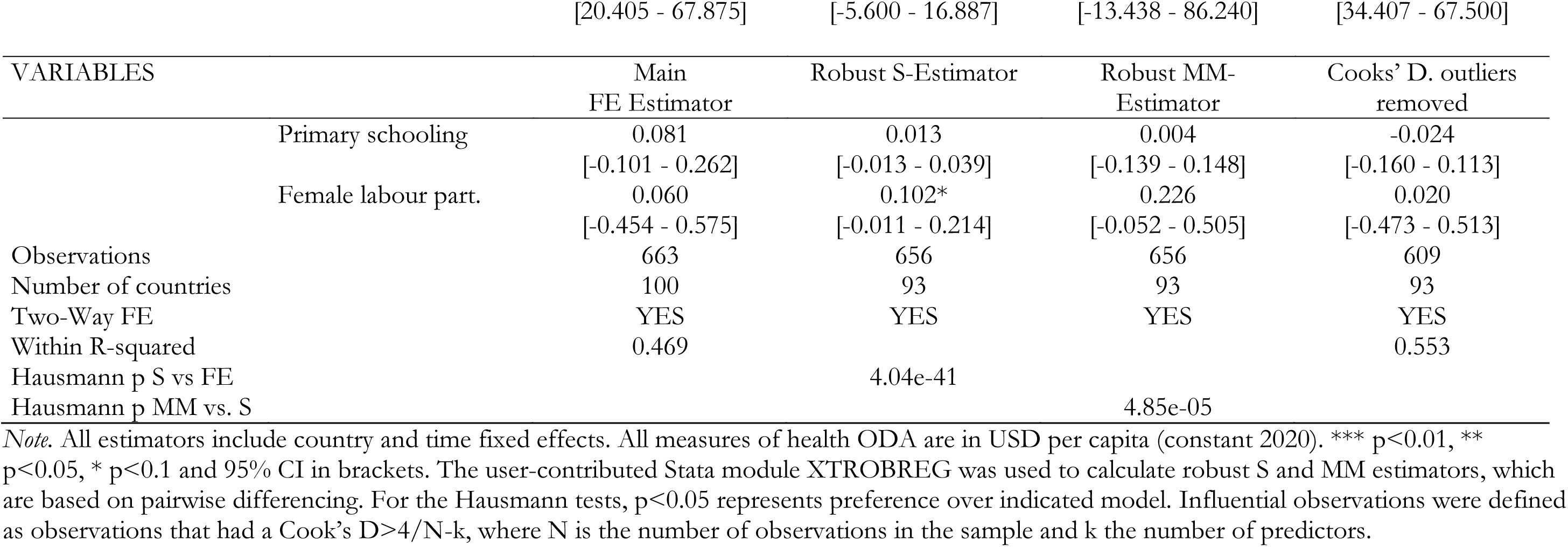
Outlier robustness analysis - Effects of sectoral ODA per capita on skilled birth attendance.

**Table E.8:**
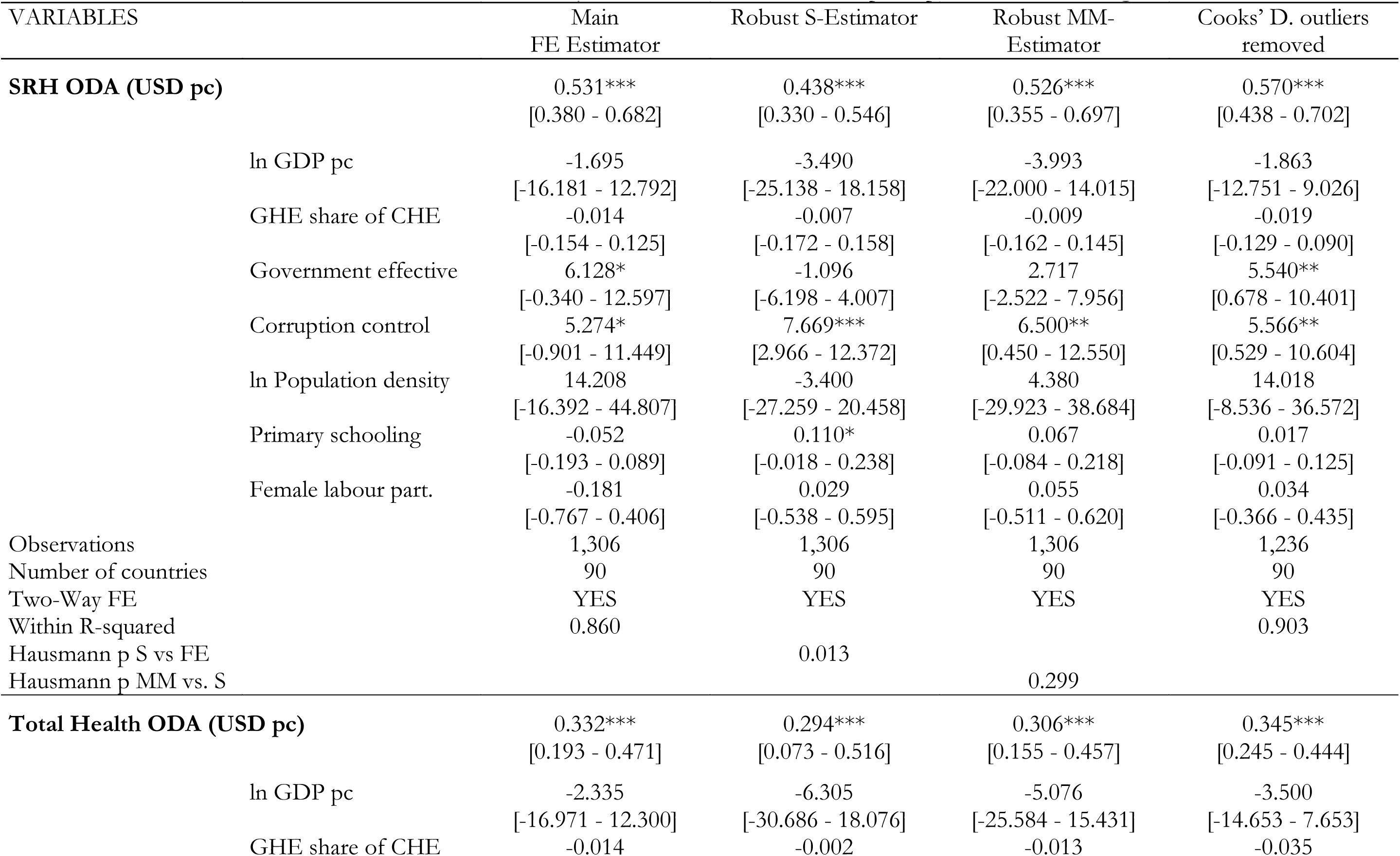
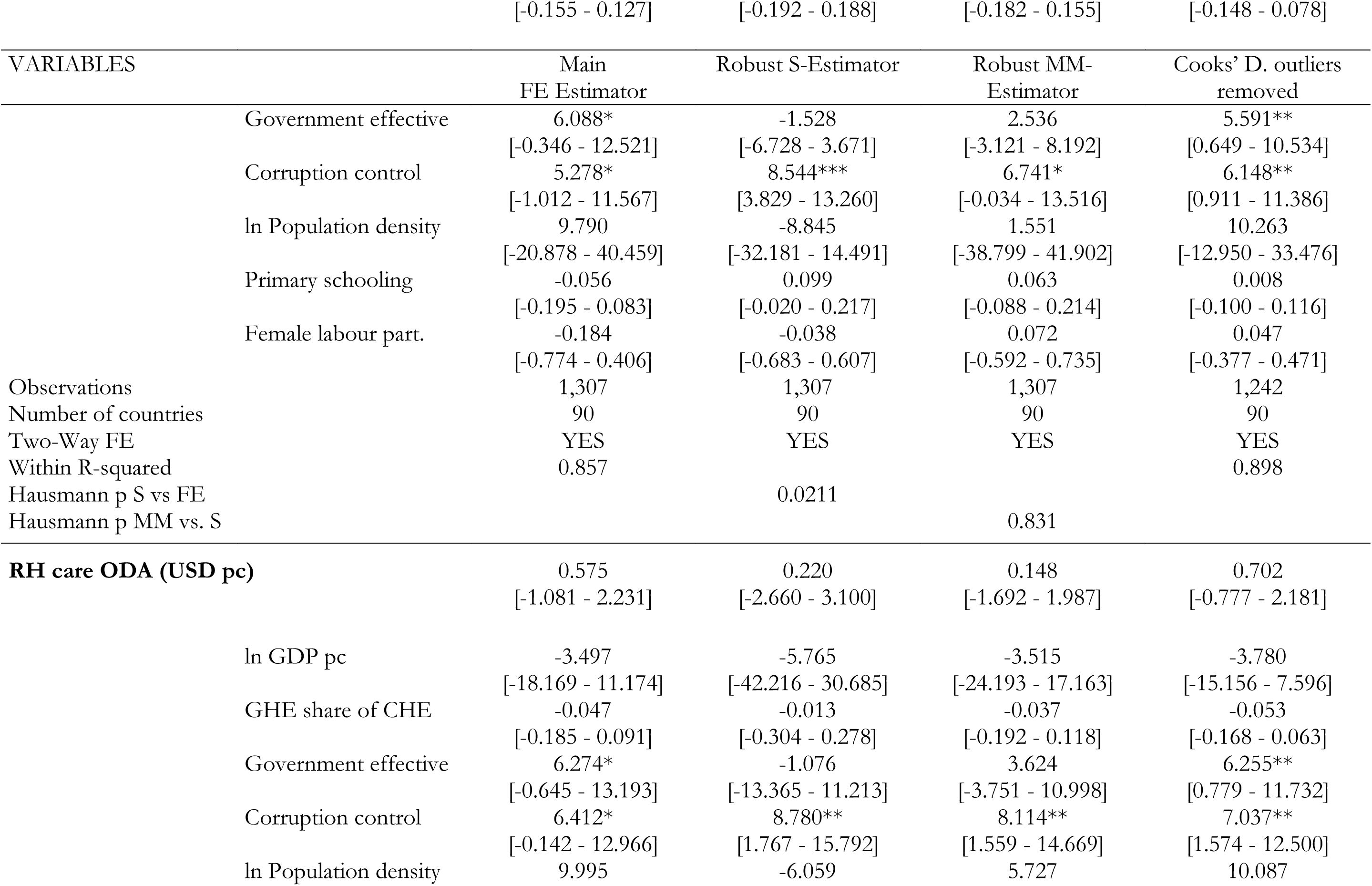
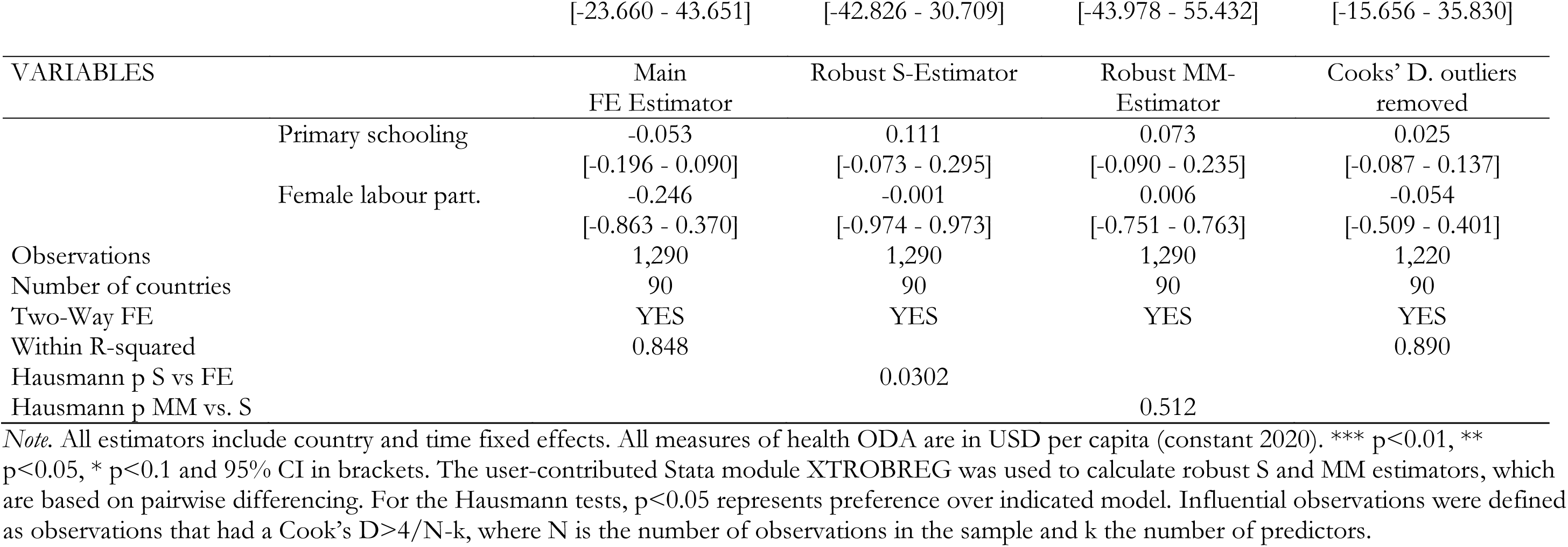
Outlier robustness analysis - Effects of sectoral ODA per capita on ART coverage.

### E.3: FE estimation with alternative functional forms

**Table E.9:** The effects of health ODA on SRH services’ summary of FE estimations with alternative functional form.

| Sectoral ODA | SRH service outcomes |  |  |
| --- | --- | --- | --- |
|  | Contraceptive Prevalence | Skilled birth attendance | ART coverage |
| **SRH ODA** |  |  |  |
| Main FE estimator (linear, USD pc) | 0.628*** | 0.190* | 0.531*** |
| FE estimator (linear-log, USD millions) | 1.904** | 0.239 | 1.121 |
| FE estimator (log-log, USD millions) | 0.0882*** | 0.0149 | 0.0626 |
| **Total Health ODA** |  |  |  |
| Main FE estimator (linear, USD pc) | 0.242** | 0.0591 | 0.332*** |
| FE estimator (linear-log, USD millions) | 1.203 | -0.224 | 2.004* |
| FE estimator (log-log, USD millions) | 0.0606 | 0.00189 | 0.136*** |
| **RH care ODA** |  |  |  |
| Main FE estimator (linear, USD pc) | 0.935 | 0.902 | 0.575 |
| FE estimator (linear-log, USD millions) | 1.676*** | 0.830** | 0.711 |
| FE estimator (log-log, USD millions) | 0.0706*** | 0.0265*** | 0.0134 |

*Note.* All estimations include country and time fixed effects. Standard errors are clustered at the country level. \*\*\* p<0.01, \*\* p<0.05, \* p<0.1. In the linear-log specification, all IVs are log-transformed, including ODA. The linear-log specification provides the absolute change in service coverage (percentage points) associated with a per cent change in ODA. In the log-log specification, all variables are log-transformed, including SRH service indicators. The log-log specification allows coefficients to be interpreted as elasticities, thus the percent change in service coverage associated with a percent change in ODA.

**Table E.10:**
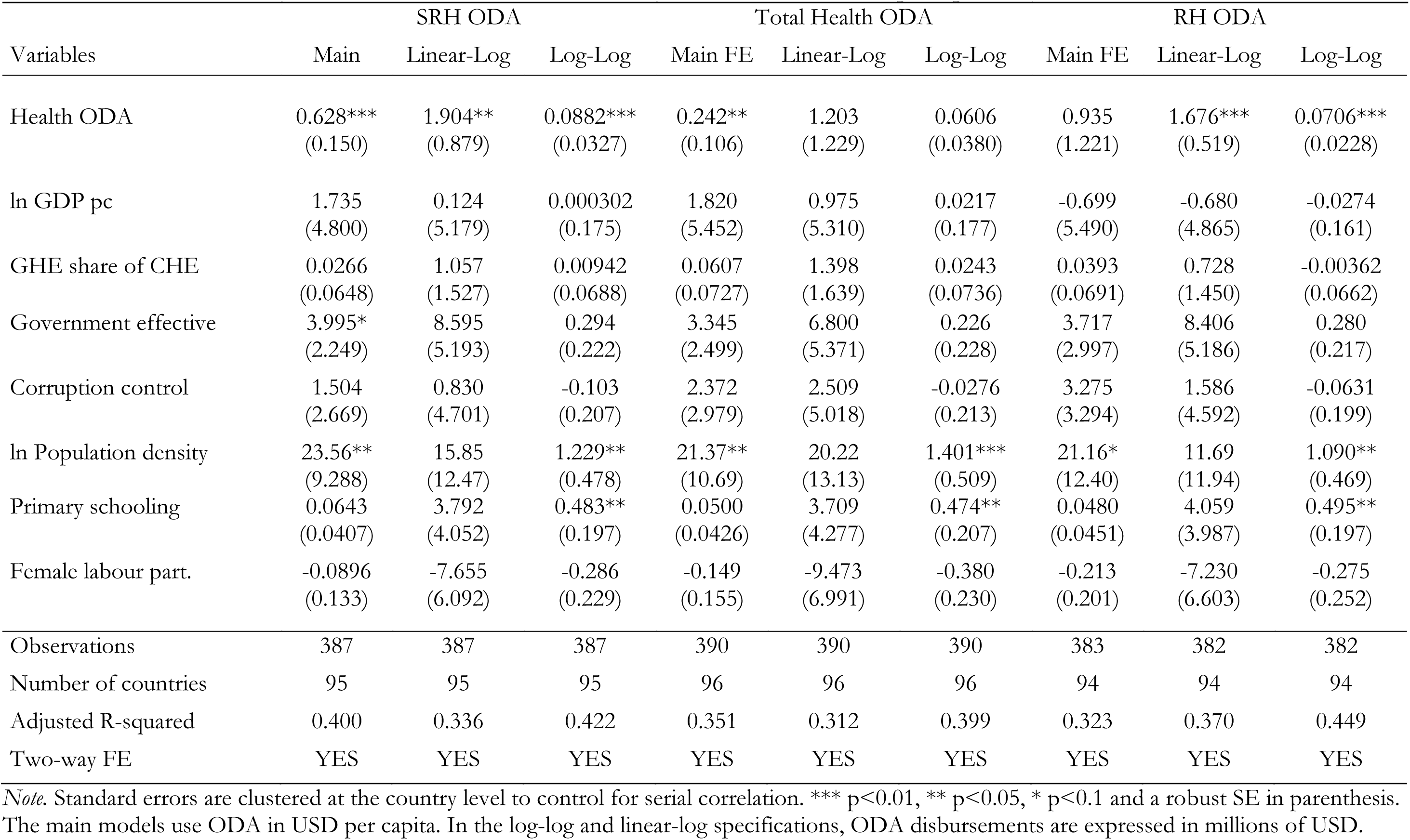
Effects of sectoral ODA on modern contraceptive prevalence.

**Table E.11:** Effects of sectoral ODA on skilled birth attendance.

| Variables | SRH ODA |  |  | Total Health ODA |  |  | RH ODA |  |  |
| --- | --- | --- | --- | --- | --- | --- | --- | --- | --- |
|  | Main | Linear-Log | Log-Log | Main | Linear-Log | Log-Log | Main | Linear-Log | Log-Log |
| Health ODA | 0.190*<br>(0.100) | 0.239<br>(0.501) | 0.0149<br>(0.00991) | 0.0591<br>(0.0439) | -0.224<br>(0.557) | 0.00189<br>(0.0117) | 0.902<br>(1.015) | 0.830**<br>(0.418) | 0.0265***<br>(0.00954) |
| ln GDP pc | 8.615<br>(6.305) | 9.247<br>(6.422) | 0.280*<br>(0.147) | 8.701<br>(6.290) | 9.950<br>(6.308) | 0.291**<br>(0.145) | 6.759<br>(6.319) | 7.762<br>(6.308) | 0.261*<br>(0.144) |
| GHE share of CHE | 0.108<br>(0.0658) | 3.175<br>(2.180) | -0.0351<br>(0.0471) | 0.108*<br>(0.0648) | 3.188<br>(2.161) | -0.0350<br>(0.0469) | 0.125*<br>(0.0653) | 3.141<br>(2.193) | -0.0414<br>(0.0471) |
| Government effective | -4.585<br>(2.887) | -7.845<br>(5.570) | -0.181<br>(0.110) | -3.935<br>(2.698) | -7.186<br>(5.306) | -0.168<br>(0.105) | -4.575<br>(2.911) | -7.909<br>(5.708) | -0.187<br>(0.113) |
| Corruption control | 5.053*<br>(2.943) | 6.842<br>(4.963) | 0.169*<br>(0.0923) | 4.772*<br>(2.704) | 6.323<br>(4.551) | 0.162*<br>(0.0856) | 5.812*<br>(3.080) | 7.729<br>(5.208) | 0.183*<br>(0.0979) |
| ln Population density | 47.32***<br>(11.77) | 46.79***<br>(11.70) | 1.010***<br>(0.276) | 48.15***<br>(11.49) | 49.33***<br>(11.15) | 1.067***<br>(0.270) | 44.14***<br>(11.96) | 41.49***<br>(11.72) | 0.897***<br>(0.283) |
| Primary schooling | 0.0767<br>(0.0911) | 6.031<br>(8.130) | 0.299*<br>(0.178) | 0.0756<br>(0.0901) | 6.187<br>(8.101) | 0.307*<br>(0.178) | 0.0809<br>(0.0914) | 5.996<br>(8.167) | 0.299*<br>(0.178) |
| Female labour part. | 0.105<br>(0.267) | 6.503<br>(10.33) | 0.300<br>(0.233) | 0.106<br>(0.267) | 6.793<br>(10.18) | 0.294<br>(0.235) | 0.0605<br>(0.259) | 7.007<br>(10.46) | 0.324<br>(0.230) |
| Observations | 685 | 685 | 685 | 695 | 695 | 695 | 663 | 662 | 662 |
| Number of countries | 100 | 100 | 100 | 101 | 101 | 101 | 100 | 100 | 100 |
| Adjusted R-squared | 0.437 | 0.431 | 0.382 | 0.432 | 0.428 | 0.378 | 0.448 | 0.450 | 0.403 |
| Two-way FE | YES | YES | YES | YES | YES | YES | YES | YES | YES |

*Note.* Standard errors are clustered at the country level to control for serial correlation. \*\*\* p<0.01, \*\* p<0.05, \* p<0.1 and a robust SE in parenthesis. The main models use ODA in USD per capita. In the log-log and linear-log specifications, ODA disbursements are expressed in millions of USD.

**Table E.12:** Effects of sectoral ODA on ART coverage.

| Variables | SRH ODA |  |  | Total Health ODA |  |  | RH ODA |  |  |
| --- | --- | --- | --- | --- | --- | --- | --- | --- | --- |
|  | Main | Linear-Log | Log-Log | Main | Linear-Log | Log-Log | Main | Linear-Log | Log-Log |
| Health ODA | 0.531***<br>(0.0761) | 1.121<br>(1.029) | 0.0626<br>(0.0442) | 0.332***<br>(0.0698) | 2.004*<br>(1.110) | 0.136***<br>(0.0510) | 0.575<br>(0.833) | 0.711<br>(0.490) | 0.0134<br>(0.0208) |
| ln GDP pc | -1.695<br>(7.291) | -3.943<br>(7.532) | 0.740**<br>(0.334) | -2.335<br>(7.366) | -4.216<br>(7.637) | 0.703**<br>(0.316) | -3.497<br>(7.384) | -3.940<br>(7.441) | 0.761**<br>(0.336) |
| GHE share of CHE | -0.0144<br>(0.0701) | -2.271<br>(1.455) | -0.0749<br>(0.1000) | -0.0140<br>(0.0709) | -2.193<br>(1.498) | -0.0676<br>(0.0988) | -0.0471<br>(0.0695) | -2.351<br>(1.471) | -0.0724<br>(0.100) |
| Government effective | 6.128*<br>(3.255) | 12.46**<br>(5.423) | 0.308<br>(0.292) | 6.088*<br>(3.238) | 11.98**<br>(5.414) | 0.308<br>(0.288) | 6.274*<br>(3.482) | 12.56**<br>(5.584) | 0.295<br>(0.297) |
| Corruption control | 5.274*<br>(3.108) | 8.374<br>(5.609) | 0.221<br>(0.222) | 5.278*<br>(3.165) | 8.768<br>(5.626) | 0.234<br>(0.224) | 6.412*<br>(3.298) | 9.225*<br>(5.548) | 0.239<br>(0.229) |
| ln Population density | 14.21<br>(15.40) | 6.466<br>(16.56) | 1.334<br>(0.807) | 9.790<br>(15.43) | 5.405<br>(16.42) | 1.228*<br>(0.734) | 9.995<br>(16.94) | 6.760<br>(16.20) | 1.483*<br>(0.792) |
| Primary schooling | -0.0522<br>(0.0711) | -4.625<br>(6.540) | 0.873**<br>(0.357) | -0.0563<br>(0.0699) | -5.335<br>(6.503) | 0.797**<br>(0.339) | -0.0529<br>(0.0720) | -4.418<br>(6.640) | 0.897**<br>(0.367) |
| Female labour part. | -0.181<br>(0.295) | -15.54<br>(15.64) | -1.182<br>(0.768) | -0.184<br>(0.297) | -15.21<br>(15.40) | -1.129<br>(0.758) | -0.246<br>(0.310) | -16.36<br>(15.73) | -1.181<br>(0.773) |
| Observations | 1,306 | 1,306 | 1,159 | 1,307 | 1,307 | 1,160 | 1,290 | 1,289 | 1,143 |
| Number of countries | 90 | 90 | 90 | 90 | 90 | 90 | 90 | 90 | 90 |
| Adjusted R-squared | 0.857 | 0.847 | 0.875 | 0.854 | 0.848 | 0.876 | 0.845 | 0.846 | 0.875 |
| Two-way FE | YES | YES | YES | YES | YES | YES | YES | YES | YES |

*Note.* Standard errors are clustered at the country level to control for serial correlation. \*\*\* p<0.01, \*\* p<0.05, \* p<0.1 and a robust SE in parenthesis. The main models use ODA in USD per capita. In the log-log and linear-log specifications, ODA disbursements are expressed in millions of USD.

### E.4: Potential delayed effect

**Table E.13:**
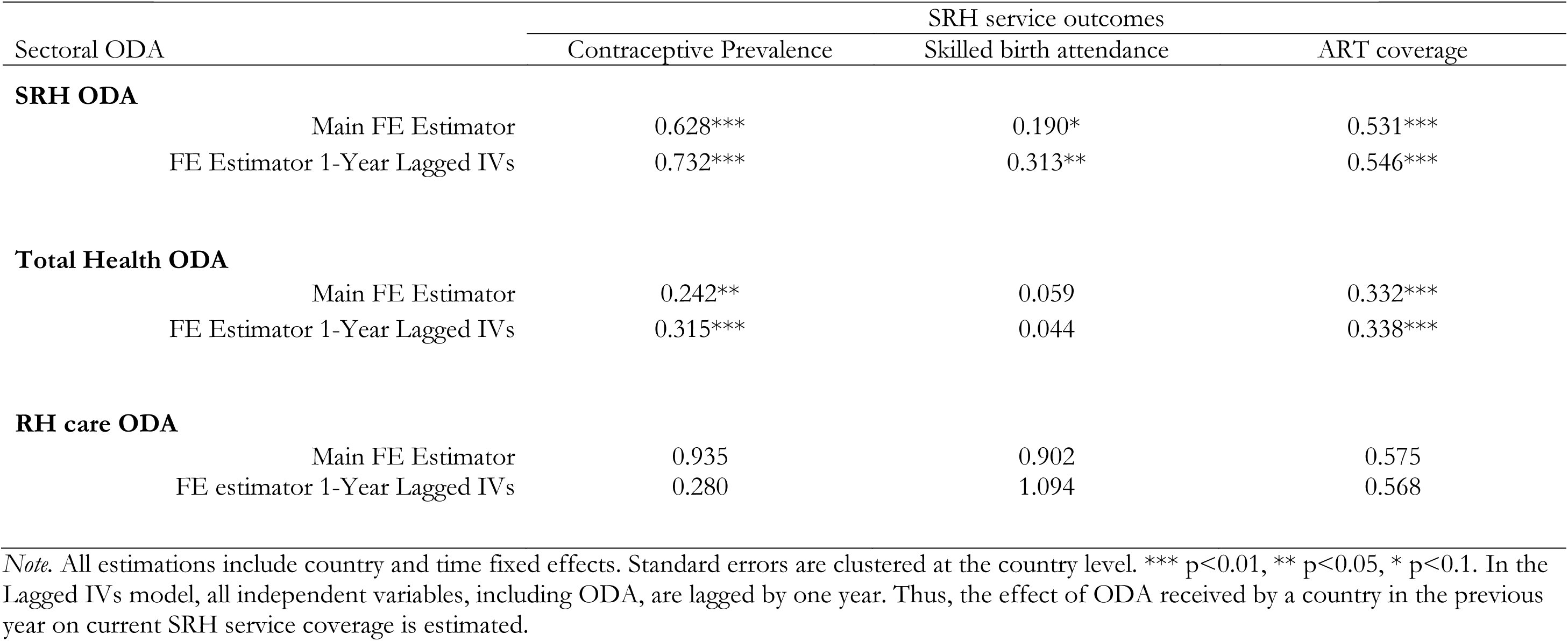
The effects of health ODA on SRH services’ summary of lagged IVs (including ODA) models.

| Sectoral ODA |  | SRH service outcomes |  |  |
| --- | --- | --- | --- | --- |
|  |  | Contraceptive Prevalence | Skilled birth attendance | ART coverage |
| SRH ODA |  |  |  |  |
|  | Main FE Estimator | 0.628*** | 0.190* | 0.531*** |
|  | FE Estimator 1-Year Lagged IVs | 0.732*** | 0.313** | 0.546*** |
| Total Health ODA |  |  |  |  |
|  | Main FE Estimator | 0.242** | 0.059 | 0.332*** |
|  | FE Estimator 1-Year Lagged IVs | 0.315*** | 0.044 | 0.338*** |
| RH care ODA |  |  |  |  |
|  | Main FE Estimator | 0.935 | 0.902 | 0.575 |
|  | FE estimator 1-Year Lagged IVs | 0.280 | 1.094 | 0.568 |

*Note.* All estimations include country and time fixed effects. Standard errors are clustered at the country level. \*\*\* p<0.01, \*\* p<0.05, \* p<0.1. In the Lagged IVs model, all independent variables, including ODA, are lagged by one year. Thus, the effect of ODA received by a country in the previous year on current SRH service coverage is estimated.

**Table E.14:** FE estimation - Effects of health ODA per capita on modern contraceptive prevalence.

| VARIABLES | SRH ODA |  | TotH ODA |  | RH ODA |  |
| --- | --- | --- | --- | --- | --- | --- |
|  | Main Model | Lagged IVs | Main Model | Lagged IVs | Main Model | Lagged IVs |
| Health ODA | 0.628***<br>(0.150) | 0.732***<br>(0.201) | 0.242**<br>(0.106) | 0.315***<br>(0.099) | 0.935<br>(1.221) | 0.280<br>(0.758) |
| ln GDP pc | 1.735<br>(4.800) | 1.398<br>(4.367) | 1.820<br>(5.452) | -1.509<br>(5.109) | -0.699<br>(5.490) | -1.586<br>(5.333) |
| GHE share of CHE | 0.027<br>(0.065) | 0.049<br>(0.073) | 0.061<br>(0.073) | 0.034<br>(0.074) | 0.039<br>(0.069) | 0.024<br>(0.077) |
| Government effective | 3.995*<br>(2.249) | 5.068**<br>(2.304) | 3.345<br>(2.499) | 5.347**<br>(2.539) | 3.717<br>(2.997) | 4.420<br>(3.481) |
| Corruption control | 1.504<br>(2.669) | 2.812<br>(1.979) | 2.372<br>(2.979) | 3.330<br>(2.238) | 3.275<br>(3.294) | 3.746<br>(2.868) |
| ln Population density | 23.558**<br>(9.288) | 26.206**<br>(10.793) | 21.369**<br>(10.689) | 19.811<br>(12.352) | 21.158*<br>(12.400) | 16.887<br>(16.439) |
| Primary schooling | 0.064<br>(0.041) | 0.057<br>(0.046) | 0.050<br>(0.043) | 0.052<br>(0.047) | 0.048<br>(0.045) | 0.059<br>(0.051) |
| Female labour part. | -0.090<br>(0.133) | 0.044<br>(0.144) | -0.149<br>(0.155) | -0.037<br>(0.161) | -0.213<br>(0.201) | -0.163<br>(0.251) |
| Observations | 387 | 376 | 390 | 377 | 383 | 372 |
| Number of countries | 95 | 97 | 96 | 98 | 94 | 97 |
| Two-way FE | YES | YES | YES | YES | YES | YES |
| Adj. within R-squared | 0.400 | 0.444 | 0.351 | 0.414 | 0.323 | 0.337 |
| Rho | 0.980 | 0.984 | 0.976 | 0.978 | 0.977 | 0.970 |

*Note.* All measures of health ODA are in USD per capita (constant 2020). Robust standard errors in parentheses. \*\*\* p<0.01, \*\* p<0.05, \* p<0.1 In the Lagged IVs model, all independent variables, including ODA, are lagged by one year.

**Table E.15:** FE estimation - Effects of health ODA per capita on skilled birth attendance.

| VARIABLES | SRH ODA |  | TotH ODA |  | RH ODA |  |
| --- | --- | --- | --- | --- | --- | --- |
|  | Main Model | Lagged IVs | Main Model | Lagged IVs | Main Model | Lagged IVs |
| Health ODA | 0.190*<br>(0.100) | 0.313**<br>(0.158) | 0.059<br>(0.044) | 0.044<br>(0.062) | 0.902<br>(1.015) | 1.094<br>(0.719) |
| ln GDP pc | 8.615<br>(6.305) | 11.603*<br>(6.656) | 8.701<br>(6.290) | 11.357*<br>(6.662) | 6.759<br>(6.319) | 10.957<br>(6.843) |
| GHE share of CHE | 0.108<br>(0.066) | 0.103<br>(0.068) | 0.108*<br>(0.065) | 0.092<br>(0.067) | 0.125*<br>(0.065) | 0.098<br>(0.067) |
| Government effective | -4.585<br>(2.887) | -5.018**<br>(2.450) | -3.935<br>(2.698) | -4.554*<br>(2.409) | -4.575<br>(2.911) | -5.213**<br>(2.605) |
| Corruption control | 5.053*<br>(2.943) | 5.701**<br>(2.666) | 4.772*<br>(2.704) | 5.365**<br>(2.584) | 5.812*<br>(3.080) | 6.346**<br>(2.882) |
| ln Population density | 47.320***<br>(11.772) | 55.020***<br>(12.280) | 48.155***<br>(11.492) | 55.040***<br>(12.257) | 44.140***<br>(11.962) | 51.193***<br>(13.486) |
| Primary schooling | 0.077<br>(0.091) | 0.107<br>(0.097) | 0.076<br>(0.090) | 0.112<br>(0.096) | 0.081<br>(0.091) | 0.118<br>(0.098) |
| Female labour part. | 0.105<br>(0.267) | 0.052<br>(0.255) | 0.106<br>(0.267) | 0.028<br>(0.253) | 0.060<br>(0.259) | -0.023<br>(0.255) |
| Observations | 685 | 665 | 695 | 670 | 663 | 641 |
| Number of countries | 100 | 101 | 101 | 101 | 100 | 101 |
| Two-way FE | YES | YES | YES | YES | YES | YES |
| Adj. within R-squared | 0.437 | 0.488 | 0.432 | 0.476 | 0.448 | 0.491 |
| Rho | 0.987 | 0.991 | 0.988 | 0.991 | 0.986 | 0.989 |

*Note.* All measures of health ODA are in USD per capita (constant 2020). Robust standard errors in parentheses. \*\*\* p<0.01, \*\* p<0.05, \* p<0.1. In the Lagged IVs model, all independent variables, including ODA, are lagged by one year.

**Table E.16:** FE estimation - Effects of health ODA per capita on ART coverage.

| VARIABLES | SRH ODA |  | TotH ODA |  | RH ODA |  |
| --- | --- | --- | --- | --- | --- | --- |
|  | Main Model | Lagged IVs | Main Model | Lagged IVs | Main Model | Lagged IVs |
| Health ODA | 0.531***<br>(0.076) | 0.546***<br>(0.081) | 0.332***<br>(0.070) | 0.338***<br>(0.071) | 0.575<br>(0.833) | 0.568<br>(0.773) |
| ln GDP pc | -1.695<br>(7.291) | -1.030<br>(7.359) | -2.335<br>(7.366) | -1.729<br>(7.445) | -3.497<br>(7.384) | -2.828<br>(7.443) |
| GHE share of CHE | -0.014<br>(0.070) | -0.027<br>(0.071) | -0.014<br>(0.071) | -0.028<br>(0.071) | -0.047<br>(0.070) | -0.060<br>(0.070) |
| Government effective | 6.128*<br>(3.255) | 6.772**<br>(3.341) | 6.088*<br>(3.238) | 6.772**<br>(3.328) | 6.274*<br>(3.482) | 6.951*<br>(3.579) |
| Corruption control | 5.274*<br>(3.108) | 5.634*<br>(3.001) | 5.278*<br>(3.165) | 5.655*<br>(3.051) | 6.412*<br>(3.298) | 6.770**<br>(3.206) |
| ln Population density | 14.208<br>(15.400) | 18.655<br>(15.619) | 9.790<br>(15.435) | 14.211<br>(15.640) | 9.995<br>(16.938) | 14.155<br>(17.248) |
| Primary schooling | -0.052<br>(0.071) | -0.044<br>(0.073) | -0.056<br>(0.070) | -0.048<br>(0.072) | -0.053<br>(0.072) | -0.043<br>(0.075) |
| Female labour part. | -0.181<br>(0.295) | -0.224<br>(0.305) | -0.184<br>(0.297) | -0.230<br>(0.305) | -0.246<br>(0.310) | -0.290<br>(0.319) |
| Observations | 1,306 | 1,306 | 1,307 | 1,307 | 1,290 | 1,290 |
| Number of countries | 90 | 90 | 90 | 90 | 90 | 90 |
| Two-way FE | YES | YES | YES | YES | YES | YES |
| Adj. within R-squared | 0.857 | 0.867 | 0.854 | 0.864 | 0.845 | 0.855 |
| Rho | 0.897 | 0.929 | 0.850 | 0.900 | 0.861 | 0.904 |

*Note.* All measures of health ODA are in USD per capita (constant 2020). Robust standard errors in parentheses. \*\*\* p<0.01, \*\* p<0.05, \* p<0.1. In the Lagged IVs model, all independent variables, including ODA, are lagged by one year.

### E.5: Dynamic FE model – System GMM

The relationship between service coverage and health ODA may be dynamic, meaning that previous SRH services coverage might be correlated with contemporary ODA allocation decisions. To account for the potential endogeneity of ODA (and covariates) due to this dynamic effect, we used a two-step system GMM estimator with country and time fixed effects on our data (Frees 2004, Cameron and Trivedi 2005). For this estimation, we used the community-contributed xtdpdgmm command in Stata (Kripfganz 2020).

The system-GMM estimator was applied to the three-year averaged data with six time periods, to limit bias from missing data and measurement error. Based on their suggested relevance from the main models, the following covariates were included in the system-GMM models; ln GDP pc, GHE/CHE, corruption control, ln population density, and primary school enrollment.

As a starting point, ODA was considered predetermined, and all other covariates were considered endogenous. Lagged forward-orthogonal-deviations (fod) of these variables were used as instruments for the current levels of the regressors. While first-differences (diff) are most commonly used, the fod-GMM estimator retains more information than the diff-GMM estimators in unbalanced panels (Kripfganz 2020), such as ours. Our initial candidate model included all available instruments for the fod-transformed models. The two-step system-GMM was implemented with robust and Windmeijer corrected standard errors and the instrument set was collapsed, to reduce the number of instruments. This was necessary given the relatively small number of countries (N) in our panel.

For each system-GMM model estimated, a Hansen test for overidentifying restrictions was conducted as well as AR(1) and AR(2) tests for first and second-order autocorrelation, respectively. Moreover, the Akaike (AIC) and Bayesian (BIC) information criteria were computed to compare candidate models and support the specification search. Specifications of individual models were systematically adjusted (from the baseline choice) to fit the stringent identifying assumptions. For instance, we experimented with including a different number of instruments (e.g., limiting instruments to two-period lags). Also, we tested the models when classifying some of the covariates as pre-determined, and even exogenous.

In line with the main FE estimation, the results from the system-GMM estimation generally suggested positive correlation between sectoral health ODA and SRH services. However, the magnitude and significance of these correlations were highly sensitive to model specifications and variable classifications. The GMM modelling approach was designed to estimate models with relatively few time periods and large panels (N) and requires quite a large number of observations to provide robust estimates. Thus, it may not be ideal for our country-level panel data. The reduction in potential endogeneity bias from using the system-GMM estimator may come at the cost of loss in precision (Frees 2004).

## Footnotes

1 For example, target 3.1 – Reduce maternal mortality, target 3.7 – Ensuring universal access to sexual and reproductive care, family planning and education, and target 5.6 – Ensuring universal access to sexual and reproductive health and reproductive rights.

2 In this report we use the terms ODA, aid, development assistance and external assistance interchangeably.

3 See https://datahelpdesk.worldbank.org/knowledgebase/articles/906519 for details.

4 The five three-year periods being 2003-2005, 2006-2008, 2009-2011, 2012-2014, 2015-2017, and 2018-2020.

5 https://www.state.gov/wp-content/uploads/2019/12/Malawi-SID-2019.pdf

## Notes

### Competing Interest Statement

The authors have declared no competing interest.

### Funding Statement

This study was funded by the Swedish public agency The Expert Group for Aid Studies.

## References

Addison, T., et al. (2005). “Development assistance and development finance: evidence and global policy agendas.” Journal of International Development 17(6): 819–836.

Akinlo, A. E. and A. O. Sulola (2019). “Health care expenditure and infant mortality in sub-Saharan Africa.” Journal of Policy Modeling 41(1): 168–178.

Arndt, C., et al. (2015). “Assessing Foreign Aid’s Long-Run Contribution to Growth and Development.” World Development 69: 6–18.

Arsenault, C., et al. (2020). “Hospital-provision of essential primary care in 56 countries: determinants and quality.” Bulletin of the World Health Organization 98(11): 735–746D.

Baltagi, B. H. (2021). Econometric analysis of panel data, Springer Cham.

Banchani, E. and L. Swiss (2019). “The Impact of Foreign Aid on Maternal Mortality.” Politics and Governance 7(2): 53–67.

Bendavid, E., et al. (2010). “The relation of price of antiretroviral drugs and foreign assistance with coverage of HIV treatment in Africa: retrospective study.” Bmj 341: c6218.

Biscaye, P. E., et al. (2017). “Relative Effectiveness of Bilateral and Multilateral Aid on Development Outcomes.” Review of Development Economics 21(4): 1425–1447.

Brooks, N., et al. (2019). “USA aid policy and induced abortion in sub-Saharan Africa: an analysis of the Mexico City Policy.” Lancet Glob Health 7(8): e1046–e1053.

Chansa, C., et al. (2008). “Exploring SWAp’s contribution to the efficient allocation and use of resources in the health sector in Zambia.” Health Policy Plan 23(4): 244–251.

Coburn, C., et al. (2017). “The World Bank, Organized Hypocrisy, and Women’s Health: A Cross-National Analysis of Maternal Mortality in Sub-Saharan Africa.” Sociological Forum 32(1): 50–71.

Dickson, K. S., et al. (2021). “Women empowerment and skilled birth attendance in sub-Saharan Africa: A multi-country analysis.” PLOS ONE 16(7): e0254281.

DSW and European Parliamentary Forum (2020). Donors delivering for SRHR: tracking OECD donor funding for sexual and reproductive health and rights.

Friberg, I. K., et al. (2017). “A method for estimating maternal and newborn lives saved from health-related investments funded by the UK government Department for International Development using the Lives Saved Tool.” BMC Public Health 17(4): 779.

Grépin, K. A. (2012). “HIV donor funding has both boosted and curbed the delivery of different non-HIV health services in sub-Saharan Africa.” Health Aff (Millwood) 31(7): 1406–1414.

Hanlon, M., et al. (2012). “Exploring the relationship between population density and maternal health coverage.” BMC Health Services Research 12(1): 416.

Herzer, D. and P. Nunnenkamp (2012). “The effect of foreign aid on income inequality: Evidence from panel cointegration.” Structural Change and Economic Dynamics 23(3): 245–255.

Hsiao, A. J. and C. A. Emdin (2015). “The association between development assistance for health and malaria, HIV and tuberculosis mortality: a cross-national analysis.” J Epidemiol Glob Health 5(1): 41–48.

Islam, M. N. (2003). “Political Regimes and the Effects of Foreign Aid on Economic Growth.” The Journal of Developing Areas 37(1): 35–53.

Kaiser, A. H., et al. (2021). “The cost-effectiveness of sexual and reproductive health and rights interventions in low- and middle-income countries: a scoping review.” Sexual and Reproductive Health Matters 29(1): 90–103.

Khatiwada, J., et al. (2020). “Dimensions of women’s empowerment on access to skilled delivery services in Nepal.” BMC Pregnancy and Childbirth 20(1).

Khorrami, N., et al. (2019). “An overview of advances in global maternal health: From broad to specific improvements.” International Journal of Gynecology & Obstetrics 146(1): 126–131.

Kotsadam, A., et al. (2018). “Development aid and infant mortality. Micro-level evidence from Nigeria.” World Development 105: 59–69.

Lee, H.-Y., et al. (2016). “Control of corruption, democratic accountability, and effectiveness of HIV/AIDS official development assistance.” Global Health Action 9(1): 30306.

Makuta, I. and B. O’Hare (2015). “Quality of governance, public spending on health and health status in Sub Saharan Africa: a panel data regression analysis.” BMC Public Health 15(1).

Marty, R., et al. (2017). “Taking the health aid debate to the subnational level: the impact and allocation of foreign health aid in Malawi.” BMJ Global Health 2(1): e000129.

Moral-Benito, E., et al. (2019). “Dynamic panel data modelling using maximum likelihood: an alternative to Arellano-Bond.” Applied Economics 51(20): 2221–2232.

Niño-Zarazúa, M., et al. (2020). Effects of Swedish and International Democracy Aid, Expertgruppen för biståndsanalys (EBA).

Novignon, J., et al. (2012). “The effects of public and private health care expenditure on health status in sub-Saharan Africa: new evidence from panel data analysis.” Health Economics Review 2(1): 22.

Nunnenkamp, P. and H. Öhler (2011). “Throwing Foreign Aid at HIV/AIDS in Developing Countries: Missing the Target?” World Development 39(10): 1704–1723.

Olakunde, B. O., et al. (2019). “Revisiting aid dependency for HIV programs in Sub-Saharan Africa.” Public Health 170: 57–60.

Pallas, S. W. and J. P. Ruger (2017). ”Does donor proliferation in development aid for health affect health service delivery and population health? Cross-country regression analysis from 1995 to 2010.” Health Policy Plan 32(4): 493–503.

Pickbourn, L. and L. Ndikumana (2016). “The Impact of the Sectoral Allocation of Foreign aid on Gender Inequality.” Journal of International Development 28(3): 396–411.

Piemonte, C. (2020). External financing to Least Developed Countries (LDCs): where we stand. Factsheet. Paris, OECD.

Ravindran, T. K. S. and V. Govender (2020). “Sexual and reproductive health services in universal health coverage: a review of recent evidence from low- and middle-income countries.” Sexual and Reproductive Health Matters 28(2): 1779632.

Requejo, J., et al. (2020). “Delivering for women, children, and adolescents during and after covid-19 [Internet]. BMJ Opinion; 2020 [cited 2022 Feb 27]. Available from: https://blogs.bmj.com/bmj/2020/09/23/delivering-for-women-children-and-adolescents-during-and-after-covid-19/.”

Schäferhoff, M., et al. Funding for sexual and reproductive health and rights in low-and middle-income countries: threats, outlook and opportunities [Internet]. Geneva: The Partnership for Maternal, Newborn & Child Health; 2019 [cited 2022 Feb 2]. Available from: https://pmnch.who.int/docs/librariesprovider9/meeting-reports/srhr_forecast.

Shepard, D. S., et al. (2003). “Cost-effectiveness of USAID’s regional program for family planning in West Africa.” Stud Fam Plann 34(2): 117–126.

Starrs, A. M., et al. (2018a). “Accelerate progress-sexual and reproductive health and rights for all: report of the Guttmacher-Lancet Commission.” Lancet 391(10140): 2642–2692.

Starrs, A. M., et al. (2018b). “Accelerate progress—sexual and reproductive health and rights for all: report of the Guttmacher– Lancet Commission.” The Lancet 391(10140): 2642–2692.

Sumner, A. and J. Glennie (2015). “Growth, Poverty and Development Assistance: When Does Foreign Aid Work?” Global Policy 6(3): 201–211.

Sundewall, J. and K. Sahlin-Andersson (2006). “Translations of health sector SWAps--a comparative study of health sector development cooperation in Uganda, Zambia and Bangladesh.” Health Policy 76(3): 277–287.

Taylor, E. M., et al. (2013). “The impact of official development aid on maternal and reproductive health outcomes: a systematic review.” PLoS One 8(2): e56271.

The Partnership for Maternal, N. C. H. (2019). Funding for sexual and reproductive health and rights in low- and middle-income countries; threats, outlook and opportunities.

Toseef, M. U., et al. (2019). “How Effective Is Foreign Aid at Improving Health Outcomes in Recipient Countries?” Atlantic Economic Journal 47(4): 429–444.

United Nations Population Fund (2014). Programme of Action of the International Conference on Population and Development. 20th Anniversary edition. New York, NY, United Nations Population Fund.

United Nations Population Fund (2019). Sexual and Reproductive Health and Rights: An essential element of Universal Health Coverage. Background document for the Nairobi summit on ICPD25 - Accelerating the promise. New York, NY, UNFPA.

Wayoro, D. and L. Ndikumana (2020). “Impact of development aid on infant mortality: Micro-level evidence from Côte d’Ivoire.” African Development Review 32(3): 432–445.

WHO (2023). World Health Statistics 2023: Monitoring health for the SDGs. Geneva, World Health Organization.

WHO, et al. (2019). Trends in maternal mortality 2000 to 2017. Geneva, World Health Organization.

Wilson, S. E. (2011). “Chasing Success: Health Sector Aid and Mortality.” World Development 39(11): 2032–2043.

Win, T. and Y. C. Cho (2018). “Exploring effectiveness of official development assistance: Comparison analysis of developing countries.” Journal of marketing thought 5(3): 22–32.

Wooldridge, J. M. (2010). Econometric Analysis of Cross Section and Panel Data. Cambridge, MA, MIT Press.

World Bank (2021). Walking the talk: Reimagining Primary Health Care After COVID-19. Washington, DC, The World Bank.

Yogo, U. T. and D. Mallaye (2015). “Health Aid and Health Improvement in Sub-Saharan Africa: Accounting for the Heterogeneity Between Stable States and Post-Conflict States.” Journal of International Development 27(7): 1178–1196.

Zhuang, H., et al. (2020). “Foreign Aid and Adolescent Fertility Rate: Cross-Country Evidence.” Journal of Globalization and Development 0(0).

## References

Cameron, A. C. and P. K. Trivedi (2005). Microeconometrics: Methods and Applications. New York, Cambridge University Press.

Frees, E. W. (2004). Longitudinal and Panel Data: Analysis and Applications in the Social Sciences. Cambridge, UK, Cambridge University Press.

Kripfganz, S. (2020). Generalized method of moments estimation of linear dynamic panel-data models. 2020 Stata conference 14, Stata Users Group.

